# Developing a research ready population-scale linked data ethnicity-spine in Wales

**DOI:** 10.1101/2022.11.28.22282810

**Authors:** Ashley Akbari, Fatemeh Torabi, Stuart Bedston, Emily Lowthian, Hoda Abbasizanjani, Richard Fry, Jane Lyons, Rhiannon K Owen, Kamlesh Khunti, Ronan A Lyons

## Abstract

**Introduction:** Ethnicity information is recorded routinely in electronic health records (EHRs); however, to date, there is no national standard or framework for harmonisation of the existing records.

**Methods and analysis:** The national ethnicity-spine uses anonymised individual-level population-scale ethnicity data from 26 EHR available through the Secure Anonymised Information Linkage (SAIL) Databank. A total of 46 million ethnicity records for 4,297,694 individuals in Wales-UK over 22 years (between 2000 and 2021) have been compiled in a harmonised, deduplicated longitudinal research ready data asset. We serialised this data and compared distribution of records over time for four selection approaches (*Latest, Mode, Weighted-Mode* and *Composite*) across age bands, sex, deprivation quintiles, health board, and residential location, against the ONS census 2011. The distribution of the dominant group (White) is minimally affected based on the four different selection approaches. Across all other ethnicity categorisations, the Mixed group was most susceptible to variation in distribution depending on the selection approach used and varied from a 0.6% prevalence across the *Latest* and *Mode* approach to a 1.1% prevalence for the *Weighted-Mode*, compared to the 3.1% prevalence for the *Composite* approach. Substantial alignment was observed with ONS census with the *Latest* group method (kappa= 0.68, 95% CI [0.67,0.71]) across all sub-groups.

**Conclusion:** We provides a reproducible EHR based resource enabling the investigation and evaluation of health inequalities related to ethnic groups in Wales. This generalisable method informs opportunities for the transferability of this methodology across the UK to platforms with comparable routine data sources.

**Ethics and dissemination:** This work was supported by the Con-COV team funded by Medical Research Council, Health Data Research UK, ADR Wales funded by ADR UK through the Economic and Social Research Council, and the Wales COVID-19 Evidence Centre, funded by Health and Care Research Wales.

Strengths and limitations of this study

- This is the first comprehensive approach for retrieving ethnicity records from linked EHR.
- The methodology presented here creates an anonymised longitudinal, population-scale individual-level linked ethnicity spine for the population of Wales.
- Enabling investigation and evaluation of health inequalities related to ethnic groups in Wales.
- Ethnicity spine is derived from 26 data sources across 22 years, complementing existing ethnicity data in census by providing a reproducible maintainable RRDA.
- As this is a data driven approach, the algorithm is limited to the reported ethnicity and actual diversity of the population.

**Article summary:** 

## Introduction

Describing disease dynamics across and within different subgroups of a population and addressing ethnic health disparities in the context of population health and biomedical research is of high importance due to its potential to depict areas of health inequalities and inform decisions towards a fair distribution of services. Recent debates around how diversity in data can enable generalisable research findings highlights to a greater extent the value of internal and external data diversity can provide to research (1). Improving existing ethnicity measures and strategies for enriching these data for research has become an urgent public health priority, specifically post-observation of disparities during the COVID-19 pandemic (2,3). This improvement can be made through harnessing the available ethnicity information in routinely collected health data. While reports of COVID-19 outcomes demonstrated severe outcomes for minority ethnic groups (4) (4–8), ethnicity is not always consistently recorded, nor are they available across all data sources and for the entire population. The availability of ethnicity data directly affects our ability to describe differences for a variety of health and social outcomes (2). Understanding the dynamics of COVID-19 and its outcomes have further revealed the enduring disparities experienced by ethnic minority groups and the need for improvements in the readiness of these data (9).

The importance of research readiness for investigating the effectiveness of interventions across and within different ethnic groups emphasises the value of ethnicity data. The availability of ethnicity data across the UK has been documented (10). Enabling ethnicity level characterisation of the population has been highlighted across various disease groups (11), in previous pandemics (12) and amongst the elderly (13). Research into the inequalities faced by different ethnic groups has continually referenced the diverse manifestation of coded ethnicity in data, including clinical and administrative data sources, and its impact on the researchers’ ability to estimate disparities (9,14). While the census provides a current snapshot of the geographical distribution of ethnicity records across the population, the culmination of the census data from 2011 means that embracing the recapturing opportunity through linkage of routinely collected ethnicity information in EHR data provides a current measure at any given time point. There is current evidence of crude approaches to processing ethnicity data, for example, the label ‘South Asian’ often includes the subpopulation groups, who are often culturally and behaviourally different from each other, e.g. Bangladeshi, Pakistani and Indian (9,15). Stakeholder groups have also emphasised that the data collected on ethnicity should be ‘strengthened’ towards enabling the inclusion of all ethnicities in our research (7).

Data diversity provides a unique opportunity to access recorded ethnicity across multiple data sources and permits the harmonisation of approaches to examine the consistency of research outputs and conclusions, which contributes to an improved understanding of health outcomes in marginalised ethnicities. Here, we aimed to address the urgent need for improved harmonised ethnicity data. We assessed the availability, record completeness and distribution of ethnicity records across EHRs and administrative data sources for various approaches in grouping and assignment of ethnicity. Our work is a direct response to the national need for these data, specifically in the context of health (NuffieldTrust, 2020; Pareek et al., 2020).

## Methods

### Study design and data sources

All anonymised individual-level, population-scale data sources within the Secure Anonymised Information Linkage (SAIL) Databank trusted research environment (TRE) (16,17) were assessed for the availability and completeness of ethnicity records. We used longitudinal records spanning 22 years, with the earliest records starting from the 1^st^ January 2000 and the most complete range of data providing full-year data up to the 31^st^ December 2021 (See Supplementary Figure 2 for data source range). Ethnicity records were extracted from 26 EHRs and administrative data sources based on their associated available coverage contributing to the 22 years longitudinal study window (Table 1 for the number and distribution of ethnicity records in each data source). Contributing data sources had various update frequencies ranging from daily flows to quarterly flows (Figure 1 – Supplementary Table 1 and Supplementary Figure 2 for further information on contributing data sources).

**Table 1.** Data source range, number of individual and total ethnicity records in each contributing data source (breakdown of ethnic group is presented using ONS categorisation).

| No. | Data source | Start date | End date | Number of people | Number of ethnicity records | Asian | Black | Mixed | Other | White |
| --- | --- | --- | --- | --- | --- | --- | --- | --- | --- | --- |
| 1 | BREC | 2000-01-01 | 2021-04-19 | 3,787 | 3,787 | 65 | 36 | 76 | 20 | 3,590 |
| 2 | CARS | 2000-01-03 | 2018-12-31 | 17,581 | 19,065 | 336 | 189 | 167 | 154 | 16,735 |
| 3 | CCDS | 2006-04-01 | 2021-12-31 | 90,303 | 108,100 | 507 | 151 | 107 | 95 | 89,493 |
| 4 | CENW | 2011-03-27 | 2011-03-27 | 2,663,411 | 2,663,726 | 55,865 | 13,207 | 26,001 | 10,150 | 2,558,451 |
| 5 | CNIS | 2000-01-01 | 2021-12-31 | 43,998 | 127,854 | 119 | 37 | 86 | 103 | 43,653 |
| 6 | CTTP | 2020-06-01 | 2021-12-01 | 364,093 | 444,833 | 8,526 | 2,230 | 6,868 | 3,689 | 342,828 |
| 7 | CVLF | 2019-10-16 | 2021-12-31 | 1,880,259 | 11,225,043 | 55,307 | 18,556 | 30,487 | 14,984 | 1,779,094 |
| 8 | CVVD | 2019-03-03 | 2021-12-31 | 1,579,429 | 4,164,721 | 39,117 | 10,196 | 12,940 | 20,494 | 1,496,818 |
| 9 | CYFI | 2000-01-01 | 2020-12-31 | 11,188 | 136,953 | 343 | 49 | 65 | 132 | 10,655 |
| 10 | DSCW | 2020-06-01 | 2020-06-01 | 24,693 | 24,693 | 214 | 283 | 244 | 127 | 23,825 |
| 11 | EDDS | 2008-04-05 | 2021-12-31 | 762,737 | 2,706,047 | 48,669 | 2,595 | 3,909 | 3,361 | 719,605 |
| 12 | EDUW | 2003-01-01 | 2020-01-01 | 1,124,234 | 8,121,162 | 26,650 | 10,934 | 34,707 | 14,971 | 1,053,707 |
| 13 | HWRA | 2020-01-31 | 2021-12-31 | 111,384 | 2,139,617 | 5,617 | 1,369 | 1,322 | 134 | 103,173 |
| 14 | ICNC | 2009-04-01 | 2021-06-30 | 100,328 | 121,401 | 864 | 292 | 210 | 248 | 98,802 |
| 15 | LACW | 2003-03-31 | 2021-03-31 | 12,451 | 61,943 | 303 | 249 | 497 | 264 | 11,408 |
| 16 | MIDS | 2014-03-31 | 2021-12-31 | 127,681 | 314,224 | 5,697 | 2,222 | 2,163 | 2,376 | 115,737 |
| 17 | NCCH | 2000-01-01 | 2021-10-08 | 609,453 | 126,6931 | 19,686 | 7,904 | 16,043 | 9,879 | 557,264 |
| 18 | NHSO | 2020-02-01 | 2021-12-31 | 14,852 | 26,273 | 103 | 26 | 87 | 31 | 14,605 |
| 19 | NSWD | 2014-03-31 | 2019-03-31 | 40,100 | 40,261 | 448 | 131 | 190 | 132 | 39,200 |
| 20 | OPRD | 2000-01-10 | 2021-12-31 | 822,337 | 3271,792 | 11,039 | 4,470 | 4,463 | 4,064 | 800,292 |
| 21 | PEDW | 2000-01-01 | 2021-12-31 | 1,194,215 | 5335,310 | 15,060 | 12,120 | 6,698 | 9,596 | 1,159,297 |
| 22 | SACT | 2001-01-15 | 2021-12-31 | Masked | 18,690 | 35 | <10 | 34 | 31 | 13,456 |
| 23 | SMDS | 2000-01-01 | 2021-12-31 | 82,167 | 217,551 | 574 | 434 | 713 | 154 | 80,605 |
| 24 | SWAC | 2019-09-01 | 2020-09-01 | 49,016 | 90,273 | 311 | 103 | 375 | 116 | 48,144 |
| 25 | WASD | 2015-08-31 | 2021-12-31 | 342,169 | 592,144 | 3,173 | 546 | 4,795 | 274 | 338,075 |
| 26 | WLGP | 2000-01-01 | 2021-12-31 | 1,834,740 | 2,825,204 | 95,156 | 27,663 | 23,720 | 48,192 | 1,666,779 |

**Figure 1.**
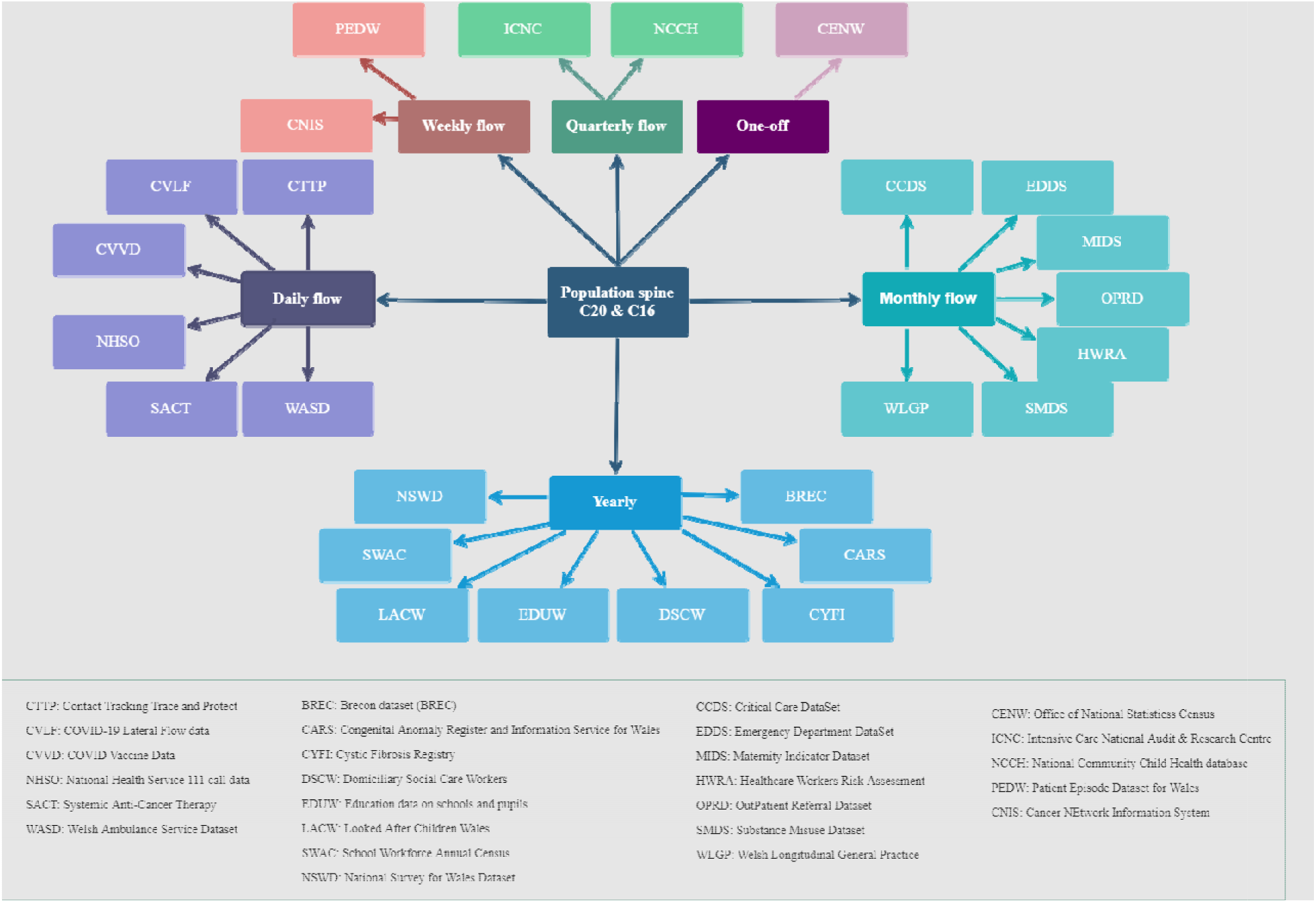
data sources used to create the harmonised ethnicity spine for Wales (data sources presented in update frequency).

Our methodology extracts ethnicity data from these 26 data sources and compiles the longitudinal, varied ethnicity values into a harmonised, research ready data asset (RRDA) using a Common Data Model (CDM). The steps in creating the population-spine ethnicity RRDA involved: record extraction, where all existing ethnicity records for each individual are extracted from the 26 data sources over time and gathered in a longitudinal dataset, followed by cleaning and harmonisation into ethnic groups using two different categorisation methods within each data source (Figure 2.A - we refer to this stage as categorisation hereafter), and finally serialising these records into a single row per individual using four different selection approaches to assign a single ethnicity to each individual (we refer to this stage as record selection hereafter). All longitudinal ethnicity records are preserved and changes over time for each individual are accessible, enabling the allocation of an individual-level ethnic grouping at any point in time to any anonymised linkable individual records. Approximately 4% of the Welsh population had two contradicting ethnicity records over time. In the record extraction stage: all data sources have only the records extracted from each respective data source that meets the required linkage quality (i.e. a match percentage of 95% quality or higher) and the date of the record occurring within the desired study window for all available longitudinal records, and then for each data source, all rows are extracted which corresponds to appropriate ethnicity values reviewed by clinical and domain experts. Once extracted, each data source has all the available records organised longitudinally and all values are harmonised into two different ethnicity categorisations consisting of five and nine ethnicity categories: I) five ethnic group categorisation consists of: ‘*White’, ‘Asian’, ‘Black’, ‘Mixed’, ‘Other’* as proposed by the Office for National Statistics (18) (referred to as ONS hereafter) and II) nine ethnic groups categorisation consists of: ‘*White’, ‘Chinese’, ‘Bangladeshi’, ’ Pakistani’, ’ Indian’, ‘Black African’, ‘Black Caribbean’, ‘Mixed’, ‘Other’* as proposed for use by the New and Emerging Respiratory Virus Threats Advisory Group (NERVTAG) (referred to as NER hereafter) (9) (Figure 2.A and Supplementary 4 for categorisation and harmonisation rules in each data source). Where we found multiple ethnicity recordings for an individual in various sources or over time, all records are ordered sequentially based on the date of recorded ethnicity. Finally, to reshape and process the data into one row per person, in the selection stage, the assignment of a single ethnic group to each individual occurs based on the following four selection approaches:

**Figure 2.**
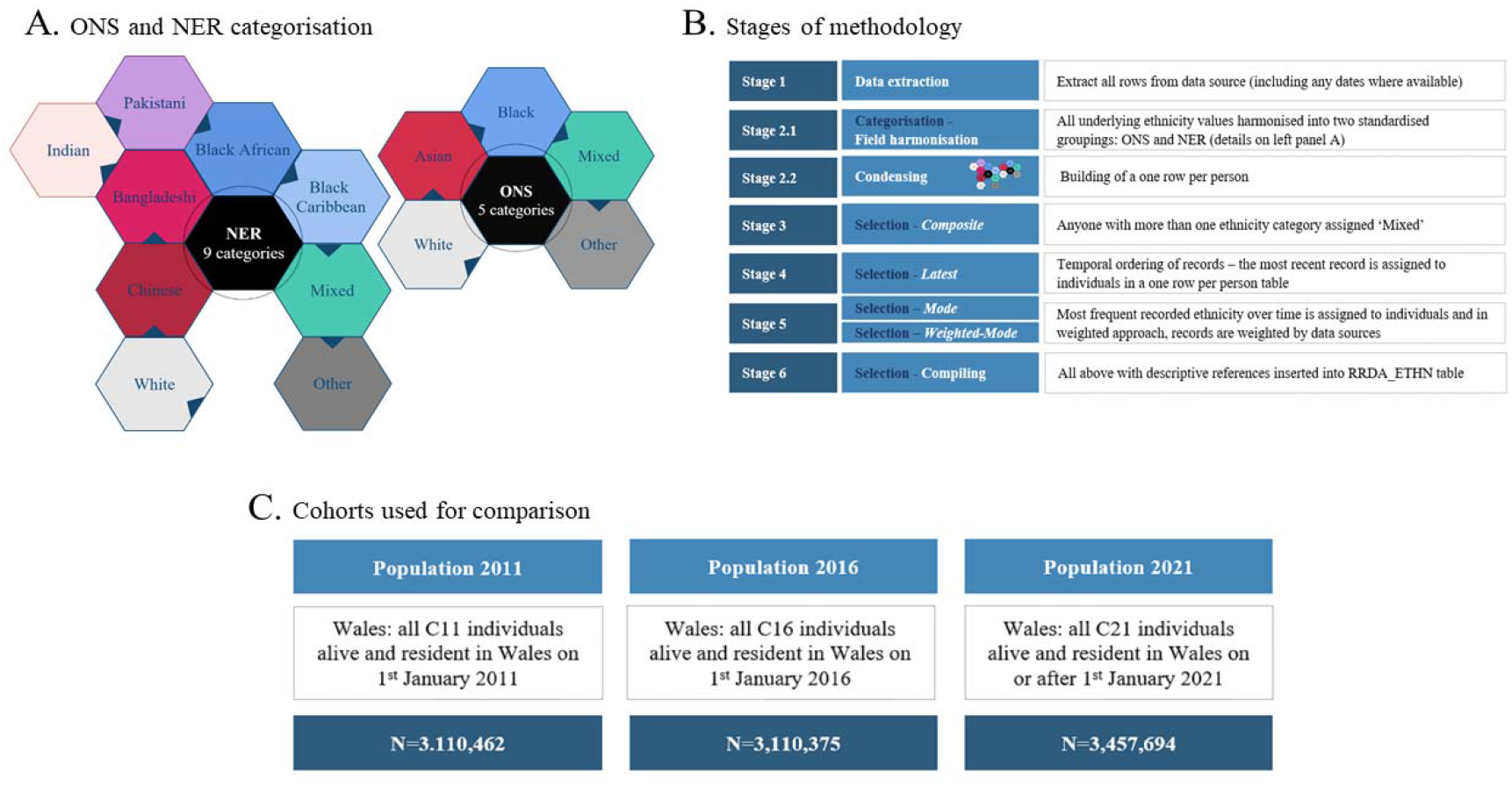
Ethnic group categorisations, stages of methodology, and cohorts used for comparing records over time.

1. *Latest*: The most recent ethnic group available in any of the available data sources is assigned to an individual as their ethnic group.
2. *Mode*: The most frequent recorded ethnic group (the mode) is extracted from all available data sources and is assigned to an individual as their ethnic group.
3. *Weighted-mode*: The mode is extracted, with appropriate weights derived from the ONS 2011 Census population composition is assigned to an individual as their ethnic group.
4. *Composite*: The ethnicity of those individuals who have declared more than one distinct ethnic group over time is assigned to the Mixed ethnic group (due to the nature of the *composite* method and no hierarchy or prioritisation given to which ethnic group should be assigned, where different categories are found for an individual over time, the *composite* method would assign all of the 4% of individuals with contradicting records into the Mixed ethnic category).

Post assessment, harmonisation and creation of a CDM for population-scale data on ethnicity, we summarise all this information in our ethnicity spine RRDA, to provide harmonised ethnic group information suitable for data linkage, maintained over time, which is available to researchers using the SAIL Databank. All available records within and across data sources are harmonised based on clinical and domain expertise into a CDM (see Figure 2.B for the steps involved in the methodology and Supplementary Table 3 for the metadata of RRDA tables).

### Statistical Analysis

Three cohorts consisting of individuals alive and living in Wales in 2011 (census year), 2016 (mid-study) and 2021 (end-of-study) (Figure 2.C) were used to compare record completeness over time from the year the census was completed in 2011. Using the ONS and NER ethnic group categorisations, we compare the distribution of records for four different selection approaches across age bands, sex, deprivation quintiles, geographic health board and residential areas. Cohen’ s kappa coefficient was calculated to assess the inter-rater reliability between the *Latest* ethnicity approach and census records for the 2011 population (19). We report the inter-rater coefficients for those with known ethnicity status in both the census and the *Latest* approach. Where 0 is defined as no agreement between the two sources and 1 is the maximal agreement of records across the two sources (See Supplementary Figure 1 for detail on agreement scales). Age is calculated based on the week of birth of individuals on 1^st^ January for each population or later if born afterwards. Week of birth, sex, health board and residential information of all individuals are sourced from Welsh Demographic Dataset (20) and in the COVID-19 e-cohort known as C20 (21).

## Results

### Cohort curation

A total of 46 million ethnicity records were recorded between 2000 and 2021 across the 26 EHRs and administrative data sources for 4,297,694 individuals (see Table 1, Supplementary Figure 2 for distribution of existing records in each contributing data source). We summarised the data for the population at three inception dates: 1^st^ January 2011 (C11) for 3,110,462 individuals, 1^st^ January 2016 (C16) for 3,110,375 and 1^st^ January 2021 (C21) for 3,457,697. Each cohort contains records of individuals alive and residing in Wales on the 1^st^ January of each year.

### Evaluation of different selection approaches for ethnic group categorisation

A comparison of four selection approaches (*Latest, Mode, Weighted-Mode*, and *Composite*) showed that the distribution of C21 records varied based on approach: for the Mixed ethnic group using the *Latest* and *Mode* method resulted in 1.3% of the population being assigned to Mixed group while using *Weighted-Mode* resulted in 2.0% assignment, this has raised to 6.0% in the Composite approach, in which anyone with more than one distinct ethnic group category is assigned to the Mixed group (ONS-Mixed-*Latest*=1.3%, ONS-Mixed-*Mode*=1.3%, ONS-Mixed-*Weighted-Mode*=2.0%, ONS-Mixed-*Composite*=6.0%) similar patterns were observed for the NER categorisation (NER-Mixed-*Latest*=1.3%, NER-Mixed-*Mode*=1.3%, NER-Mixed-*Weighted-Mode*=1.9%, NER-Mixed-*Composite*=6.3%) (Table 2). Breaking down the records for each approach by sex and age for the two ONS and NER categorisations showed that the White ethnic group was recorded as the highest proportion, and its distribution was minimally affected by the approach used (Female-White-ONS-*Latest*=44%, Female-White-ONS-*Mode*=44%, Female-White-ONS-*Weighted*-*Mode*=43%, Female-White-ONS-*Composite*=43% & Female-White-NER-*Latest*=44%, Female-White-NER-*Mode*=44%, Female-White-NER-*Weighted-Mode*=43%, Female-White-NER-*Composite*=43%). Across all the minority ethnic groups, the Mixed group was the group for which the record distribution varied from the *Latest* and *Mode* compared to the *Composite* approach (Female-Mixed-ONS-*Latest*=0.6%, Female-Mixed-ONS-*Mode*=0.6%, Female-Mixed-ONS-*Weighted*-*Mode*=1.1%, Female-Mixed-ONS-*Composite*=3.1% & Female-Mixed-NER-*Latest*=0.6%, Female-Mixed-NER-*Mode*=0.6%, Female-Mixed-NER-*Weighted-Mode*=1.0%, Female-Mixed-NER-*Composite*=3.2%). Whilst for the rest of the ethnic groups, the distribution of records across age bands and sex were similar for four approaches across both ONS and NER categorisations, we illustrated these across all age bands in Figures 3 and 4 (Supplementary Table 2A & 2B for distribution of records over each age band and selection method). We observed an improved categorisation achieved by the *Latest* method followed by the *Mode* selection method in nine NER categorisation.

**Table 2.** characteristics of the cohort based on ONS and NER categorisation.

| Characteristics | Latest |  |  | Mode |  | Weighted Mod |  | Composite |  |
| --- | --- | --- | --- | --- | --- | --- | --- | --- | --- |
|  | Ethnic group |  |  | Ethnic group |  | Ethnic group |  | Ethnic group |  |
|  | N = 3,457,694 |  |  | N = 3,457,694 |  | N = 3,457,694 |  | N = 3,457,694 |  |
|  | N (%) |  |  | N (%) |  | N (%) |  | N (%) |  |
| Ethnic Group (ONS) |  |  |  |  |  |  |  |  |  |
| White | 3,080,277 | (89.0%) |  | 3,093,752 | (89.0%) | 2,989,648 | (86.0%) | 2,987,245 | (86.0%) |
| Asian | 110,809 | (3.2%) |  | 100,502 | (2.9%) | 133,258 | (3.9%) | 72,564 | (2.1%) |
| Black | 30,880 | (0.9%) |  | 30,817 | (0.9%) | 41,559 | (1.2%) | 19,036 | (0.6%) |
| Mixed | 43,672 | (1.3%) |  | 43,422 | (1.3%) | 70,880 | (2.0%) | 207,627 | (6.0%) |
| Other | 38,724 | (1.1%) |  | 35,869 | (1.0%) | 69,017 | (2.0%) | 17,890 | (0.5%) |
| Unknown | 153,332 | (4.4%) |  | 153,332 | (4.4%) | 153,332 | (4.4%) | 153,332 | (4.4%) |
| Ethnic Group (NER) |  |  |  |  |  |  |  |  |  |
| White | 3,080,277 | (89.0%) |  | 3,093,817 | (89.0%) | 2,987,420 | (86.0%) | 2,987,245 | (86.0%) |
| Bangladeshi | 25,780 | (0.7%) |  | 16,243 | (0.5%) | 55,675 | (1.6%) | 9,204 | (0.3%) |
| Chinese | 17,970 | (0.5%) |  | 17,851 | (0.5%) | 20,302 | (0.6%) | 13,703 | (0.4%) |
| Indian | 25,058 | (0.7%) |  | 25,190 | (0.7%) | 27,078 | (0.8%) | 17,075 | (0.5%) |
| Pakistani | 17,035 | (0.5%) |  | 17,043 | (0.5%) | 20,287 | (0.6%) | 11,030 | (0.3%) |
| Black African | 21,578 | (0.6%) |  | 22,003 | (0.6%) | 32,671 | (0.9%) | 9,904 | (0.3%) |
| Black Caribbean | 4,730 | (0.1%) |  | 4,083 | (0.1%) | 7,145 | (0.2%) | 1,135 | (<0.1%) |
| Mixed | 43,672 | (1.3%) |  | 43,572 | (1.3%) | 66,895 | (1.9%) | 217,059 | (6.3%) |
| Other | 68,262 | (2.0%) |  | 64,560 | (1.9%) | 86,889 | (2.5%) | 38,007 | (1.1%) |
| Unknown | 153,332 | (4.4%) |  | 153,332 | (4.4%) | 153,332 | (4.4%) | 153,332 | (4.4%) |

**Figure 3.**
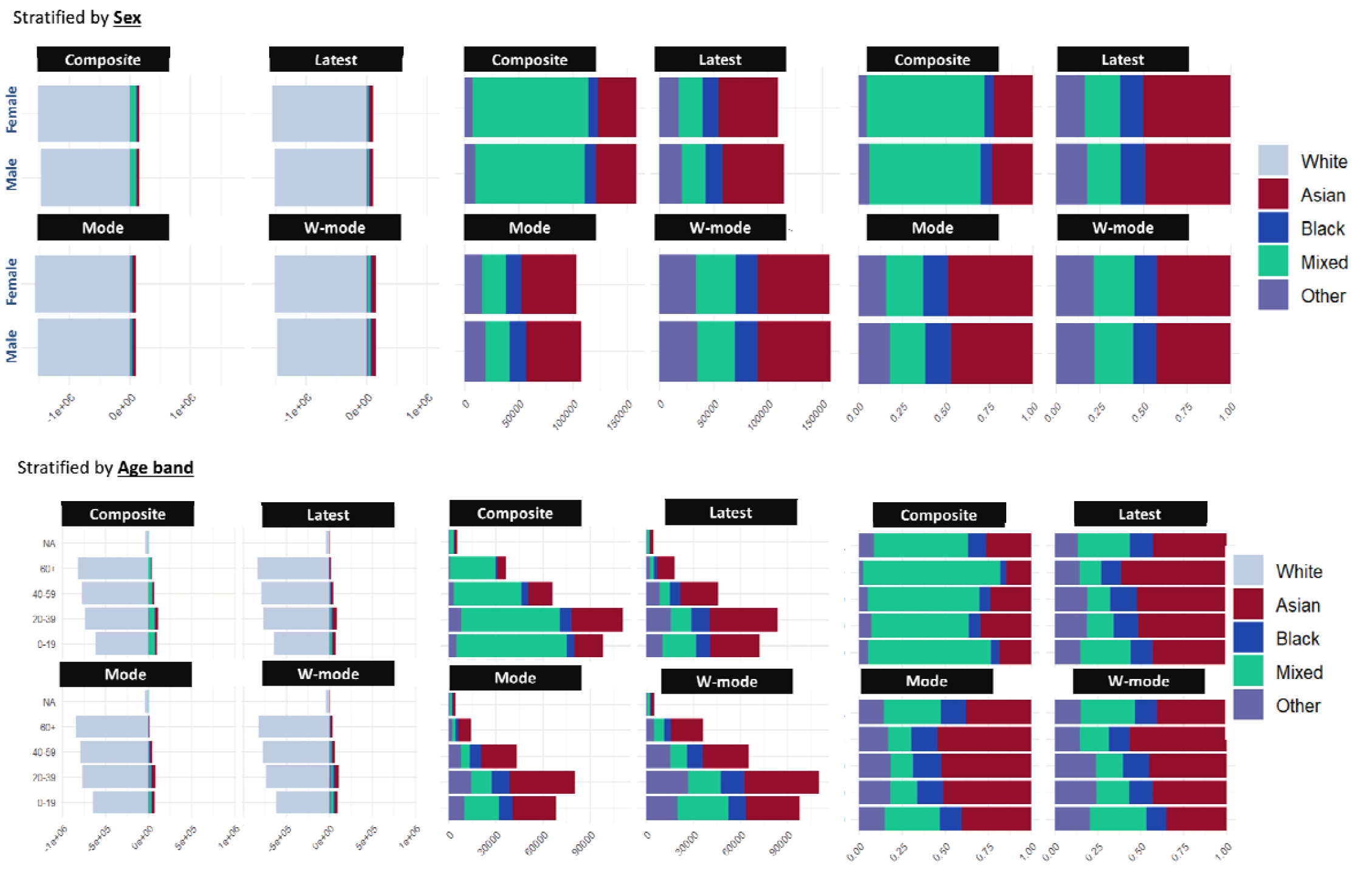
distribution of ethnic groups across sex & age groups for ONS categorisation for four different retrieval method approaches.

**Figure 4.**
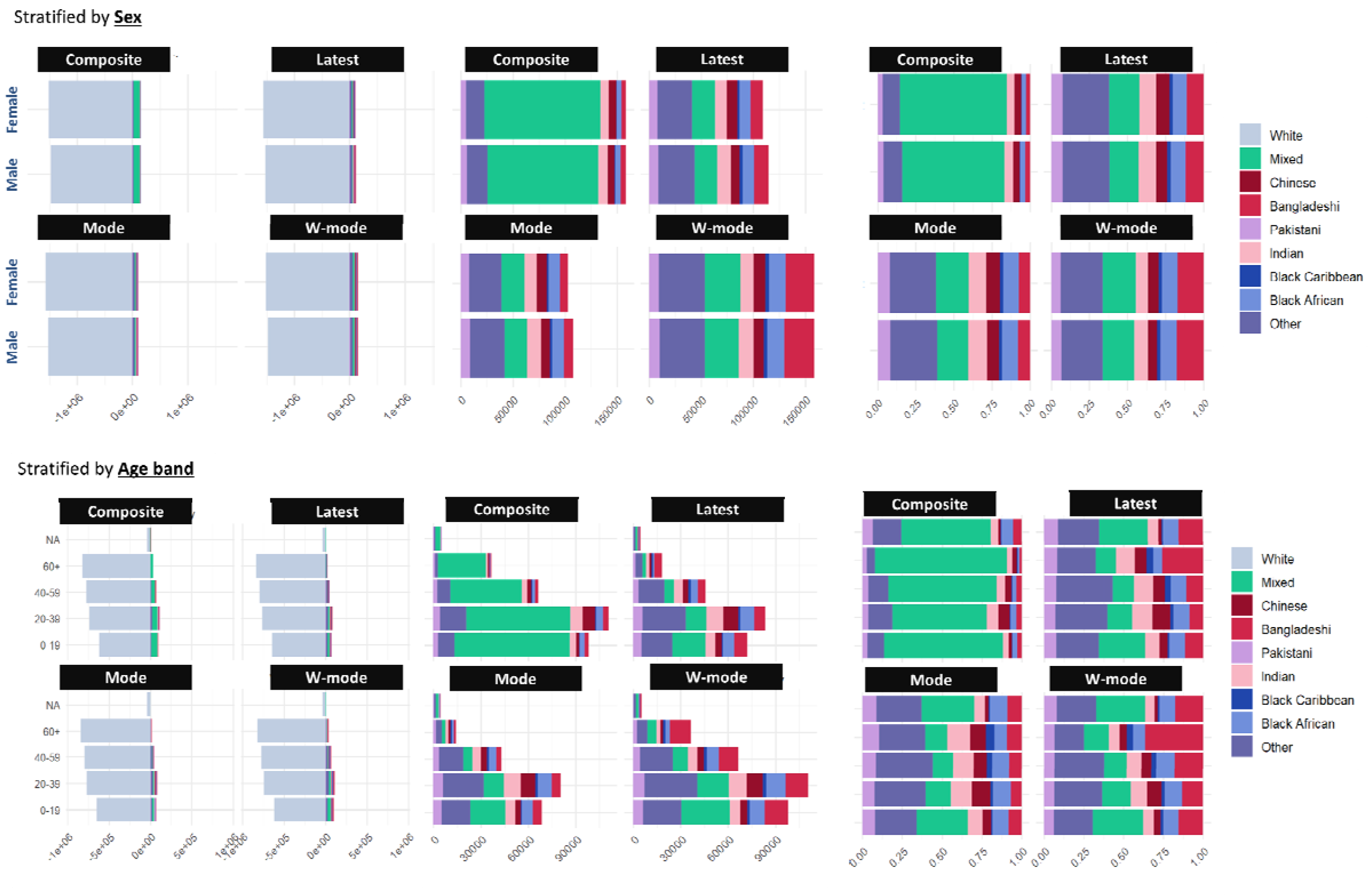
distribution of ethnic groups across sex & age groups for NER categorisation for four different retrieval method approaches.

In 2011, census data provided the most complete distribution of ethnicity records, where 2,546,403 (82%) of the C11 population had a recorded ethnicity in the census, leaving 18% of the population with an unknown ethnic group across both ONS and NER categorisation. The contribution of EHR overtime increases from 10% in 2011 to 31% in 2021, resulting in a 13·6% reduction of missingness of an available ethnic group over time for both ONS and NER categorisation with the *Latest* method. This is in the background of changes in the population, including births and moving in and out of Wales (Table 3 and Supplementary Figure 3). A comparison of individual-level category assignment between those who had a record both in the ethnicity-spine and in the census (Eliminating 1,663,775 individuals who had an unknown ethnicity either in the census or in the C11 cohort) showed substantial agreement between the census and ethnicity-spine RRDA (Cohen’ s kappa=0·68, 95%CI [0·67, 0·71])). Post 2011, record completeness improved by 2·8% from 2016 to 2021, with agreement level of 0·46% across the two years (Cohen’ s kappa=0·46, 95% CI [0·44, 0·49]) (see Figure 5 and Supplementary Figure 4 for Cohen’ s kappa matrix).

**Table 3.** record comparison for ONS categorisation between the census and the Latest method approaches (Note: ethnic group results are presented specifically for ONS and NER categorisation).

| Characteristics | Census-ETHN 2011<br>N = 3,110,462 | Latest-ETHN<br>2011<br>N = 3,110,462 | Latest-ETHN 2016<br>N = 3,110,375 | Latest-ETHN 2021<br>N = 3,457,694 |
| --- | --- | --- | --- | --- |
| **Ethnic group source** |  |  |  |  |
| Census | 2,546,403 (81.9%) | 2,546,403 (81.9%) | 2,331,911 (75.0%) | 2,243,846 (64.9%) |
| EHR | --- | 313,211 (10.1%) | 554,179 (17.8%) | 1,060,516 (30.7%) |
| Unknown | 564,059 (18.1%) | 250,848 (8.1%) | 224,285 (7.2%) | 153,332 (4.4%) |
| **Ethnic group (ONS)** |  |  |  |  |
| White | 2,451,348 (78.8%) | 1,671,303 (53.7%) | 2,747,991 (88.3%) | 3,080,277 (89.1%) |
| Asian | 50,054 (1.6%) | 40,045 (1.3%) | 64,997 (2.1%) | 110,809 (3.2%) |
| Black | 11,823 (0.4%) | 13,730 (0.4%) | 19,491 (0.6%) | 30,880 (0.9%) |
| Mixed | 24,313 (0.8%) | 19,352 (0.6%) | 32,777 (1.1%) | 43,672 (1.3%) |
| Other | 8,865 (0.3%) | 15,468 (0.5%) | 20,834 (0.7%) | 38,724 (1.1%) |
| Unknown | 564,059 (18.1%) | 1,350,564 (43.4%) | 224,285 (7.2%) | 153,332 (4.4%) |
| **Ethnic group (NER)** |  |  |  |  |
| White | 2,451,348 (78.8%) | 1,671,303 (53.7%) | 2,747,991 (88.3%) | 3,080,277 (89.1%) |
| Indian | 12,445 (0.4%) | 10,588 (0.3%) | 15,337 (0.5%) | 25,058 (0.7%) |
| Chinese | 7,928 (0.3%) | 6,262 (0.2%) | 10,439 (0.3%) | 17,970 (0.5%) |
| Bangladeshi | 8,630 (0.3%) | 7,952 (0.3%) | 11,454 (0.4%) | 25,780 (0.7%) |
| Pakistani | 9,836 (0.3%) | 6,059 (0.2%) | 12,169 (0.4%) | 17,035 (0.5%) |
| Black African | 2,441 (0.1%) | 9,733 (0.3%) | 11,158 (0.4%) | 21,578 (0.6%) |
| Black Caribbean | 7,458 (0.2%) | 1,689 (0.1%) | 4,920 (0.2%) | 4,730 (0.1%) |
| Mixed | 24,313 (0.8%) | 19,352 (0.6%) | 32,777 (1.1%) | 43,672 (1.3%) |
| Other | 22,004 (0.7%) | 26,960 (0.9%) | 39,845 (1.3%) | 68,262 (2.0%) |
| Unknownwn | 564,059 (18.1%) | 1,350,564 (43.4%) | 224,285 (7.2%) | 153,332 (4.4%) |
| **Age band** |  |  |  |  |
| 0-19 | 706,461 (22.7%) | 706,461 (22.7%) | 686,558 (22.1%) | 739,814 (21.4%) |
| 20-39 | 808,723 (26.0%) | 808,723 (26.0%) | 788,700 (25.4%) | 898,841 (26.0%) |
| 40-59 | 839,910 (27.0%) | 839,910 (27.0%) | 840,572 (27.0%) | 882,458 (25.5%) |
| 60+ | 755,368 (24.3%) | 755,368 (24.3%) | 794,545 (25.5%) | 881,107 (25.5%) |
| **Sex** |  |  |  |  |
| Male | 1,549,319 (49.8%) | 1,549,319 (49.8%) | 1,551,616 (49.9%) | 1,728,015 (50.0%) |
| Female | 1,561,143 (50.2%) | 1,561,143 (50.2%) | 1,558,759 (50.1%) | 1,729,679 (50.0%) |
| **Deprivation quintile** |  |  |  |  |
| 1-Most deprived | 622,879 (20.0%) | 622,879 (20.0%) | 631,347 (20.3%) | 658,913 (19.1%) |
| 2 | 619,359 (19.9%) | 619,359 (19.9%) | 619,099 (19.9%) | 638,925 (18.5%) |
| 3 | 627,624 (20.2%) | 627,624 (20.2%) | 624,789 (20.1%) | 643,624 (18.6%) |
| 4 | 618,525 (19.9%) | 618,525 (19.9%) | 615,938 (19.8%) | 630,614 (18.2%) |
| 5-Least deprived | 622,075 (20.0%) | 622,075 (20.0%) | 619,202 (19.9%) | 635,890 (18.4%) |
| **Local Health Board (HB)** |  |  |  |  |
| Aneurin Bevan University HB | 588,352 (18.9%) | 588,352 (18.9%) | 587,737 (18.9%) | 605,782 (19.5%) |
| Betsi Cadwaladr University HB | 689,758 (22.2%) | 689,758 (22.2%) | 683,671 (22.0%) | 701,543 (20.3%) |
| Cardiff and Vale University HB | 493,312 (15.9%) | 493,312 (15.9%) | 505,232 (16.2%) | 526,652 (15.2%) |
| Cwm Taf Morgannwg University HB | 444,806 (14.3%) | 444,806 (14.3%) | 444,300 (14.3%) | 459,118 (14.3%) |
| Hywel Dda University HB | 376,346 (12.1%) | 376,346 (12.1%) | 372,309 (12.0%) | 385,174 (12.2%) |
| Powys Teaching HB | 129,363 (4.2%) | 129,363 (4.2%) | 126,644 (4.1%) | 128,461 (3.7%) |
| Swansea Bay University HB | 388,525 (12.5%) | 388,525 (12.5%) | 390,482 (12.6%) | 401,236 (11.6%) |
| **Urban Rural classification** |  |  |  |  |
| Rural town and fringe | 412,275 (13.3%) | 412,275 (13.3%) | 410,122 (13.2%) | 419,900 (13.1%) |
| Rural town and fringe in a sparse setting | 120,097 (3.9%) | 120,097 (3.9%) | 117,362 (3.8%) | 118,480 (3.7%) |
| Rural village and dispersed | 205,555 (6.6%) | 205,555 (6.6%) | 201,231 (6.5%) | 204,993 (6.4%) |
| Rural village and dispersed in a sparse setting | 227,732 (7.3%) | 227,732 (7.3%) | 221,427 (7.1%) | 225,307 (7.0%) |
| Urban city and town | 2,086,855 (67.1%) | 2,086,855 (67.1%) | 2,102,605 (67.6%) | 2,179,684 (68.1%) |
| Urban city and town in a sparse setting | 57,948 (1.9%) | 57,948 (1.9%) | 57,628 (1.9%) | 59,602 (1.9%) |

**Figure 5.**
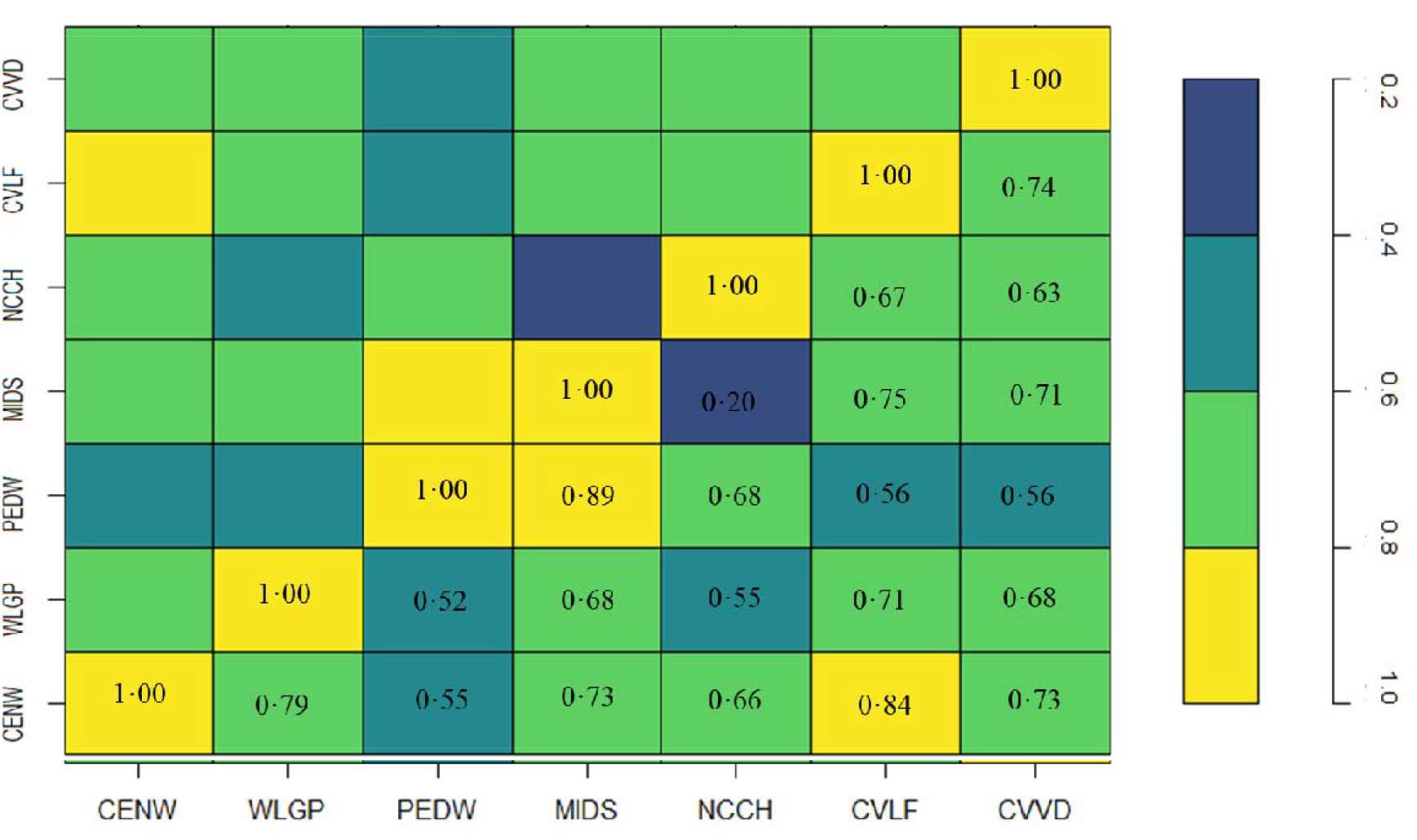
inter-rater coefficients between existing ethnic groups in Census 2011 (CENW), Welsh Longitudinal General Practice (WLGP), Patient Episode Dataset for Wales (PEDW), Maternity Indicator DataSet (MIDS), National Community Child Health data (NCCH), COVID-19 Lateral Flow data (CVLF) and COVID Vaccine Data (CVVD).

## Discussion

Our development of the population-scale ethnicity spine provides robust ethnicity measures to healthcare research in Wales and a template which can easily be deployed in other TREs in the UK and beyond. The development of the ethnicity CDM, presented in this study, involved categorising and cleaning data and values recorded across 26 longitudinal data sources over 22 years. This resulted in record completeness and recapturing of current ethnic group distribution for the population. Our approach also provides a CDM where variable names, contributing data sources, and categorisation is readily accessible to users, enabling transferability of the final output tables across projects. We report a 2.8% improvement in record completeness from 2016 to 2021, with a major contributor to this being the regular update and population-scale nature of recent COVID-19 related data sources, such as COVID-19 vaccination (CVVD) and lateral flow testing (CVLF) data sources, which have provided the opportunity of capturing ethnicity records across the entire Welsh population. While multiple data sources have contributed to achieving an accurate ethnicity status for each individual, we can see which data sources have contributed longitudinally and across the whole population, with no prioritisation applied, with records from every data source having equal value (See Supplementary Figure 2 for contributing datasets and the ranges over time).

We present a methodology for creating a longitudinal population-scale individual-level linked data ethnicity spine for the population of Wales. Additionally, to assess the robustness of our proposed methodology, we provide a comparison of different categorisation and selection approaches. We further summarised the contribution level of each data source and compared the EHR ethnicity with the ONS 2011 census over time, considering changes in the population dynamics due to births, deaths and migration. Our methodology showed that using the *Latest* method provides substantial agreement measure of ethnicity in the population when compared directly with the census (Cohen’ s kappa=0.68). Our methodology has demonstrated the benefits of including ethnic group information when understanding clinical outcomes at a population scale.

The continuous recapturing of records across EHRs ensures the underrepresentation of ethnic minority (EM) groups over time is addressed, leading to fewer erroneous results among other studies using this data. As we have shown during the COVID-19 pandemic, the availability of various data sources resulted in an opportunity for recapturing ethnicity records. The proposed method in this manuscript have shown usefulness in various research enabling investigations around health inequalities for minority ethnic groups (22–28). There are further recent reports on variations in outcomes for EM groups with documented effects on vaccine hesitancy (29) as well as post-vaccination COVID-19 deaths that were found to be higher among those of Pakistani or Indian background (30). Our study provides a population-scale national ethnicity CDM in the form of an RRDA to support ongoing activities for the assessment of health risks, behaviours and outcomes across the less representative subgroups of the population of Wales, enabling the evaluation of potential inequalities due to ethnic groups across the population of Wales. To date, this methodology has enabled national level reporting on vaccination uptake in the general population of Wales (24) as well as supporting strategic vaccination decisions for subgroups of the population such as healthcare workers (23).

We explored the effect of categorising people with inconsistent records throughout time into the Mixed ethnic group category through the *Composite* method. We have shown that this method skews records for descriptive analysis across the COVID-19 measure. We would like to further outline the limitations of the *Composite* method as demonstrated throughout, with the availability of more data over time contributing to the *Composite* method only by increasing the proportion of records in the Mixed group and the decrease in those individuals being assigned to a unique ethnic group category. As such, the *Composite* approach is only appropriate in certain study design circumstances with a fixed point in time, no updates in data coverage and a need to retain the knowledge of individuals that declare to more than one ethnic group over time as a single row per individual variable. We would like to acknowledge that although within the SAIL Databank, we had access to a collection of longitudinal data sources, the scope of our work is limited to the quality of records available in each data source. Further improvement in primary data collection, encouragement on accurate reporting of ethnicity, as well as further work to understand those people who do not disclose their ethnicity and who are unknown in data sources to be interacting with services is needed, especially in potential groups where inequalities are thought to be higher or certain groups who are potentially at greater risk of COVID-19 infection or outcomes such as health care providers or social workers. Our study demonstrates that the *Latest, Mode* and *Weighted-Mode* approaches all provide suitable categorisations for ethnic group data and the choice of ethnic group methodological approach chosen by any study down to their respective study design, with the standard choice from current studies across the UK being the *Latest* (Figures 3 & 4 - Table 3).

Our population-level CDM and methodology, utilising 22 years of longitudinal data in 26 data sources, provides a ground for investigating existing associations and health inequalities at the population level in Wales. It also informs opportunities for the transferability of this methodology across the UK to other data sources and TREs, platforms and systems which hold comparable routine data sources that contain ethnicity information, who can take the learning, harmonisation and implementation of this methodology and replicate it for wider use and further harmonisation and opportunities to implement UK wide inequalities research. The ethnicity spine RRDA is currently available to all COVID-19 researchers within the SAIL Databank to utilise towards COVID-19 research and evaluations, with further work planned to discuss with data owners around the opportunities to share this implemented RRDA within the SAIL Databank with all researchers in SAIL directly, following appropriate governance and application approvals.

## Data Availability

The anonymised individual-level data sources used in this study are available in the SAIL Databank at Swansea University, Swansea, UK, but as restrictions apply, they are not publicly available. All proposals to use SAIL data are subject to review by the independent Information Governance Review Panel (IGRP). Before any data can be accessed, approval must be given by the IGRP. The IGRP gives careful consideration to each project to ensure proper and appropriate use of SAIL data. When access has been granted, it is gained through a privacy-protecting safe haven and remote access system referred to as the SAIL Gateway. SAIL has established an application process to be followed by anyone who would like to access data via SAIL at: https://www.saildatabank.com/application-process/

## Author contributions

AA, RAL and KK were responsible for the conceptualisation, data curation, and development of the methodology. KK provided the ethnicity harmonisation of the various ethnicity values into the harmonised ONS and NER ethnic group categories. AA developed the methodology. All results are generated by FT and SB and reviewed by AA and RO. FT and AA developed the first draft of the paper. All authors have reviewed the work in various stages and approved the final version of this manuscript. The corresponding author confirms that all authors have reviewed and approved the final manuscript. AA and FT are the guarantors and had full access to the raw and derived study data.

## Acknowledgement

This work uses data provided by patients and collected by the NHS as part of their care and support. We would also like to acknowledge all data providers who make anonymised data available for research.

We wish to acknowledge the collaborative partnership that enabled the acquisition and access to the de-identified data, which led to this output. The collaboration was led by the Swansea University Health Data Research UK team under the direction of the Welsh Government Technical Advisory Cell (TAC) and includes the following groups and organisations: the Secure Anonymised Information Linkage (SAIL) Databank, Administrative Data Research (ADR) Wales, Digital Health and Care Wales (DHCW), Public Health Wales, NHS Shared Services Partnership and the Welsh Ambulance Service Trust (WAST). All research conducted has been completed under the permission and approval of the SAIL independent Information Governance Review Panel (IGRP) project number 0911.

## Public and patient involvement

This project is undertaken under a proposal submitted to the independent Information Governance Review Panel (IGRP), including members of the public (IGRP Project: 0911). Two public members contributed to the project’ s approval as part of the IGRP panel.

