## Supplementary Information for "Developing a research ready population-scale linked data ethnicity-spine in Wales"

Authorship list

Ashley Akbari * ^1^, Fatemeh Torabi * ^1^, Stuart Bedston ^1^, Emily Lowthian ^1,3^, Hoda Abbasizanjani ^1^, Rich Fry ^1^, Jane Lyons ^1^, Rhiannon Owen ^1^, Kamlesh Khunti ₸ ^2^, Ronan A. Lyons ₸ ^1^

^1^ Population Data Science, Swansea University Medical School, Faculty of Medicine, Health & Life Science, Swansea University, Swansea, Wales, UK

^2^ Diabetes Research Centre, University of Leicester, Leicester, LE5 4PW

^3^. Department of Education & Childhood, School of Social Sciences, Swansea University, Swansea, Wales, UK

* Authors have equally contributed

₸ Senior Authors

Supplementary Figure 1-inter-rater coefficient scales of agreement

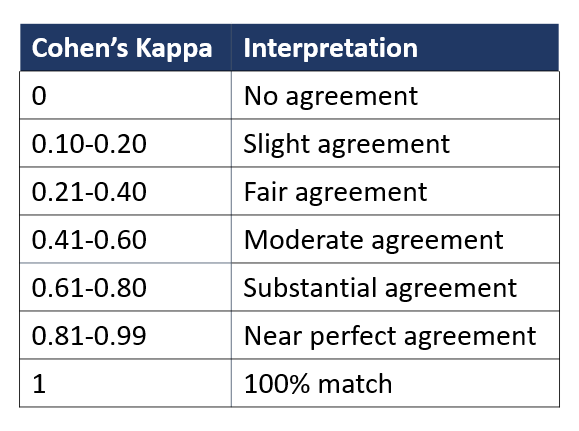

Supplementary Table 1 – name and coverage of data sources used in the creation of ethnicity spine for Wales

| **Data flow** | **Data Source** | **Full name** |
| --- | --- | --- |
| Daily | CTTP | Contact Tracking Trace and Protect |
| Daily | CVLF | COVID-19 Lateral Flow test |
| Daily | CVVD | COVID Vaccine Data |
| Daily | NHSO | NHS 111 Call data |
| Daily | SACT | Systemic Anti-Cancer Therapy |
| Daily | WASD | Welsh Ambulance Service Dataset |
| Weekly | CNIS | Cancer Network Information System Cymru (CaNISC) |
| Weekly | PEDW | Patient Episode Database for Wales |
| Quarterly | ICNC | ICNARC – Intensive Care National Audit & Research Centre |
| Quarterly | NCCH | National Community Child Health database |
| One-off | CENW | Office for National Statistics 2011 Census |
| Monthly | CCDS | Critical Care Data Set |
| Monthly | EDDS | Emergency Department Data Set |
| Monthly | MIDS | Maternity and childbirth Indicator DataSet |
| Monthly | HWRA | Healthcare Workers Risk Assessment |
| Monthly | OPRD | OutPatient Referral Dataset |
| Monthly | SMDS | Substance Misuse Dataset |
| Monthly | WLGP | Welsh Longitudinal General Practice |
| Yearly | BREC | Brecon dataset |
| Yearly | CARS | Congenital Anomaly Register and Information Services for Wales |
| Yearly | CYFI | Cystic Fibrosis Registry |
| Yearly | DSCW | Domiciliary Social Care Workers |
| Yearly | EDUW | Education data on schools and pupils |
| Yearly | LACW | Looked After Children Wales |
| Yearly | SWAC | School Workforce Annual Census |
| One-off | NSWD | National Survey for Wales Dataset |

**Supplementary Figure 2-longitudinal coverage of contributing data sources into ethnicity spine**

Ethnicity records have been extracted from Electronic Health Records (EHRs). While multiple data sources have contributed to achieving an accurate ethnic group for each individual, we can see which data sources have contributed the most in the following figures A and B.

*Source of ethnicity records over the years – from all data sources (*we sequentially remove high contributing data sources to visualise which data sources contribute ethnicity records over time).

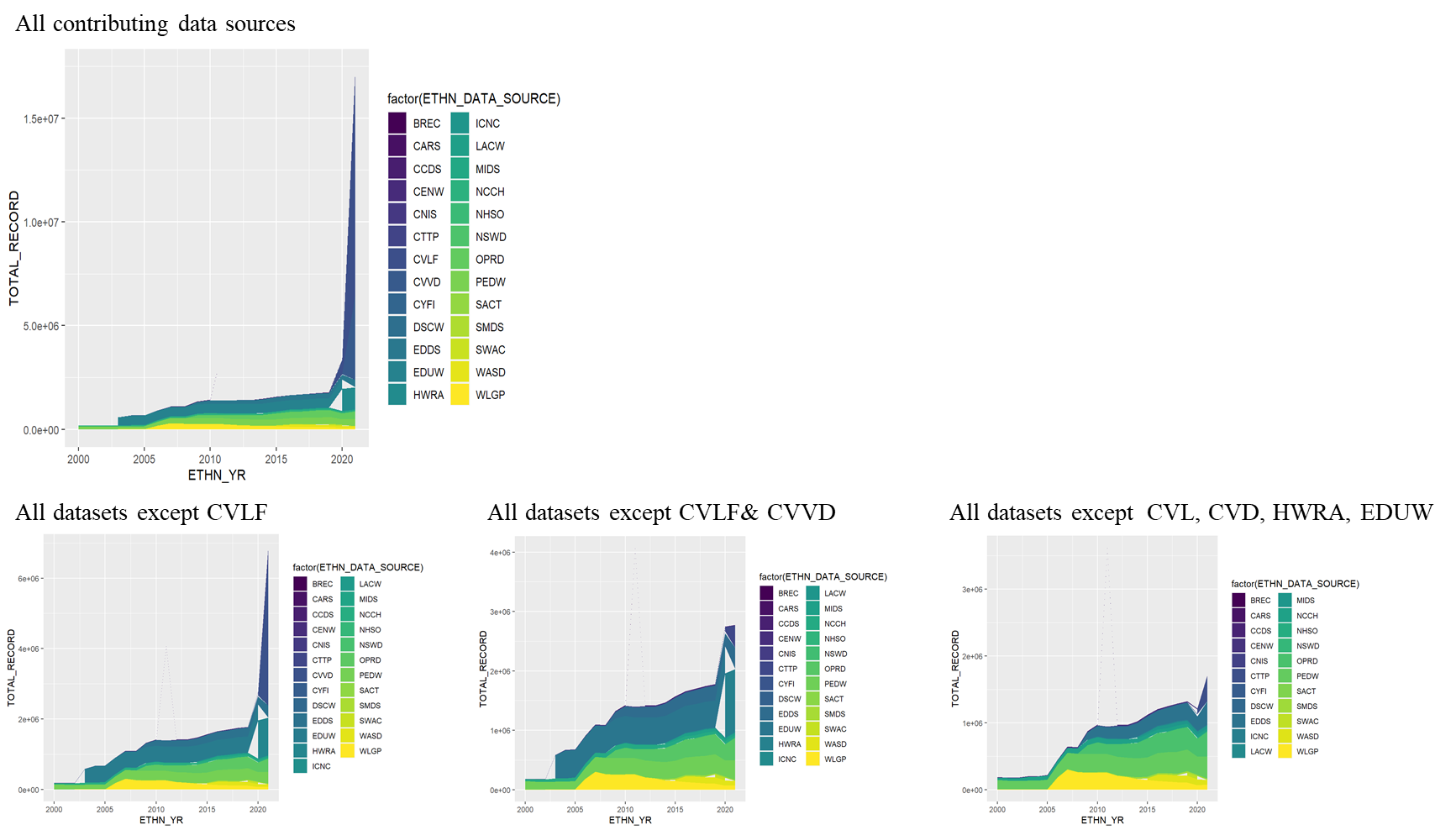

No prioritisation was applied, records from every data source have **equal value**.

In the following figures, we sequentially remove high-contributing data sources to drill down on the data sources contributing to the ethnic group over time.

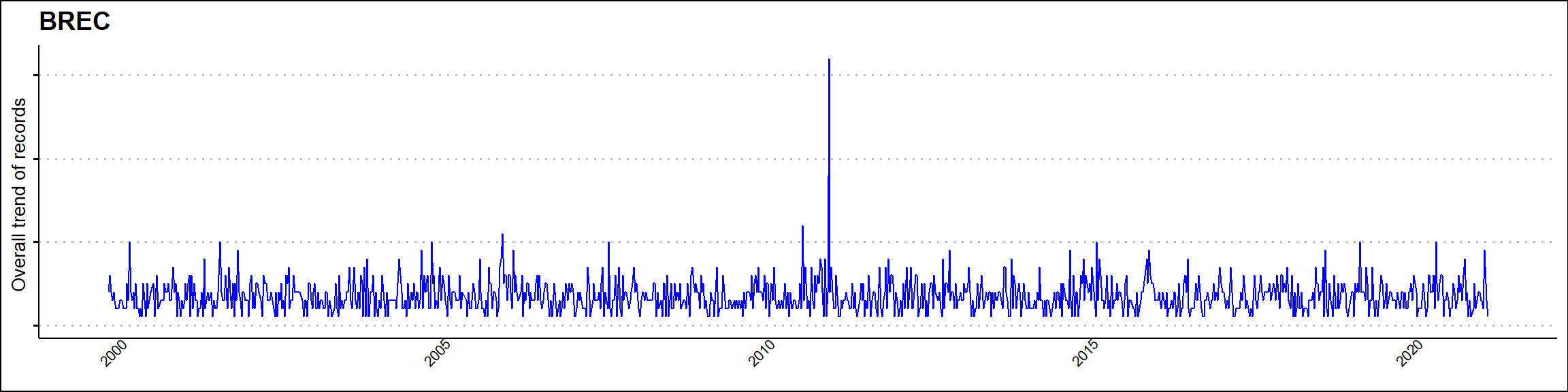

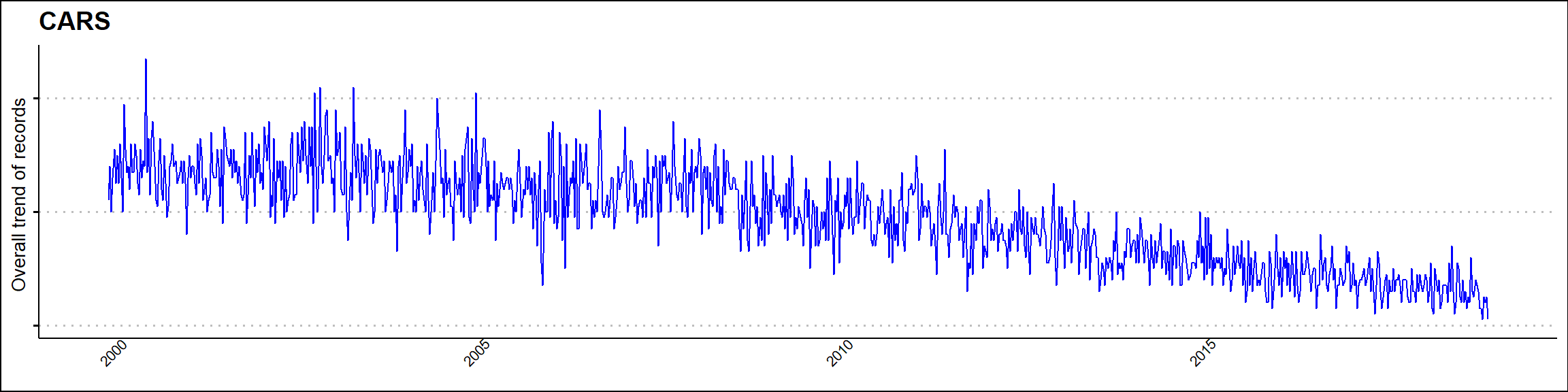

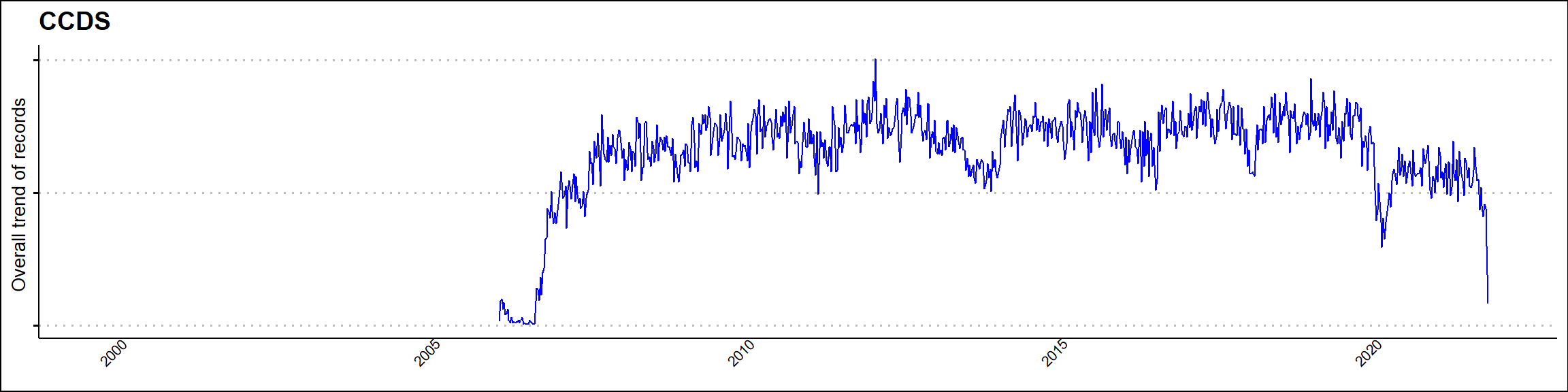

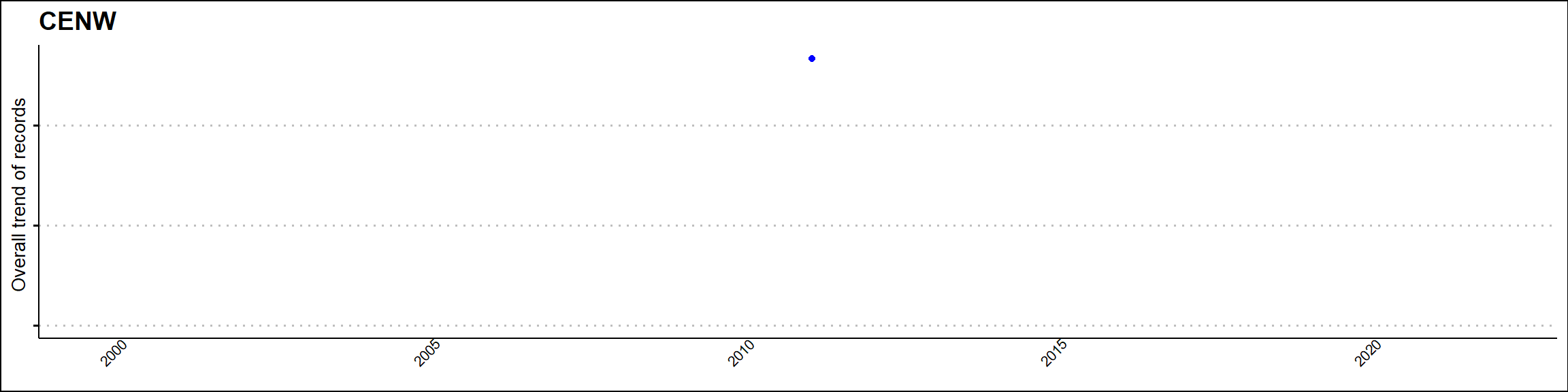

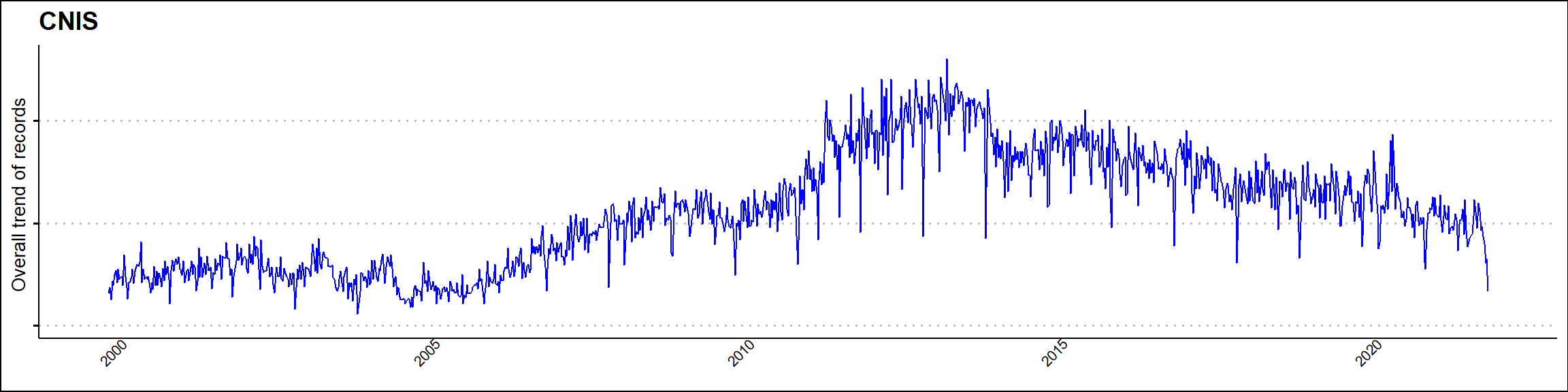

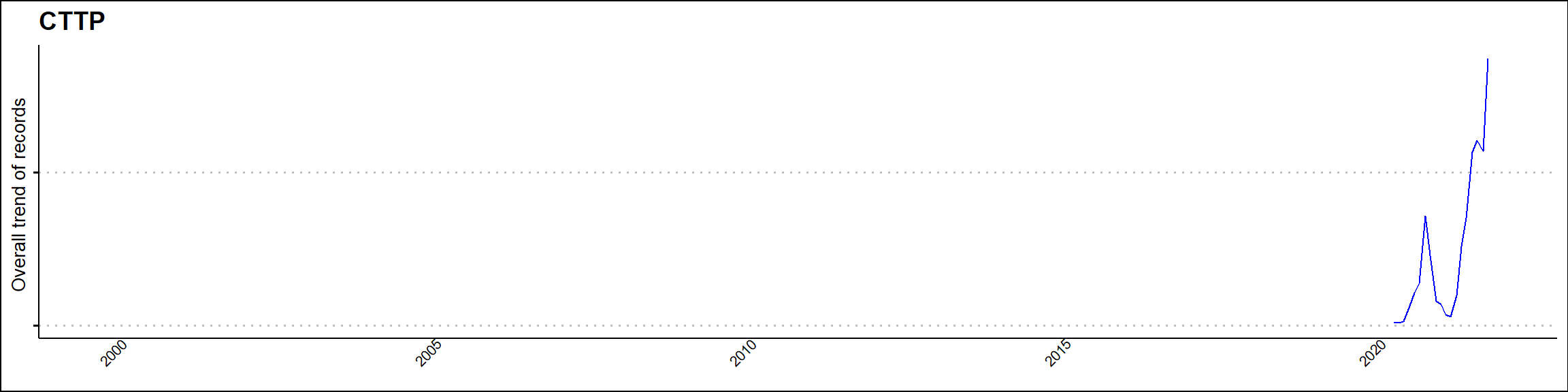

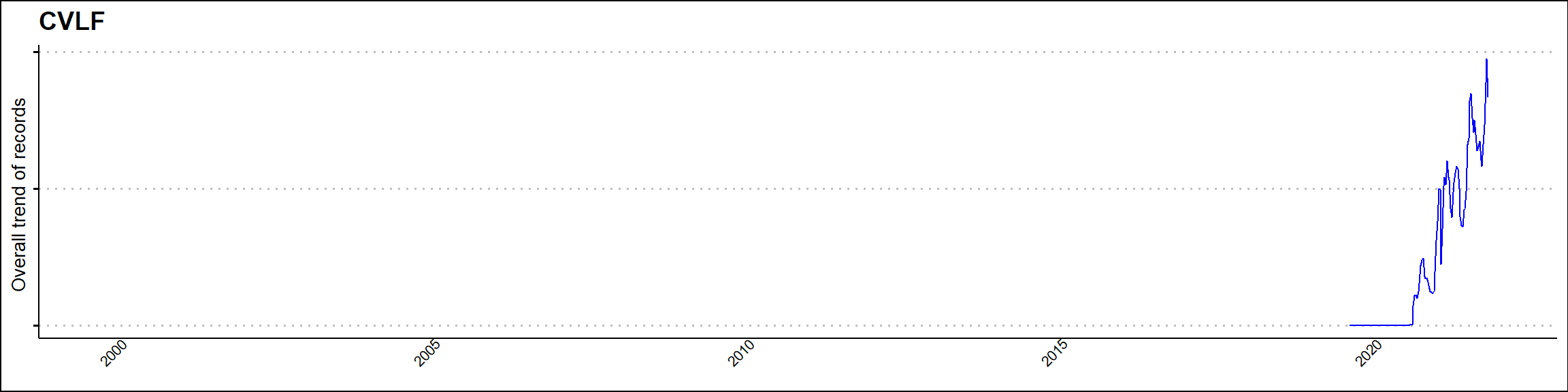

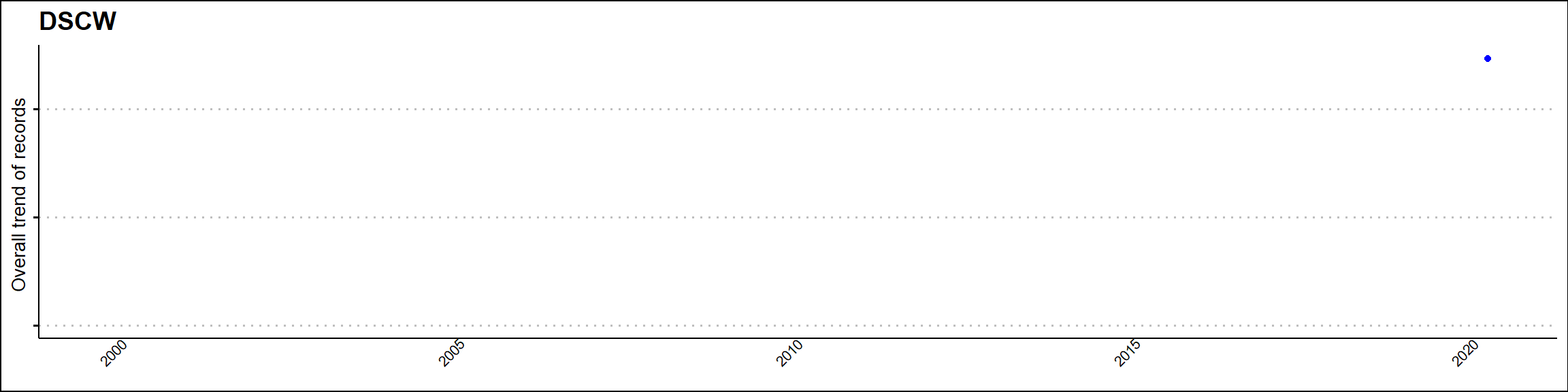

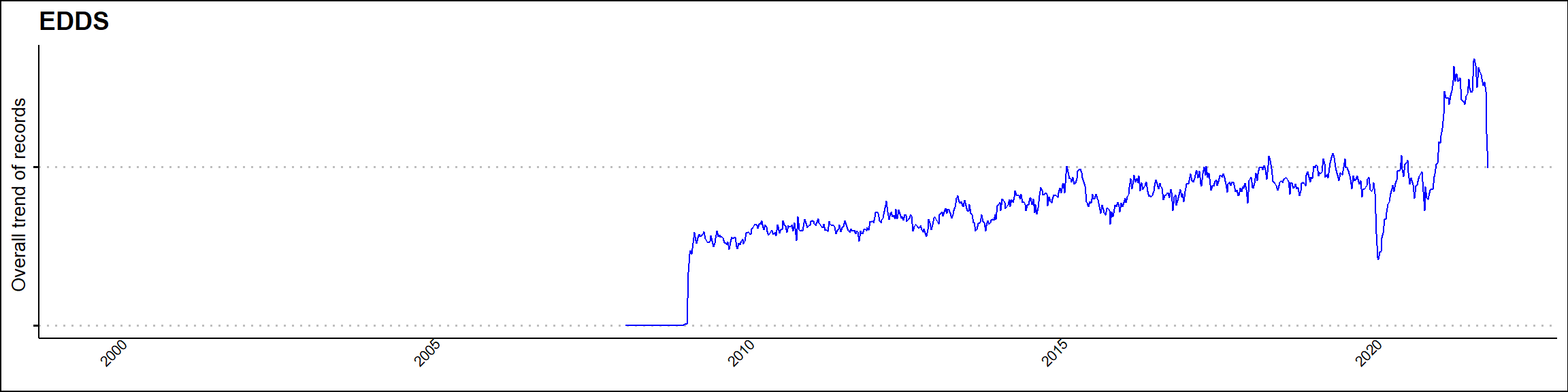

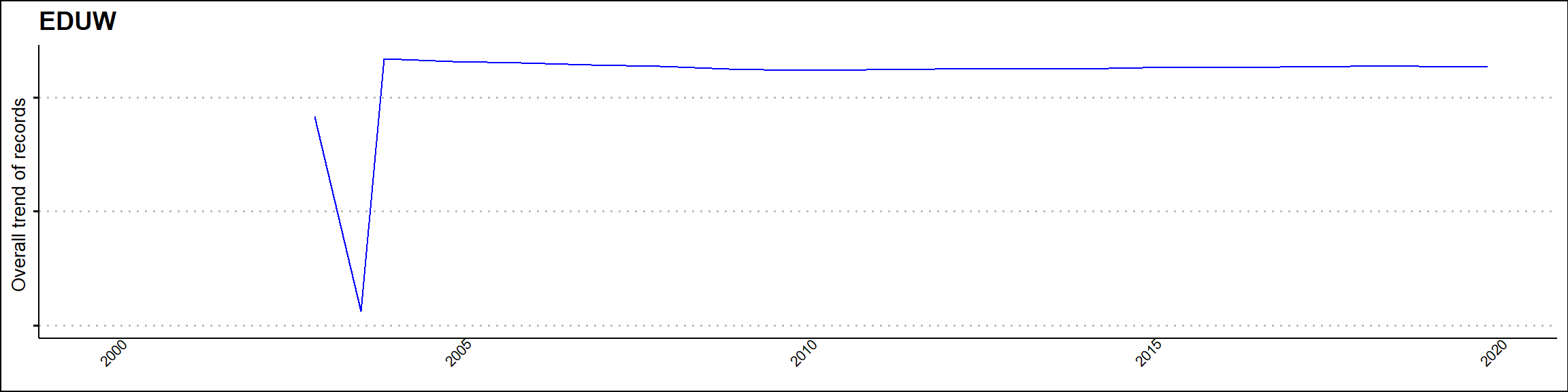

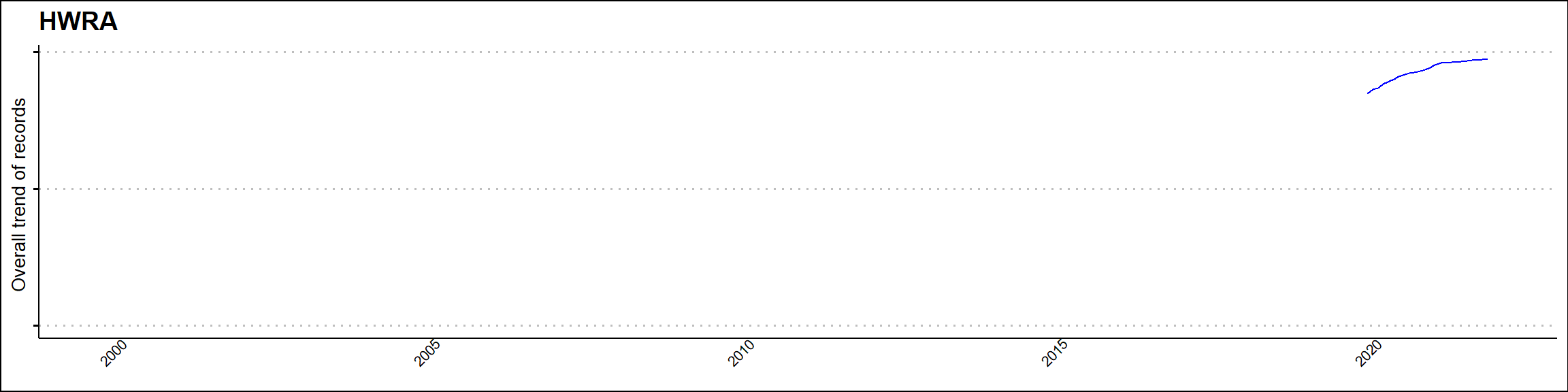

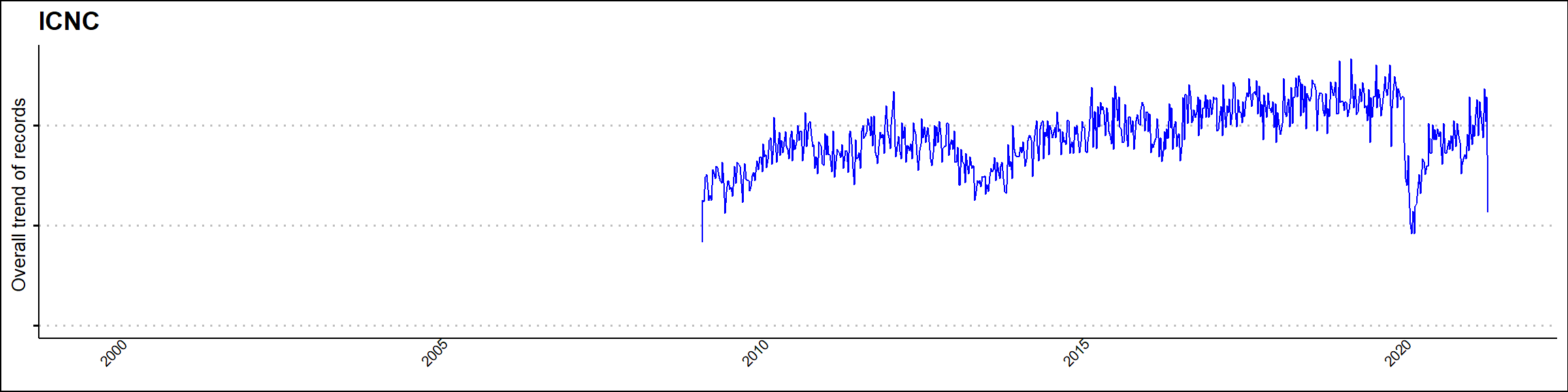

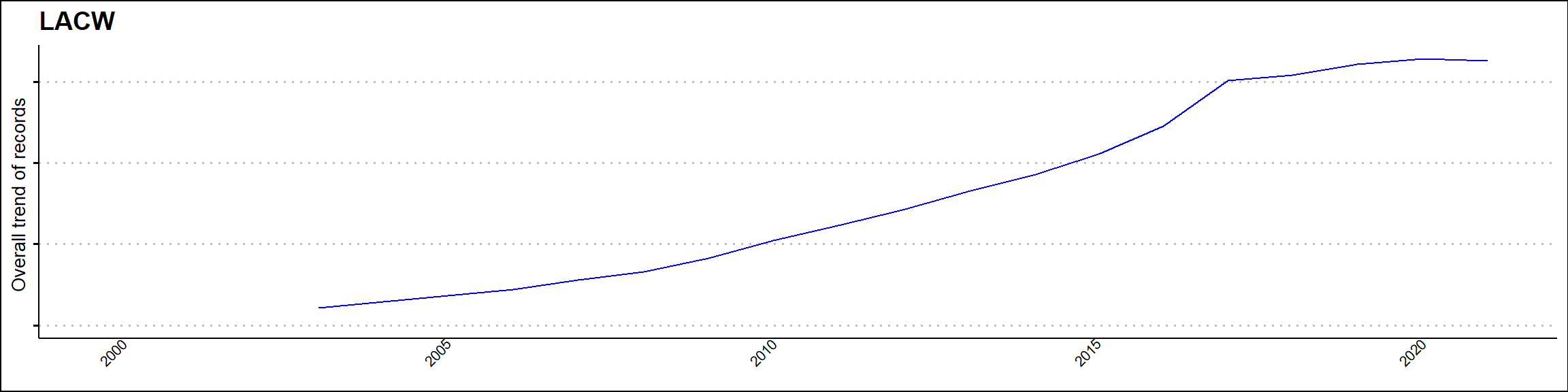

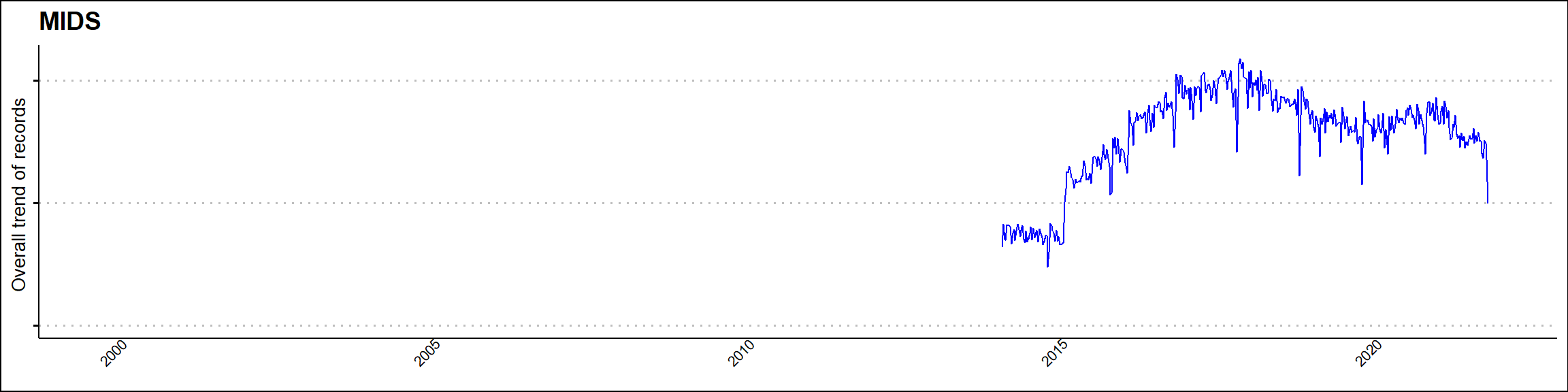

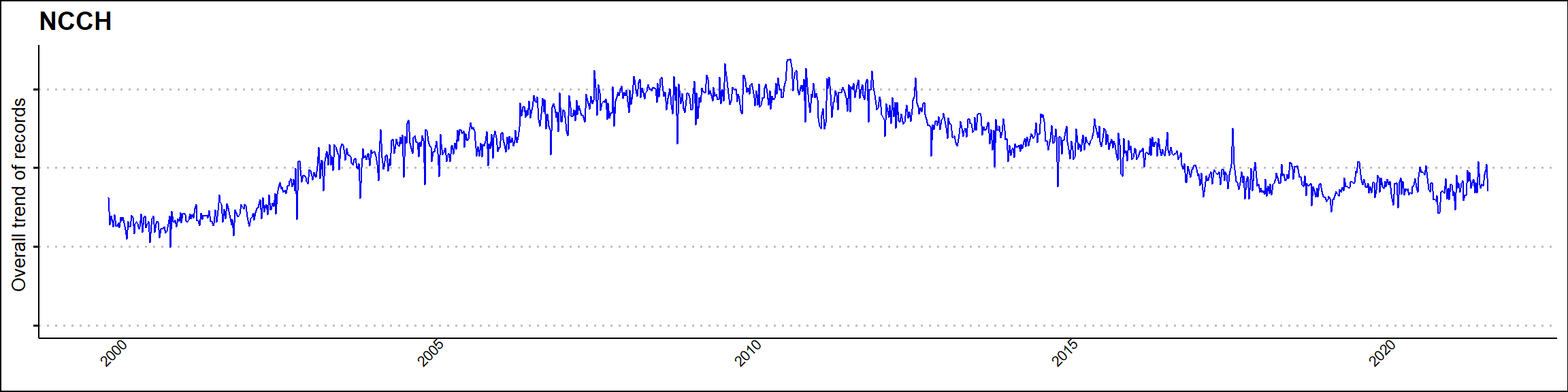

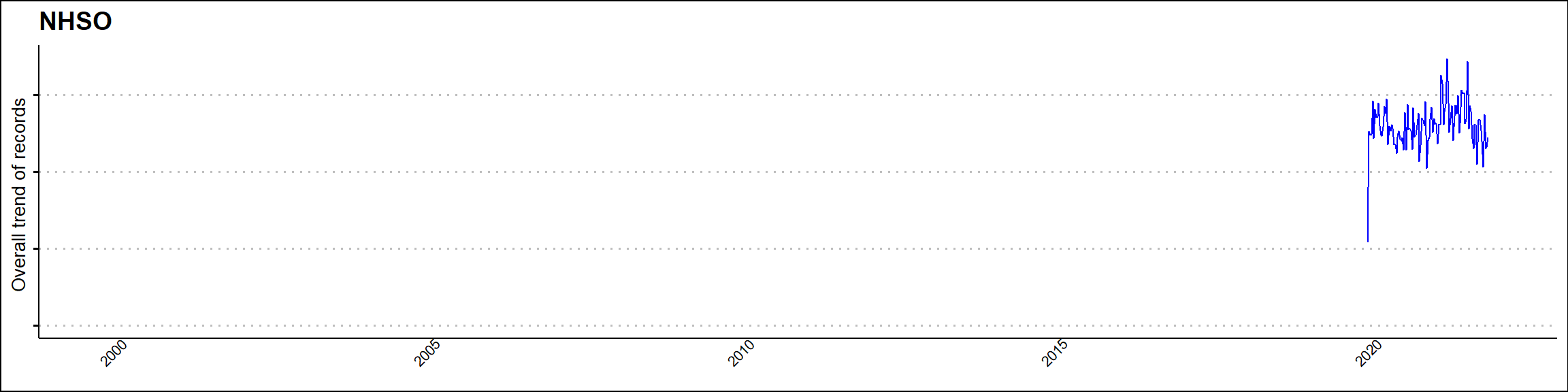

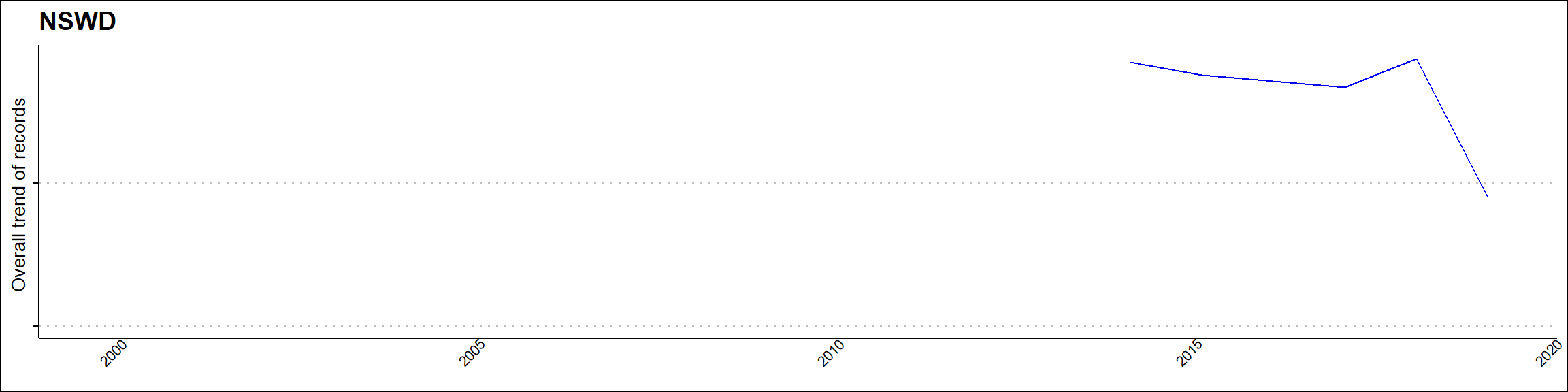

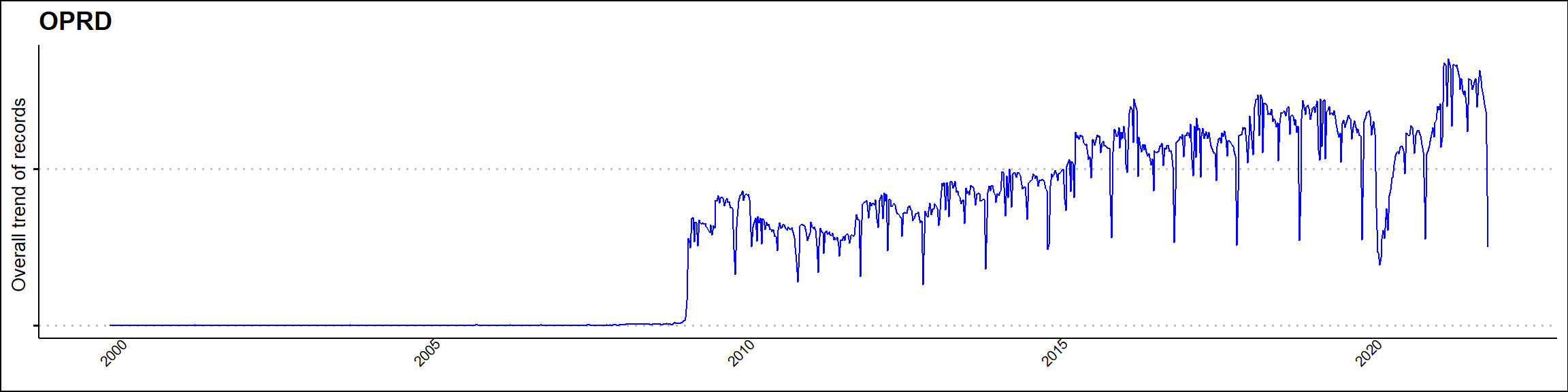

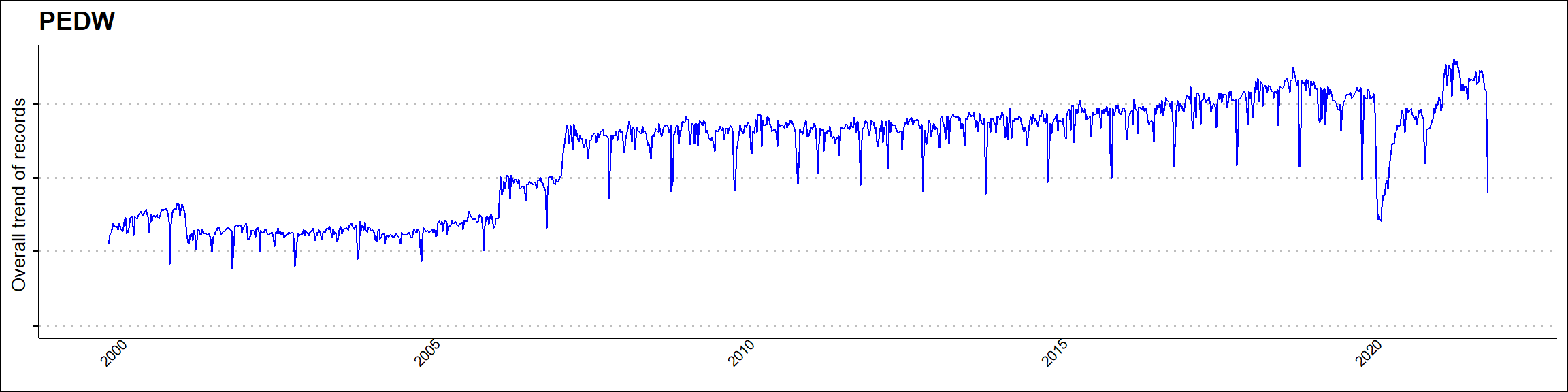

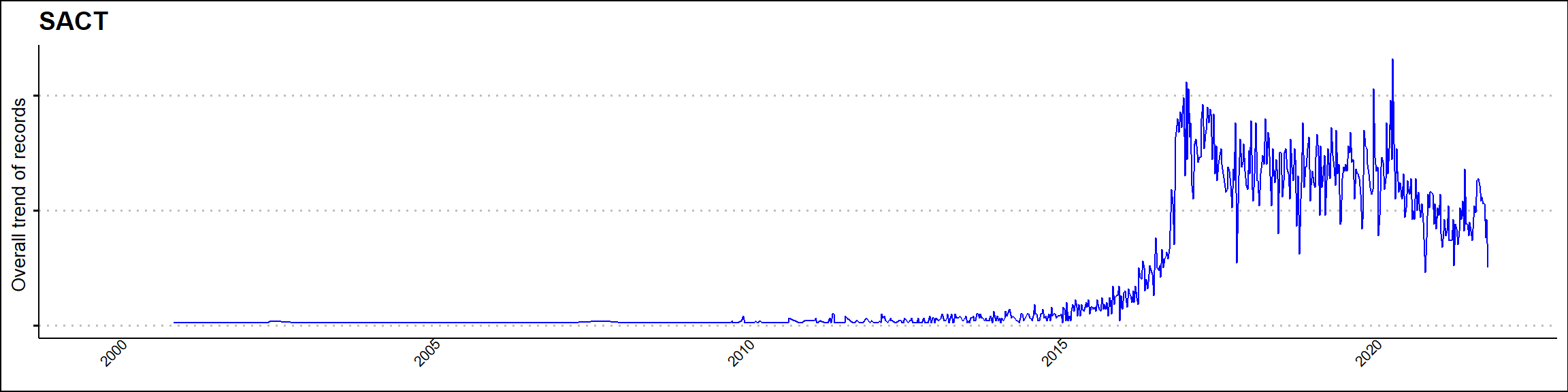

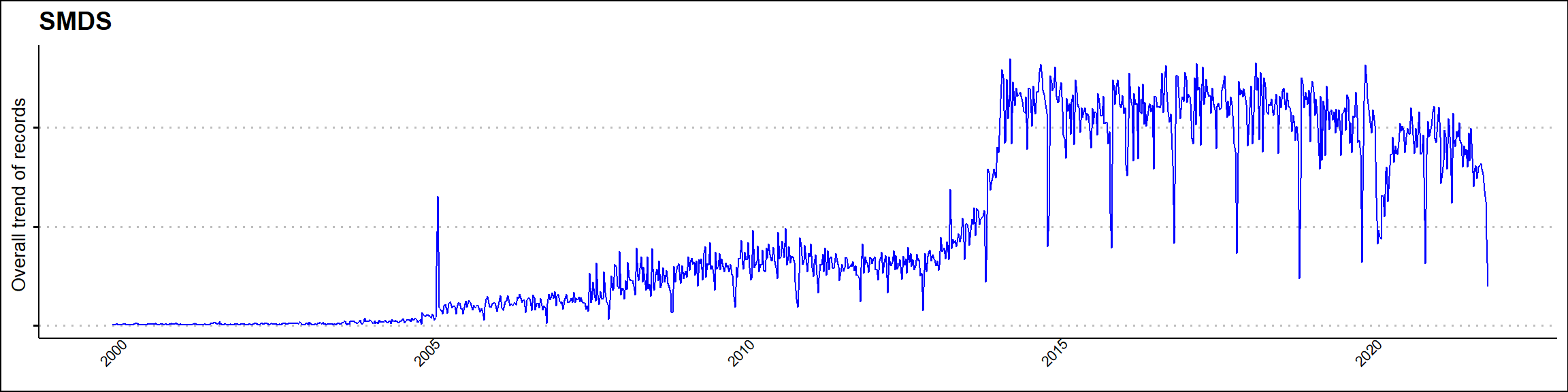

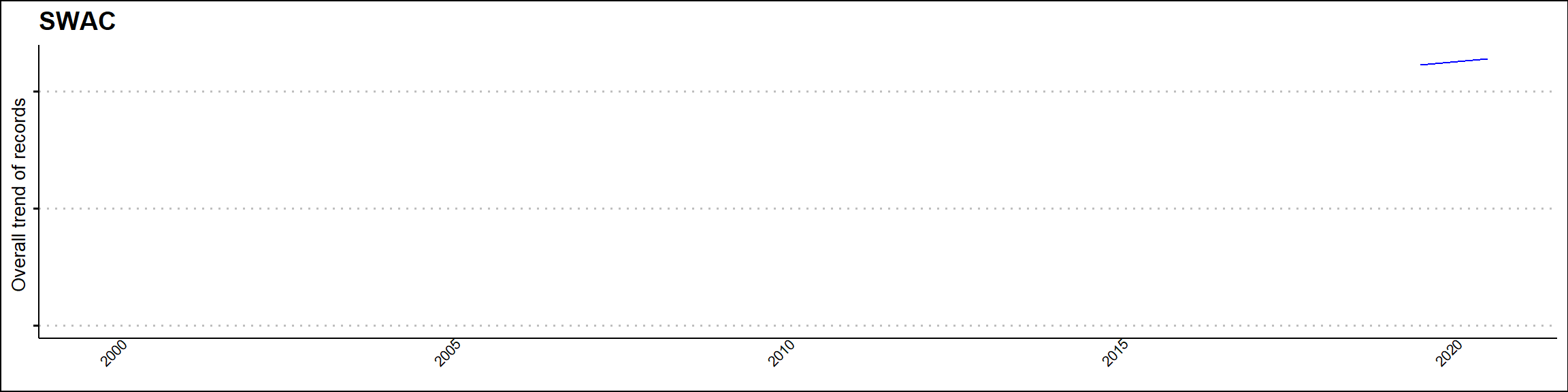

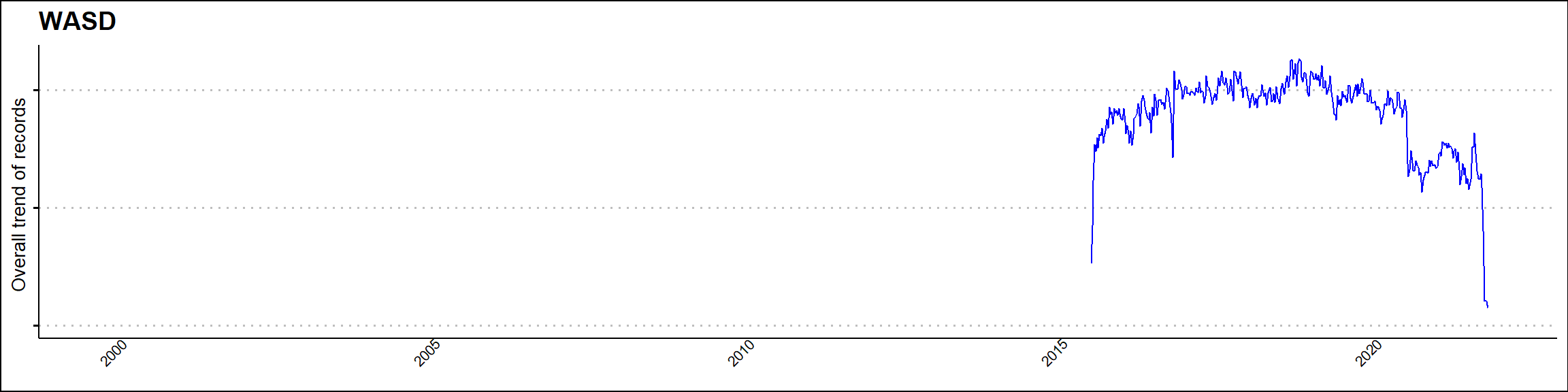

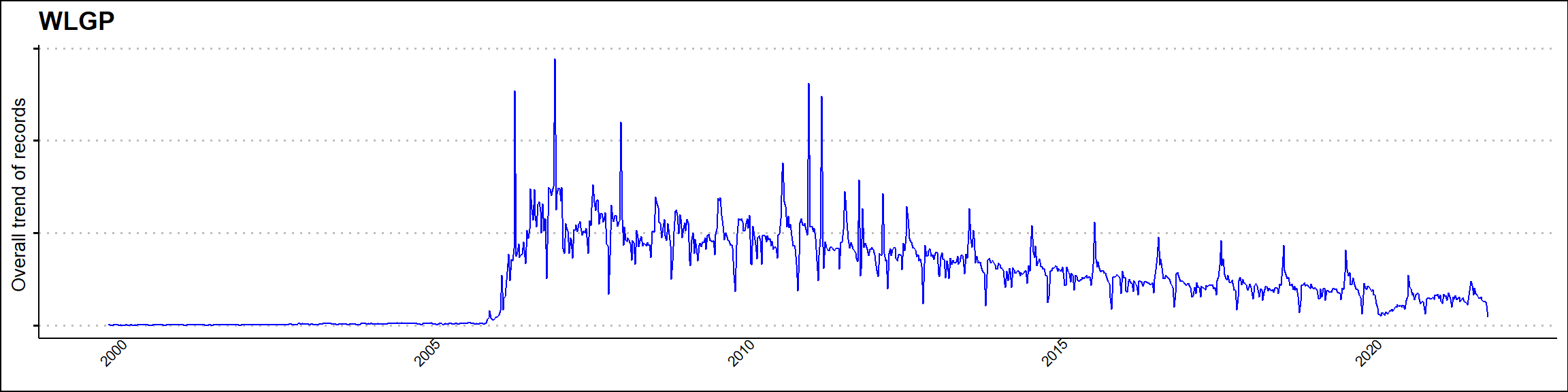

Supplementary Figure 3 – changes in population over time since 2011 (x-axis shows days since 2011-01-01)

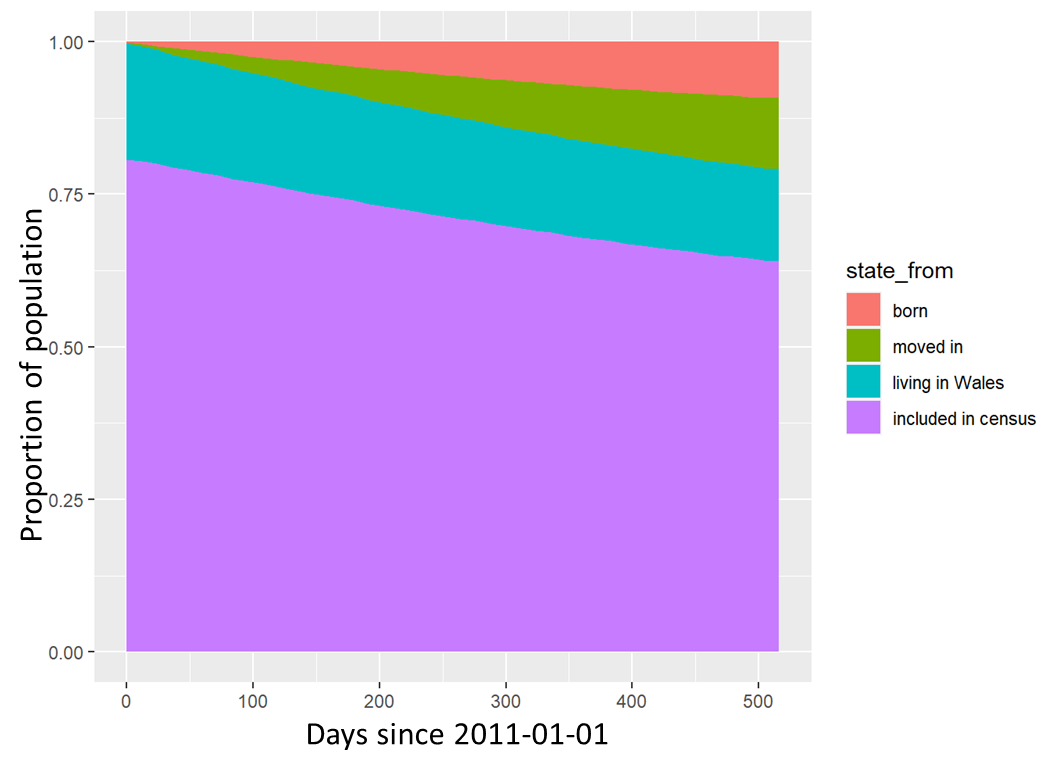

Supplementary Figure 4 – kappa index between all longitudinal records and Census data (Please note: The kappa of a dataset with itself has demonstrated to a less than 1.00 where individuals had contradicting records over time in a single dataset with itself)

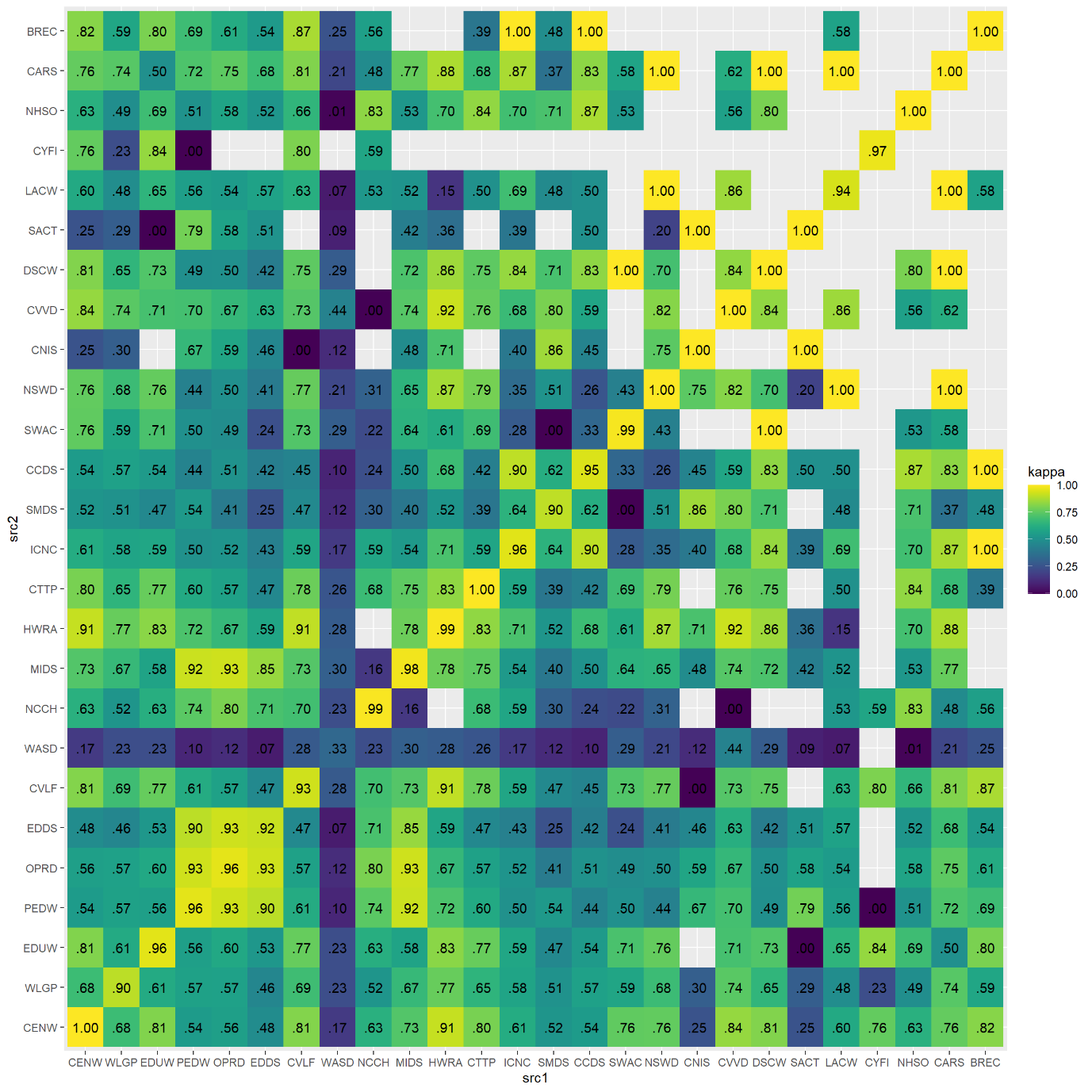

Supplementary Table 2A– characteristics of the cohort based on ONS categorisation.

| **Characteristics** | **Latest Ethnic group**  **N = 3,457,694**  **N (%)** | | **Mode Ethnic group**  **N = 3,457,694**  **N (%)** | | **Weighted Mod Ethnic group**  **N = 3,457,694**  **N (%)** | | **Composite Ethnic group**  **N = 3,457,694**  **N (%)** | |
| --- | --- | --- | --- | --- | --- | --- | --- | --- |
| **Age group** |  |  |  |  |  |  |  |  |
| 0-4 | 169,928 | (5.0%) | 169,928 | (5.0%) | 169,928 | (5.0%) | 169,928 | (5.0%) |
| 5-9 | 187,475 | (5.5%) | 187,475 | (5.5%) | 187,475 | (5.5%) | 187,475 | (5.5%) |
| 10-14 | 186,229 | (5.5%) | 186,229 | (5.5%) | 186,229 | (5.5%) | 186,229 | (5.5%) |
| 15-19 | 196,182 | (5.8%) | 196,182 | (5.8%) | 196,182 | (5.8%) | 196,182 | (5.8%) |
| 20-24 | 229,415 | (6.7%) | 229,415 | (6.7%) | 229,415 | (6.7%) | 229,415 | (6.7%) |
| 25-29 | 227,170 | (6.7%) | 227,170 | (6.7%) | 227,170 | (6.7%) | 227,170 | (6.7%) |
| 30-34 | 227,025 | (6.7%) | 227,025 | (6.7%) | 227,025 | (6.7%) | 227,025 | (6.7%) |
| 35-39 | 215,231 | (6.3%) | 215,231 | (6.3%) | 215,231 | (6.3%) | 215,231 | (6.3%) |
| 40-44 | 192,936 | (5.7%) | 192,936 | (5.7%) | 192,936 | (5.7%) | 192,936 | (5.7%) |
| 45-49 | 216,848 | (6.4%) | 216,848 | (6.4%) | 216,848 | (6.4%) | 216,848 | (6.4%) |
| 50-54 | 236,812 | (7.0%) | 236,812 | (7.0%) | 236,812 | (7.0%) | 236,812 | (7.0%) |
| 55-59 | 235,862 | (6.9%) | 235,862 | (6.9%) | 235,862 | (6.9%) | 235,862 | (6.9%) |
| 60-64 | 203,203 | (6.0%) | 203,203 | (6.0%) | 203,203 | (6.0%) | 203,203 | (6.0%) |
| 65-69 | 186,101 | (5.5%) | 186,101 | (5.5%) | 186,101 | (5.5%) | 186,101 | (5.5%) |
| 70-74 | 182,148 | (5.4%) | 182,148 | (5.4%) | 182,148 | (5.4%) | 182,148 | (5.4%) |
| 75-79 | 131,869 | (3.9%) | 131,869 | (3.9%) | 131,869 | (3.9%) | 131,869 | (3.9%) |
| 80-84 | 91,464 | (2.7%) | 91,464 | (2.7%) | 91,464 | (2.7%) | 91,464 | (2.7%) |
| 85+ | 86,322 | (2.5%) | 86,322 | (2.5%) | 86,322 | (2.5%) | 86,322 | (2.5%) |
| **Sex** |  |  |  |  |  |  |  |  |
| Male | 1,728,015 | (50.0%) | 1,728,015 | (50.0%) | 1,728,015 | (50.0%) | 1,728,015 | (50.0%) |
| Female | 1,729,679 | (50.0%) | 1,729,679 | (50.0%) | 1,729,679 | (50.0%) | 1,729,679 | (50.0%) |
| **WIMD 2019 quintiles** |  |  |  |  |  |  |  |  |
| 1-Most deprived | 658,913 | (21.0%) | 658,913 | (21.0%) | 658,913 | (21.0%) | 658,913 | (21.0%) |
| 2 | 638,925 | (20.0%) | 638,925 | (20.0%) | 638,925 | (20.0%) | 638,925 | (20.0%) |
| 3 | 643,624 | (20.0%) | 643,624 | (20.0%) | 643,624 | (20.0%) | 643,624 | (20.0%) |
| 4 | 630,614 | (20.0%) | 630,614 | (20.0%) | 630,614 | (20.0%) | 630,614 | (20.0%) |
| 5-Least deprived | 635,890 | (20.0%) | 635,890 | (20.0%) | 635,890 | (20.0%) | 635,890 | (20.0%) |
| **UHB of residence** |  |  |  |  |  |  |  |  |
| Aneurin Bevan | 605,782 | (19.0%) | 605,782 | (19.0%) | 605,782 | (19.0%) | 605,782 | (19.0%) |
| Betsi Cadwaladr | 701,543 | (22.0%) | 701,543 | (22.0%) | 701,543 | (22.0%) | 701,543 | (22.0%) |
| Cardiff and Vale | 526,652 | (16.0%) | 526,652 | (16.0%) | 526,652 | (16.0%) | 526,652 | (16.0%) |
| Cwm Taf Morgannwg | 459,118 | (14.0%) | 459,118 | (14.0%) | 459,118 | (14.0%) | 459,118 | (14.0%) |
| Hywel Dda | 385,174 | (12.0%) | 385,174 | (12.0%) | 385,174 | (12.0%) | 385,174 | (12.0%) |
| Powys | 128,461 | (4.0%) | 128,461 | (4.0%) | 128,461 | (4.0%) | 128,461 | (4.0%) |
| Swansea Bay | 401,236 | (13.0%) | 401,236 | (13.0%) | 401,236 | (13.0%) | 401,236 | (13.0%) |
| **Urban-rural category of residence** |  |  |  |  |  |  |  |  |
| Urban city and town | 2,179,684 | (68.0%) | 2,179,684 | (68.0%) | 2,179,684 | (68.0%) | 2,179,684 | (68.0%) |
| Urban city and town in a sparse setting | 59,602 | (1.9%) | 59,602 | (1.9%) | 59,602 | (1.9%) | 59,602 | (1.9%) |
| Rural town and fringe | 419,900 | (13.0%) | 419,900 | (13.0%) | 419,900 | (13.0%) | 419,900 | (13.0%) |
| Rural village and dispersed | 204,993 | (6.4%) | 204,993 | (6.4%) | 204,993 | (6.4%) | 204,993 | (6.4%) |
| Rural village and dispersed in a sparse setting | 225,307 | (7.0%) | 225,307 | (7.0%) | 225,307 | (7.0%) | 225,307 | (7.0%) |
| Rural town and fringe in a sparse setting | 118,480 | (3.7%) | 118,480 | (3.7%) | 118,480 | (3.7%) | 118,480 | (3.7%) |

Supplementary Table 2B – characteristics of the cohort based on NER categorisation.

| **Characteristics** | **Latest Ethnic group**  **N = 3,457,694**  N(%) | | **Mode Ethnic group**  **N = 3,457,694**  N(%) | | **Weighted Mode Ethnic group**  **N = 3,457,694**  N(%) | | **Composite Ethnic group**  **N = 3,457,694**  N(%) | |
| --- | --- | --- | --- | --- | --- | --- | --- | --- |
| **Age group** |  |  |  |  |  |  |  |  |
| 0-4 | 169,928 | (5.0%) | 169,928 | (5.0%) | 169,928 | (5.0%) | 169,928 | (5.0%) |
| 5-9 | 187,475 | (5.5%) | 187,475 | (5.5%) | 187,475 | (5.5%) | 187,475 | (5.5%) |
| 10-14 | 186,229 | (5.5%) | 186,229 | (5.5%) | 186,229 | (5.5%) | 186,229 | (5.5%) |
| 15-19 | 196,182 | (5.8%) | 196,182 | (5.8%) | 196,182 | (5.8%) | 196,182 | (5.8%) |
| 20-24 | 229,415 | (6.7%) | 229,415 | (6.7%) | 229,415 | (6.7%) | 229,415 | (6.7%) |
| 25-29 | 227,170 | (6.7%) | 227,170 | (6.7%) | 227,170 | (6.7%) | 227,170 | (6.7%) |
| 30-34 | 227,025 | (6.7%) | 227,025 | (6.7%) | 227,025 | (6.7%) | 227,025 | (6.7%) |
| 35-39 | 215,231 | (6.3%) | 215,231 | (6.3%) | 215,231 | (6.3%) | 215,231 | (6.3%) |
| 40-44 | 192,936 | (5.7%) | 192,936 | (5.7%) | 192,936 | (5.7%) | 192,936 | (5.7%) |
| 45-49 | 216,848 | (6.4%) | 216,848 | (6.4%) | 216,848 | (6.4%) | 216,848 | (6.4%) |
| 50-54 | 236,812 | (7.0%) | 236,812 | (7.0%) | 236,812 | (7.0%) | 236,812 | (7.0%) |
| 55-59 | 235,862 | (6.9%) | 235,862 | (6.9%) | 235,862 | (6.9%) | 235,862 | (6.9%) |
| 60-64 | 203,203 | (6.0%) | 203,203 | (6.0%) | 203,203 | (6.0%) | 203,203 | (6.0%) |
| 65-69 | 186,101 | (5.5%) | 186,101 | (5.5%) | 186,101 | (5.5%) | 186,101 | (5.5%) |
| 70-74 | 182,148 | (5.4%) | 182,148 | (5.4%) | 182,148 | (5.4%) | 182,148 | (5.4%) |
| 75-79 | 131,869 | (3.9%) | 131,869 | (3.9%) | 131,869 | (3.9%) | 131,869 | (3.9%) |
| 80-84 | 91,464 | (2.7%) | 91,464 | (2.7%) | 91,464 | (2.7%) | 91,464 | (2.7%) |
| 85+ | 86,322 | (2.5%) | 86,322 | (2.5%) | 86,322 | (2.5%) | 86,322 | (2.5%) |
| **Sex** |  |  |  |  |  |  |  |  |
| Male | 1,728,015 | (50.0%) | 1,728,015 | (50.0%) | 1,728,015 | (50.0%) | 1,728,015 | (50.0%) |
| Female | 1,729,679 | (50.0%) | 1,729,679 | (50.0%) | 1,729,679 | (50.0%) | 1,729,679 | (50.0%) |
| **WIMD 2019 quintiles** |  |  |  |  |  |  |  |  |
| 1-Most deprived | 658,913 | (21.0%) | 658,913 | (21.0%) | 658,913 | (21.0%) | 658,913 | (21.0%) |
| 2 | 638,925 | (20.0%) | 638,925 | (20.0%) | 638,925 | (20.0%) | 638,925 | (20.0%) |
| 3 | 643,624 | (20.0%) | 643,624 | (20.0%) | 643,624 | (20.0%) | 643,624 | (20.0%) |
| 4 | 630,614 | (20.0%) | 630,614 | (20.0%) | 630,614 | (20.0%) | 630,614 | (20.0%) |
| 5-Least deprived | 635,890 | (20.0%) | 635,890 | (20.0%) | 635,890 | (20.0%) | 635,890 | (20.0%) |
| **University Health Board (UHB)** |  |  |  |  |  |  |  |  |
| Aneurin Bevan | 605,782 | (19.0%) | 605,782 | (19.0%) | 605,782 | (19.0%) | 605,782 | (19.0%) |
| Betsi Cadwaladr | 701,543 | (22.0%) | 701,543 | (22.0%) | 701,543 | (22.0%) | 701,543 | (22.0%) |
| Cardiff and Vale | 526,652 | (16.0%) | 526,652 | (16.0%) | 526,652 | (16.0%) | 526,652 | (16.0%) |
| Cwm Taf Morgannwg | 459,118 | (14.0%) | 459,118 | (14.0%) | 459,118 | (14.0%) | 459,118 | (14.0%) |
| Hywel Dda | 385,174 | (12.0%) | 385,174 | (12.0%) | 385,174 | (12.0%) | 385,174 | (12.0%) |
| Powys | 128,461 | (4.0%) | 128,461 | (4.0%) | 128,461 | (4.0%) | 128,461 | (4.0%) |
| Swansea Bay | 401,236 | (13.0%) | 401,236 | (13.0%) | 401,236 | (13.0%) | 401,236 | (13.0%) |
| **Urban-rural category of residence** |  |  |  |  |  |  |  |  |
| Urban city and town | 2,179,684 | (68.0%) | 2,179,684 | (68.0%) | 2,179,684 | (68.0%) | 2,179,684 | (68.0%) |
| Urban city and town in a sparse setting | 59,602 | (1.9%) | 59,602 | (1.9%) | 59,602 | (1.9%) | 59,602 | (1.9%) |
| Rural town and fringe | 419,900 | (13.0%) | 419,900 | (13.0%) | 419,900 | (13.0%) | 419,900 | (13.0%) |
| Rural village and dispersed in a sparse setting | 225,307 | (7.0%) | 225,307 | (7.0%) | 225,307 | (7.0%) | 225,307 | (7.0%) |
| Rural village and dispersed | 204,993 | (6.4%) | 204,993 | (6.4%) | 204,993 | (6.4%) | 204,993 | (6.4%) |
| Rural town and fringe in a sparse setting | 118,480 | (3.7%) | 118,480 | (3.7%) | 118,480 | (3.7%) | 118,480 | (3.7%) |

Supplementary Table 3– meta-data of existing ethnicity RRDA tables

| # | Column name | Type | Description |
| --- | --- | --- | --- |
| 01 | ALF_E | BIGINT | Anonymised Linkage Field (ALF) |
| 02 | ALF_E_C16 | SMALLINT | ALF flag if in C19_COHORT16 (0 = NO / 1 = YES) |
| 03 | ALF_E_C20 | SMALLINT | ALF flag if in C19_COHORT20 (0 = NO / 1 = YES) |
| 04 | ALF_E_C16_C20 | SMALLINT | ALF flag if in C19_COHORT16 or C19_COHORT20 (0 = NO / 1 = YES) |
| 05 | ETHN_DATE_LATEST | DATE | Latest date method - Date of latest ethnicity recording |
| 06 | ETHN_EC_ONS_DATE_LATEST_CODE | SMALLINT | Latest date method code - Harmonised aggregate ethnicity - ONS 5 categories |
| 07 | ETHN_EC_ONS_DATE_LATEST_DESC | VARCHAR(15) | Latest date method description - Harmonised aggregate ethnicity - ONS 5 categories |
| 08 | ETHN_EC_NER_DATE_LATEST_CODE | SMALLINT | Latest date method code - Harmonised aggregate ethnicity - NER 9 categories |
| 08 | ETHN_EC_NER_DATE_LATEST_DESC | VARCHAR(15) | Latest date method description - Harmonised aggregate ethnicity - NER 9 categories |
| 09 | ETHN_EC_MIN_DATE_LATEST_CODE | SMALLINT | Latest date method code - Harmonised aggregate ethnicity - MIN 5 categories |
| 10 | ETHN_EC_MIN_DATE_LATEST_DESC | VARCHAR(15) | Latest date method description - Harmonised aggregate ethnicity - MIN 5 categories |
| 11 | ETHN_DATE_COUNTS | INTEGER | Number of ethnicity recordings in all data sources |
| 12 | ETHN_EC_ONS_MODE_CODE | SMALLINT | Mode (most common) method code - Harmonised aggregate ethnicity - ONS 5 categories |
| 13 | ETHN_EC_ONS_MODE_DESC | VARCHAR(15) | Mode (most common) method description - Harmonised aggregate ethnicity - ONS 5 categories |
| 14 | ETHN_EC_NER_MODE_CODE | SMALLINT | Mode (most common) method code - Harmonised aggregate ethnicity - NER 9 categories |
| 15 | ETHN_EC_NER_MODE_DESC | VARCHAR(15) | Mode (most common) method description - Harmonised aggregate ethnicity - NER 9 categories |
| 16 | ETHN_EC_MIN_MODE_CODE | SMALLINT | Mode (most common) method code - Harmonised aggregate ethnicity - MIN 5 categories |
| 17 | ETHN_EC_MIN_MODE_DESC | VARCHAR(15) | Mode (most common) method description - Harmonised aggregate ethnicity - MIN 5 categories |
| 18 | ETHN_EC_ONS_WMOD_CODE | SMALLINT | Weighted mode method code - Harmonised aggregate ethnicity - ONS 5 categories |
| 19 | ETHN_EC_ONS_WMOD_DESC | VARCHAR(15) | Weighted mode method description - Harmonised aggregate ethnicity - ONS 5 categories |
| 20 | ETHN_EC_NER_WMOD_CODE | SMALLINT | Weighted mode method code - Harmonised aggregate ethnicity - NER 9 categories |
| 21 | ETHN_EC_NER_WMOD_DESC | VARCHAR(15) | Weighted mode method description - Harmonised aggregate ethnicity - NER 9 categories |
| 22 | ETHN_EC_MIN_WMOD_CODE | SMALLINT | Weighted mode method code - Harmonised aggregate ethnicity - MIN 5 categories |
| 23 | ETHN_EC_MIN_WMOD_DESC | VARCHAR(15) | Weighted mode method description - Harmonised aggregate ethnicity - MIN 5 categories |
| 24 | ETHN_EC_ONS_COMP_LENGTH | INTEGER | Composite method - number of harmonised categories recorded - ONS 5 categories |
| 25 | ETHN_EC_ONS_COMP_ORIG_CODE | INTEGER | Composite method code - duplicate categories remain as duplicates - ONS 5 categories |
| 26 | ETHN_EC_ONS_COMP_ORIG_DESC | INTEGER | Composite method description - duplicate categories remain as duplicates - ONS 5 categories |
| 27 | ETHN_EC_ONS_COMP_MIXD_CODE | INTEGER | Composite method code - duplicate categories are assigned as MIXED - ONS 5 categories |
| 28 | ETHN_EC_ONS_COMP_MIXD_DESC | INTEGER | Composite method description - duplicate categories are assigned as MIXED - ONS 5 categories |
| 29 | ETHN_EC_ONS_COMP_NULL_CODE | INTEGER | Composite method code - duplicate categories are assigned as NULL - ONS 5 categories |
| 30 | ETHN_EC_ONS_COMP_NULL_DESC | INTEGER | Composite method description - duplicate categories are assigned as NULL - ONS 5 categories |
| 31 | ETHN_EC_NER_COMP_LENGTH | INTEGER | Composite method - number of harmonised categories recorded - NER 9 categories |
| 32 | ETHN_EC_NER_COMP_ORIG_CODE | INTEGER | Composite method code - duplicate categories remain as duplicates - NER 9 categories |
| 33 | ETHN_EC_NER_COMP_ORIG_DESC | INTEGER | Composite method description - duplicate categories remain as duplicates - NER 9 categories |
| 34 | ETHN_EC_NER_COMP_MIXD_CODE | INTEGER | Composite method code - duplicate categories are assigned as MIXED - NER 9 categories |
| 35 | ETHN_EC_NER_COMP_MIXD_DESC | INTEGER | Composite method description - duplicate categories are assigned as MIXED - NER 9 categories |
| 36 | ETHN_EC_NER_COMP_NULL_CODE | INTEGER | Composite method code - duplicate categories are assigned as NULL - NER 9 categories |
| 37 | ETHN_EC_NER_COMP_NULL_DESC | INTEGER | Composite method description - duplicate categories are assigned as NULL - NER 9 categories |
| 38 | ETHN_EC_MIN_COMP_LENGTH | INTEGER | Composite method - number of harmonised categories recorded - MIN 5 categories |
| 32 | ETHN_EC_MIN_COMP_ORIG_CODE | INTEGER | Composite method code - duplicate categories remain as duplicates -MIN 5 categories |
| 33 | ETHN_EC_MIN_COMP_ORIG_DESC | INTEGER | Composite method description - duplicate categories remain as duplicates - MIN 5 categories |
| 34 | ETHN_EC_MIN_COMP_MIXD_CODE | INTEGER | Composite method code - duplicate categories are assigned as MIXED - MIN 5 categories |
| 35 | ETHN_EC_MIN_COMP_MIXD_DESC | INTEGER | Composite method description - duplicate categories are assigned as MIXED - MIN 5 categories |
| 36 | ETHN_EC_MIN_COMP_NULL_CODE | INTEGER | Composite method code - duplicate categories are assigned as NULL - MIN 5 categories |
| 37 | ETHN_EC_MIN_COMP_NULL_DESC | INTEGER | Composite method description - duplicate categories are assigned as NULL - MIN 5 categories |

**RRDA_ETHN_PREP_DATE**

| # | Column name | Type | Description |
| --- | --- | --- | --- |
| 01 | ALF_E | BIGINT | Anonymised Linkage Field (ALF) |
| 02 | ALF_E_C16 | SMALLINT | ALF flag if in C19_COHORT16 (0 = NO / 1 = YES) |
| 03 | ALF_E_C20 | SMALLINT | ALF flag if in C19_COHORT20 (0 = NO / 1 = YES) |
| 04 | ALF_E_C16_C20 | SMALLINT | ALF flag if in C19_COHORT16 or C19_COHORT20 (0 = NO / 1 = YES) |
| 05 | ETHN_DATA_SOURCE | CHAR(4) | Data source the ethnicity recording is from |
| 06 | ETHN_DATE | DATE | Date of ethnicity recording |
| 07 | ETHN_EC_ONS_CODE | SMALLINT | Harmonised aggregate ethnicity code - ONS 5 categories |
| 08 | ETHN_EC_ONS_DESC | VARCHAR(15) | Harmonised aggregate ethnicity description - ONS 5 categories |
| 09 | ETHN_EC_ONS_ORDER | SMALLINT | Ordered list (newest num 1) of all ethnicity recorded dates |
| 10 | ETHN_EC_NER_CODE | SMALLINT | Harmonised aggregate ethnicity code - NER 9 categories |
| 11 | ETHN_EC_NER_DESC | VARCHAR(15) | Harmonised aggregate ethnicity description - NER 9 categories |
| 12 | ETHN_EC_NER_ORDER | SMALLINT | Ordered list (newest num 1) of all ethnicity recorded dates |
| 13 | ETHN_EC_MIN_CODE | SMALLINT | Harmonised aggregate ethnicity code - MIN 5 categories |
| 14 | ETHN_EC_MIN_DESC | VARCHAR(15) | Harmonised aggregate ethnicity description - MIN 5 categories |
| 15 | ETHN_EC_MIN_ORDER | SMALLINT | Ordered list (newest num 1) of all ethnicity recorded dates |
| 16 | ETHN_DATE_LATEST | DATE | Latest date method - Date of latest ethnicity recording |
| 17 | ETHN_EC_ONS_DATE_LATEST_CODE | SMALLINT | Latest date method code - Harmonised aggregate ethnicity - ONS 5 categories |
| 18 | ETHN_EC_ONS_DATE_LATEST_DESC | VARCHAR(15) | Latest date method description - Harmonised aggregate ethnicity - ONS 5 categories |
| 19 | ETHN_EC_NER_DATE_LATEST_CODE | SMALLINT | Latest date method code - Harmonised aggregate ethnicity - NER 9 categories |
| 20 | ETHN_EC_NER_DATE_LATEST_DESC | VARCHAR(15) | Latest date method description - Harmonised aggregate ethnicity - NER 9 categories |
| 21 | ETHN_EC_MIN_DATE_LATEST_CODE | SMALLINT | Latest date method code - Harmonised aggregate ethnicity - MIN 5 categories |
| 22 | ETHN_EC_MIN_DATE_LATEST_DESC | VARCHAR(15) | Latest date method description - Harmonised aggregate ethnicity - MIN 5 categories |
| 23 | ETHN_DATE_COUNTS | INTEGER | Number of ethnicity recordings in all data sources |

**RRDA_ETHN_EHRD / RRDA_ETHN_CENW / RRDA_ETHN_CHHV**

The structure of these tables is the same as RRDA_ETHN - but the "ETHN" in the field name is replaced by "EHRD" / "CENW" / "CHHV" respectively

**RRDA_ETHN_PREP_EHRD_DATE / RRDA_ETHN_PREP_CENW_DATE / RRDA_ETHN_PREP_CHHV_DATE**

The structure of these tables is the same as RRDA_ETHN_PREP_DATE - but the "ETHN" in the field name is replaced by "EHRD" / "CENW" / "CHHV" respectively

**Supplementary 4– Ethnic group categorisation and harmonisation rules**.

[https://portal.caliberresearch.org/phenotypes/ethnic-statu](https://portal.caliberresearch.org/phenotypes/ethnic-status)s

### BREC (Brecon data)

| ONS category | Data source code | Data source description |
| --- | --- | --- |
| 1 White |  | 'any other white background'  'british'  'irish' |
| 2 Mixed |  | ‘any other mixed background'  ‘white and asian'  'white and black african'  'white and black caribbean’ |
| 3 Asian |  | 'any other asian background'  'asian'  'bangladeshi'  ’chinese'  'indian'  'pakistani' |
| 4 Black |  | 'african'  'any other black background'  ’black african'  'caribbean' |
| 5 Other |  | ‘any other ethnic group’ |

| NER category | Data source code | Data source description |
| --- | --- | --- |
| 1 White |  | 'any other white background'  'british'  'irish' |
| 2 Mixed |  | ‘any other mixed background'  ‘white and asian'  'white and black african'  'white and black caribbean’ |
| 3 Indian |  | 'indian' |
| 4 Pakistani |  | 'pakistani' |
| 5 Bangladeshi |  | 'bangladeshi' |
| 6 Chinese |  | ’chinese' |
| 7 Black Caribbean |  | 'caribbean' |
| 8 Black African |  | 'african'  ’black african' |
| 9 Other |  | 'any other asian background'  'any other black background'  ‘any other ethnic group’  'asian' |

### CARS (Congenital Anomaly Register and Information Services for Wales)

| ONS category | Data source code | Data source description |
| --- | --- | --- |
| 1 White | 1  2  3 | White British  White Irish  White any other |
| 2 Mixed | 4  5  6  7 | Mixed W& B Car  Mixed W & B Afr  Mixed W & Asian  Mixed any other |
| 3 Asian | 8  10  11  12  18 | Indian  Pakistani  Bangladeshi  Asian, other  Other, Chinese |
| 4 Black | 13  14  16 | Black Caribbean  Black African  Black, other |
| 5 Other | 19 | Other, any other |

| NER category | Data source code | Data source description |
| --- | --- | --- |
| 1 White | 1  2  3 | White British  White Irish  White any other |
| 2 Mixed | 4  5  6  7 | Mixed W& B Car  Mixed W & B Afr  Mixed W & Asian  Mixed any other |
| 3 Indian | 8 | Indian |
| 4 Pakistani | 10 | Pakistani |
| 5 Bangladeshi | 11 | Bangladeshi |
| 6 Chinese | 18 | Other, Chinese |
| 7 Black Caribbean | 13 | Black Caribbean |
| 8 Black African | 14 | Black African |
| 9 Other | 16  19 | Asian, other  Black, other |

### CCDS (Critical Care DataSet)

| ONS category | Data source code | Data source description |
| --- | --- | --- |
| 1 White | A  B | Any White Background, including Welsh, English, Scottish, Northern Irish, Irish, British  Gypsy or Irish Traveller |
| 2 Mixed | D  E  F  G | White and Black Caribbean  White and Black African  White and Asian  Any other mixed background / multiple ethnic background |
| 3 Asian | H  J  K  L  R | Indian  Pakistani  Bangladeshi  Any other Asian Background  Chinese |
| 4 Black | M  N  P | Caribbean  African  Any other Black background |
| 5 Other | S  T | Any other ethnic group  Arab |

| NER category | Data source code | Data source description |
| --- | --- | --- |
| 1 White | A  B | Any White Background, including Welsh, English, Scottish, Northern Irish, Irish, British  Gypsy or Irish Traveller |
| 2 Mixed | D  E  F  G | White and Black Caribbean  White and Black African  White and Asian  Any other mixed background / multiple ethnic background |
| 3 Indian | H | Indian |
| 4 Pakistani | J | Pakistani |
| 5 Bangladeshi | K | Bangladeshi |
| 6 Chinese | R | Chinese |
| 7 Black Caribbean | M | Caribbean |
| 8 Black African | N | African |
| 9 Other | L  P  S  T | Any other Asian Background  Any other Black background  Any other ethnic group  Arab |

### CENW (ONS Census 2011)

| ONS category | Data source code | Data source description |
| --- | --- | --- |
| 1 White | 01  02  03  04 | English / Welsh / Scottish / Northern Irish / British  Irish  Gypsy or Irish Traveller  Other White |
| 2 Mixed | 05  06  07  08 | White and Black Caribbean  White and Black African  White and Asian  Other Mixed |
| 3 Asian | 09  10  11  12  13 | Indian  Pakistani  Bangladeshi  Chinese  Other Asian |
| 4 Black | 14  15  16 | African  Caribbean  Other Black |
| 5 Other | 17  18 | Arab  Other Ethnic Group |

| NER category | Data source code | Data source description |
| --- | --- | --- |
| 1 White | 01  02  03  04 | English / Welsh / Scottish / Northern Irish / British  Irish  Gypsy or Irish Traveller  Other White |
| 2 Mixed | 05  06  07  08 | White and Black Caribbean  White and Black African  White and Asian  Other Mixed |
| 3 Indian | 09 | Indian |
| 4 Pakistani | 10 | Pakistani |
| 5 Bangladeshi | 11 | Bangladeshi |
| 6 Chinese | 12 | Chinese |
| 7 Black Caribbean | 14 | African |
| 8 Black African | 15 | Caribbean |
| 9 Other | 13  16  17  18 | Other Asian  Other Black  Arab  Other Ethnic Group |

### CNIS (CANISC – Cancer Network Information System Cymru)

| ONS category | Data source code | Data source description |
| --- | --- | --- |
| 1 White | A  B | Any White Background, including Welsh, English, Scottish, Northern Irish, Irish, British  Gypsy or Irish Traveller |
| 2 Mixed | D  E  F  G | White and Black Caribbean  White and Black African  White and Asian  Any other mixed background / multiple ethnic background |
| 3 Asian | H  J  K  L  R | Indian  Pakistani  Bangladeshi  Any other Asian Background  Chinese |
| 4 Black | M  N  P | Caribbean  African  Any other Black background |
| 5 Other | S  T | Any other ethnic group  Arab |

| NER category | Data source code | Data source description |
| --- | --- | --- |
| 1 White | A  B | Any White Background, including Welsh, English, Scottish, Northern Irish, Irish, British  Gypsy or Irish Traveller |
| 2 Mixed | D  E  F  G | White and Black Caribbean  White and Black African  White and Asian  Any other mixed background / multiple ethnic background |
| 3 Indian | H | Indian |
| 4 Pakistani | J | Pakistani |
| 5 Bangladeshi | K | Bangladeshi |
| 6 Chinese | R | Chinese |
| 7 Black Caribbean | M | Caribbean |
| 8 Black African | N | African |
| 9 Other | L  P  S  T | Any other Asian Background  Any other Black background  Any other ethnic group  Arab |

### CTTP (COVID-19 Test Trace and Protect)

| ONS category | Data source code | Data source description |
| --- | --- | --- |
| 1 White |  | 'Albanian'  'Any other White background'  'Any White background'  'Bulgarian'  'Croatian'  'Czech'  'English, Welsh, Scottish, Northern Irish or British'  'EU Roma'  'French'  'German'  'Greek/Greek Cypriot'  'Gypsy or Irish Traveller'  'Hungarian'  'Irish'  'Italian'  'Latvian'  'Lithuanian'  'Maltese'  'Other White'  'Polish'  'Portuguese'  'Roma'  'Romanian'  'Russian'  'Scandinavian'  'Serbian'  'Slovakian'  'Slovenian'  'Spanish'  'Turkish/Turkish Cypriot'  'Ukrainian'  'White ? British'  'White European Other’ |
| 2 Mixed |  | 'Any Other Mixed Background'  'Any other mixed background'  'Any other Mixed/Multiple background'  'Black And Any Other Ethnic Group'  'Chinese And Any Other Ethnic Group'  'Other Mixed Background'  'White And Any Other Ethnic Group'  'White and Asian'  'White and Black African'  'White and Black Caribbean' |
| 3 Asian |  | 'Any other Asian background'  'Asian And Any Other Ethnic Group'  'Asian And Chinese'  'Bangladeshi'  'Chinese'  'Hong Kong Chinese'  'Indian'  'Japanese'  'Korean'  'Malaysian Chinese'  'Nepali'  'Other Asian'  'Other Pakistani'  'Pakistani'  'Sinhalese'  'Sri Lankan Tamil'  'Thai' |
| 4 Black |  | 'African'  'Any other Black background'  'Any other Black or African or Caribbean background'  'Black European'  'Black North American'  'Caribbean'  'Ghanaian'  'Nigerian'  'Other Black African'  'Somali'  'Sudanese' |
| 5 Other |  | 'Any other ethnic background'  'Any other ethnic group'  'Arab'  'Egyptian'  'Filipino'  'Irani'  'Iraqi'  'Kurdish'  'Latin/South/Central American'  'Lebanese'  'Libyan'  'Malay'  'Moroccan'  'Other Ethnic Group'  'Polynesian'  'Saudi Arabian'  'Syrian'  'Vietnamese'  'Yemeni' |

| NER category | Data source code | Data source description |
| --- | --- | --- |
| 1 White |  | 'Albanian'  'Any other White background'  'Any White background'  'Bulgarian'  'Croatian'  'Czech'  'English, Welsh, Scottish, Northern Irish or British'  'EU Roma'  'French'  'German'  'Greek/Greek Cypriot'  'Gypsy or Irish Traveller'  'Hungarian'  'Irish'  'Italian'  'Latvian'  'Lithuanian'  'Maltese'  'Other White'  'Polish'  'Portuguese'  'Roma'  'Romanian'  'Russian'  'Scandinavian'  'Serbian'  'Slovakian'  'Slovenian'  'Spanish'  'Turkish/Turkish Cypriot'  'Ukrainian'  'White ? British'  'White European Other’ |
| 2 Mixed |  | 'Any Other Mixed Background'  'Any other mixed background'  'Any other Mixed/Multiple background'  'Black And Any Other Ethnic Group'  'Chinese And Any Other Ethnic Group'  'Other Mixed Background'  'White And Any Other Ethnic Group'  'White and Asian'  'White and Black African'  'White and Black Caribbean' |
| 3 Indian |  | ‘Indian’ |
| 4 Pakistani |  | 'Other Pakistani'  'Pakistani' |
| 5 Bangladeshi |  | ‘Bangladeshi’ |
| 6 Chinese |  | 'Asian And Chinese'  'Chinese'  'Hong Kong Chinese'  'Malaysian Chinese' |
| 7 Black Caribbean |  | ‘Caribbean’ |
| 8 Black African |  | 'African'  'Ghanaian'  'Nigerian'  'Other Black African'  'Somali'  'Sudanese' |
| 9 Other |  | 'Any other Asian background'  'Any other Black background'  'Any other Black or African or Caribbean background'  'Any other ethnic background'  'Any other ethnic group'  'Arab'  'Asian And Any Other Ethnic Group'  'Asian And Chinese'  'Black European'  'Black North American'  'Egyptian'  'Filipino'  'Irani'  'Iraqi'  'Japanese'  'Korean'  'Kurdish'  'Latin/South/Central American'  'Lebanese'  'Libyan'  'Malay'  'Moroccan'  'Nepali'  'Other Asian'  'Other Ethnic Group'  'Polynesian'  'Saudi Arabian'  'Sinhalese'  'Sri Lankan Tamil'  'Syrian'  'Thai'  'Vietnamese'  'Yemeni' |

### CVLF (COVID-19 Lateral Flow)

| ONS category | Data source code | Data source description |
| --- | --- | --- |
| 1 White |  | 'Another White background'  'British~ English~ Northern Irish~ Scottish~ or Welsh'  'Irish Traveller or Gypsy'  'Irish'  'White' |
| 2 Mixed |  | 'Another Mixed background'  'Asian and White'  'Black African and White'  'Black Caribbean and White'  'Mixed or multiple ethnic groups' |
| 3 Asian |  | 'Another Asian background'  'Asian or Asian British'  'Bangladeshi'  'Chinese'  'Indian'  'Pakistani' |
| 4 Black |  | 'African'  'Another Black background'  'Black~ African~ Black British or Caribbean'  'Caribbean' |
| 5 Other |  | 'Another ethnic background'  'Another ethnic group'  'Arab' |

| NER category | Data source code | Data source description |
| --- | --- | --- |
| 1 White |  | 'Another White background'  'British~ English~ Northern Irish~ Scottish~ or Welsh'  'Irish Traveller or Gypsy'  'Irish'  'White' |
| 2 Mixed |  | 'Another Mixed background'  'Asian and White'  'Black African and White'  'Black Caribbean and White'  'Mixed or multiple ethnic groups' |
| 3 Indian |  | 'Indian' |
| 4 Pakistani |  | 'Pakistani' |
| 5 Bangladeshi |  | 'Bangladeshi' |
| 6 Chinese |  | 'Chinese' |
| 7 Black Caribbean |  | 'Caribbean' |
| 8 Black African |  | 'African' |
| 9 Other |  | 'Another Asian background'  'Another Black background'  'Another ethnic background'  'Another ethnic group'  'Arab'  'Asian or Asian British'  'Black~ African~ Black British or Caribbean' |

### CVVD (COVID-19 Vaccine Data)

| ONS category | Data source code | Data source description |
| --- | --- | --- |
| 1 White | A  B | Any White Background, including Welsh, English, Scottish, Northern Irish, Irish, British  Gypsy or Irish Traveller |
| 2 Mixed | D  E  F  G | White and Black Caribbean  White and Black African  White and Asian  Any other mixed background / multiple ethnic background |
| 3 Asian | H  J  K  L  R | Indian  Pakistani  Bangladeshi  Any other Asian Background  Chinese |
| 4 Black | M  N  P | Caribbean  African  Any other Black background |
| 5 Other | S  T | Any other ethnic group  Arab |

| NER category | Data source code | Data source description |
| --- | --- | --- |
| 1 White | A  B | Any White Background, including Welsh, English, Scottish, Northern Irish, Irish, British  Gypsy or Irish Traveller |
| 2 Mixed | D  E  F  G | White and Black Caribbean  White and Black African  White and Asian  Any other mixed background / multiple ethnic background |
| 3 Indian | H | Indian |
| 4 Pakistani | J | Pakistani |
| 5 Bangladeshi | K | Bangladeshi |
| 6 Chinese | R | Chinese |
| 7 Black Caribbean | M | Caribbean |
| 8 Black African | N | African |
| 9 Other | L  P  S  T | Any other Asian Background  Any other Black background  Any other ethnic group  Arab |

### CYFI (Cystic Fibrosis Register)

| ONS category | Data source code | Data source description |
| --- | --- | --- |
| 1 White |  | 'WBritish', 'WIrish', 'WOther' |
| 2 Mixed |  | ‘MWBC', 'MWAs', 'MWBA', 'Mother’ |
| 3 Asian |  | ‘ABangladeshi', 'AIndian', 'AOther', 'APakistani', 'OChinese' |
| 4 Black |  | 'BAfrican', 'BCaribbean', 'BOther' |
| 5 Other |  | 'OOther' |

| NER category | Data source code | Data source description |
| --- | --- | --- |
| 1 White |  | 'WBritish', 'WIrish', 'WOther' |
| 2 Mixed |  | ‘MWBC', 'MWAs', 'MWBA', 'Mother’ |
| 3 Indian |  | 'AIndian' |
| 4 Pakistani |  | 'APakistani' |
| 5 Bangladeshi |  | ‘ABangladeshi' |
| 6 Chinese |  | 'OChinese' |
| 7 Black Caribbean |  | 'BCaribbean' |
| 8 Black African |  | 'BAfrican', |
| 9 Other |  | 'AOther', 'BOther', 'OOther' |

### DSCW (Domiciliary Social Care Wales)

| ONS category | Data source code | Data source description |
| --- | --- | --- |
| 1 White |  | 'Any other White background, please state'  'European'  'Irish traveller'  'Welsh'  'White'  'White British'  'White English'  'White Irish'  'White Welsh'  'White or White British'  'White welsh'  'white welsh' |
| 2 Mixed |  | 'Any other mixed background'  'Mixed White and Asian'  'Mixed White and Black African'  'Mixed White and Black Caribbean'  'Mixed ethnic group' |
| 3 Asian |  | 'Any other Asian background, please state'  'Any other Chinese background'  'Asian or Asian British Bangladeshi'  'Asian or Asian British Indian'  'Asian or Asian British Pakistani'  'Bangladeshi'  'Chinese'  'Chinese British'  'Chinese or Chinese British Chinese'  'Indian'  'Pakistani' |
| 4 Black |  | 'Any other Black background, please state'  'Black African'  'Black Caribbean'  'Black or Black British African'  'Black or Black British Caribbean' |
| 5 Other |  | 'Any Other'  'Any other ethnic background, please state'  'Other Ethnicity' |

| NER category | Data source code | Data source description |
| --- | --- | --- |
| 1 White |  | 'Any other White background, please state'  'European'  'Irish traveller'  'Welsh'  'White'  'White British'  'White English'  'White Irish'  'White Welsh'  'White or White British'  'White welsh'  'white welsh' |
| 2 Mixed |  | 'Any other mixed background'  'Mixed White and Asian'  'Mixed White and Black African'  'Mixed White and Black Caribbean'  'Mixed ethnic group' |
| 3 Indian |  | 'Asian or Asian British Indian'  'Indian' |
| 4 Pakistani |  | 'Asian or Asian British Pakistani'  'Pakistani' |
| 5 Bangladeshi |  | 'Asian or Asian British Bangladeshi'  'Bangladeshi' |
| 6 Chinese |  | 'Any other Chinese background'  'Chinese'  'Chinese British'  'Chinese or Chinese British Chinese' |
| 7 Black Caribbean |  | 'Black Caribbean'  'Black or Black British Caribbean' |
| 8 Black African |  | Black African'  'Black or Black British African' |
| 9 Other |  | 'Any Other'  'Any other Asian background, please state'  'Any other Black background, please state'  'Any other ethnic background, please state'  'Other Ethnicity' |

### EDDS (Emergency Department DataSet)

| ONS category | Data source code | Data source description |
| --- | --- | --- |
| 1 White | 0  A  B | White  Any White Background, including Welsh, English, Scottish, Northern Irish, Irish, British  Gypsy or Irish Traveller |
| 2 Mixed | D  E  F  G | White and Black Caribbean  White and Black African  White and Asian  Any other mixed background / multiple ethnic background |
| 3 Asian | 4  5  6  7  H  J  K  L  R | Indian  Pakistani  Bangladeshi  Chinese  Indian  Pakistani  Bangladeshi  Any other Asian Background  Chinese |
| 4 Black | 1  2  3  M  N  P | Black - Caribbean  Black - African  Black - Other  Caribbean  African  Any other Black background |
| 5 Other | 8  S  T | Any other ethnic group  Any other ethnic group  Arab |

| NER category | Data source code | Data source description |
| --- | --- | --- |
| 1 White | 0  A  B | White  Any White Background, including Welsh, English, Scottish, Northern Irish, Irish, British  Gypsy or Irish Traveller |
| 2 Mixed | D  E  F  G | White and Black Caribbean  White and Black African  White and Asian  Any other mixed background / multiple ethnic background |
| 3 Indian | 4  H | Indian  Indian |
| 4 Pakistani | 5  J | Pakistani  Pakistani |
| 5 Bangladeshi | 6  K | Bangladeshi  Bangladeshi |
| 6 Chinese | 7  R | Chinese  Chinese |
| 7 Black Caribbean | 1  M | Black - Caribbean  Caribbean |
| 8 Black African | 2  N | Black - African  African |
| 9 Other | 3  8  L  P  S  T | Black - Other  Any other ethnic group  Any other Asian Background  Any other Black background  Any other ethnic group  Arab |

### EDUW (Education data)

| ONS category | Data source code | Data source description |
| --- | --- | --- |
| 1 White | WALB  WBGR  WBOS  WBRI  WBUL  WCRO  WCZE  WEEU  WEUR  WFRE  WGER  WGRE  WGRO  WHUN  WITA  WITH  WIRT  WKOS  WLAT  WLIT  WMAL  WMON  WNAG  WOBG  WOCC  WOER  WOGR  WOOG  WOOR  WOTG  WOTH  WOTR  WOTT  WOTW  WPOL  WPOR  WRMA  WROM  WRUS  WSCA  WSER  WSPA  WSVK  WSVN  WTUR  WUKR  WWEU | Albanian  British Gypsy / Gypsy Roma  Bosnian-Herzegovinian  White – British  Bulgarian  Croatian  Czech  White Eastern European  White European Other  French  German  Greek / Greek Cypriot  Gypsy / Gypsy Roma from Other Countries  Hungarian  Italian  Traveller of Irish Heritage  Traveller  Kosovan  Latvian  Lithuanian  Maltese  Montenegran  New Traveller  British Gypsy  Occupational Traveller  EU Roma  Other Gypsy / Gypsy Roma  Gypsy from Other Countries  Roma from Other Countries  Other Gypsy  Any other White Background  Other Roma  Other Traveller  Other White  Polish  Portuguese  Romanian  Gypsy / Gypsy Roma  Russian  Scandinavian  Serbian  Spanish  Slovakian  Slovenian  Turkish / Turkish Cypriot  Ukrainian  White Western European |
| 2 Mixed | MABL  MACH  MAOE  MBCH  MBOE  MCOE  MOTH  MOTM  MWAS  MWBA  MWBC  MWCH  MWOE | Asian and Black  Asian and Chinese  Asian and Any Other Ethnic Group  Black and Chinese  Black and Other Ethnic Group  Chinese and Any Other Ethnic Group  Any Other Mixed Background  Other Mixed Background  White and Asian  White and Black African  White and Black Caribbean  White and Chinese  White and Any Other Ethnic Group |
| 3 Asian | AAFR  ABAN  AIND  AKAS  AMPK  ANEP  AOTA  AOTH  APOK  APKN  ASNL  ASLT  CHKC  CHNE  CMAL  CSNG  CTWN  OFIL  OJPN  OKOR  OMAL  OOCH | African Asian  Bangladeshi  Indian  Kashmiri  Mirpuri Pakistani  Nepali  Other Asian  Any Other Asian Background  Other Pakistani  Pakistani  Sinhalese  Sri Lankan Tamil  Hong Kong Chinese  Chinese or Chinese British  Malaysian Chinese  Singaporean Chinese  Taiwanese  Filipino  Japanese  Korean  Malay  Other Chinese |
| 4 Black | BAFR  BAOF  BCRB  BERI  BEUR  BGHA  BNAM  BOTB  BOTH  BNGN  BSLN  BSOM  BSUD | African  Other Black African  Caribbean  Eritrean  Black European  Ghanaian  Black North American  Other Black  Any other Black background  Nigerian  Sierra Leonian  Somali  Sudanese |
| 5 Other | OAFG  OARA  OEGY  OIRN  OIRQ  OKRD  OLAM  OLEB  OLIB  OMRC  OOEG  OOTH  OPOL  OSYR  OTHA  OSAU  OVIE  OYEM | Afghanistani  Arab  Egyptian  Iranian  Iraqi  Kurdish  Latin / South / Central American  Lebanese  Libyan  Moroccan  Other ethnic group  Any other ethnic background  Polynesian  Syrian  Thai  Saudi Arabian  Vietnamese  Yemeni |

| NER category | Data source code | Data source description |
| --- | --- | --- |
| 1 White | WALB  WBGR  WBOS  WBRI  WBUL  WCRO  WCZE  WEEU  WEUR  WFRE  WGER  WGRE  WGRO  WHUN  WITA  WITH  WIRT  WKOS  WLAT  WLIT  WMAL  WMON  WNAG  WOBG  WOCC  WOER  WOGR  WOOG  WOOR  WOTG  WOTH  WOTR  WOTT  WOTW  WPOL  WPOR  WRMA  WROM  WRUS  WSCA  WSER  WSPA  WSVK  WSVN  WTUR  WUKR  WWEU | Albanian  British Gypsy / Gypsy Roma  Bosnian-Herzegovinian  White – British  Bulgarian  Croatian  Czech  White Eastern European  White European Other  French  German  Greek / Greek Cypriot  Gypsy / Gypsy Roma from Other Countries  Hungarian  Italian  Traveller of Irish Heritage  Traveller  Kosovan  Latvian  Lithuanian  Maltese  Montenegran  New Traveller  British Gypsy  Occupational Traveller  EU Roma  Other Gypsy / Gypsy Roma  Gypsy from Other Countries  Roma from Other Countries  Other Gypsy  Any other White Background  Other Roma  Other Traveller  Other White  Polish  Portuguese  Romanian  Gypsy / Gypsy Roma  Russian  Scandinavian  Serbian  Spanish  Slovakian  Slovenian  Turkish / Turkish Cypriot  Ukrainian  White Western European |
| 2 Mixed | MABL  MACH  MAOE  MBCH  MBOE  MCOE  MOTH  MOTM  MWAS  MWBA  MWBC  MWCH  MWOE | Asian and Black  Asian and Chinese  Asian and Any Other Ethnic Group  Black and Chinese  Black and Other Ethnic Group  Chinese and Any Other Ethnic Group  Any Other Mixed Background  Other Mixed Background  White and Asian  White and Black African  White and Black Caribbean  White and Chinese  White and Any Other Ethnic Group |
| 3 Indian | AIND | Indian |
| 4 Pakistani | AMPK  AOPK  APKN | Mirpuri Pakistani  Other Pakistani  Pakistani |
| 5 Bangladeshi | ABAN | Bangladeshi |
| 6 Chinese | CHKC  CHNE  CMAL  CSNG  CTWN  OOCH | Hong Kong Chinese  Chinese or Chinese British  Malaysian Chinese  Singaporean Chinese  Taiwanese  Other Chinese |
| 7 Black Caribbean | BCRB | Caribbean |
| 8 Black African | BAFR  BAOF  BERI  BGHA  BNGN  BSLN  BSOM  BSUD | African  Other Black African  Eritrean  Ghanaian  Nigerian  Sierra Leonian  Somali  Sudanese |
| 9 Other | AAFR  AKAS  ANEP  AOTA  AOTH  ASNL  ASLT  BEUR  BNAM  BOTB  BOTH  OAFG  OARA  OEGY  OFIL  OIRN  OIRQ  OJPN  OKOR  OKRD  OLAM  OLEB  OLIB  OMAL  OOEG  OOTH  OPOL  OSAU  OSYR  OTHA  OVIE  OYEM | African Asian  Kashmiri  Nepali  Other Asian  Any Other Asian Background  Sinhalese  Sri Lankan Tamil  Black European  Black North American  Other Black  Any other Black background  Afghanistani  Arab  Egyptian  Filipino  Iranian  Iraqi  Japanese  Korean  Kurdish  Latin / South / Central American  Lebanese  Libyan  Malay  Other ethnic group  Any other ethnic background  Polynesian  Saudi Arabian  Syrian  Thai  Vietnamese  Yemeni |

### HWRA (Health Worker Risk Assessment)

| ONS category | Data source code | Data source description |
| --- | --- | --- |
| 1 White |  | 'A White - British'  'B White - Irish'  'C White - Any other White background'  'C2 White Northern Irish'  'C3 White Unspecified'  'CA White English'  'CB White Scottish'  'CC White Welsh'  'CD White Cornish'  'CE White Cypriot (non specific)'  'CF White Greek'  'CG White Greek Cypriot'  'CH White Turkish'  'CJ White Turkish Cypriot'  'CK White Italian'  'CL White Irish Traveller'  'CM White Traveller'  'CN White Gypsy/Romany'  'CP White Polish'  'CQ White ex-USSR'  'CU White Croatian'  'CV White Serbian'  'CW White Other Ex-Yugoslav'  'CY White Other European' |
| 2 Mixed |  | 'CX White Mixed'  'D Mixed - White & Black Caribbean'  'E Mixed - White & Black African'  'F Mixed - White & Asian'  'G Mixed - Any other mixed background'  'GA Mixed - Black & Asian'  'GB Mixed - Black & Chinese'  'GC Mixed - Black & White'  'GD Mixed - Chinese & White'  'GE Mixed - Asian & Chinese'  'GF Mixed - Other/Unspecified' |
| 3 Asian |  | 'H Asian or Asian British - Indian'  'J Asian or Asian British - Pakistani'  'K Asian or Asian British - Bangladeshi'  'L Asian or Asian British - Any other Asian background'  'LA Asian Mixed'  'LB Asian Punjabi'  'LC Asian Kashmiri'  'LD Asian East African'  'LE Asian Sri Lankan'  'LF Asian Tamil'  'LG Asian Sinhalese'  'LH Asian British'  'LK Asian Unspecified'  'R Chinese'  'SA Vietnamese'  'SB Japanese'  'SC Filipino'  'SD Malaysian' |
| 4 Black |  | 'M Black or Black British - Caribbean'  'N Black or Black British - African'  'P Black or Black British - Any other Black background'  'PA Black Somali'  'PB Black Mixed'  'PC Black Nigerian'  'PD Black British'  'PE Black Unspecified' |
| 5 Other |  | 'SE Other Specified' |

| NER category | Data source code | Data source description |
| --- | --- | --- |
| 1 White |  | 'A White - British'  'B White - Irish'  'C White - Any other White background'  'C2 White Northern Irish'  'C3 White Unspecified'  'CA White English'  'CB White Scottish'  'CC White Welsh'  'CD White Cornish'  'CE White Cypriot (non specific)'  'CF White Greek'  'CG White Greek Cypriot'  'CH White Turkish'  'CJ White Turkish Cypriot'  'CK White Italian'  'CL White Irish Traveller'  'CM White Traveller'  'CN White Gypsy/Romany'  'CP White Polish'  'CQ White ex-USSR'  'CU White Croatian'  'CV White Serbian'  'CW White Other Ex-Yugoslav'  'CY White Other European' |
| 2 Mixed |  | 'CX White Mixed'  'D Mixed - White & Black Caribbean'  'E Mixed - White & Black African'  'F Mixed - White & Asian'  'G Mixed - Any other mixed background'  'GA Mixed - Black & Asian'  'GB Mixed - Black & Chinese'  'GC Mixed - Black & White'  'GD Mixed - Chinese & White'  'GE Mixed - Asian & Chinese'  'GF Mixed - Other/Unspecified' |
| 3 Indian |  | 'H Asian or Asian British – Indian’ |
| 4 Pakistani |  | 'J Asian or Asian British - Pakistani' |
| 5 Bangladeshi |  | 'K Asian or Asian British - Bangladeshi' |
| 6 Chinese |  | 'R Chinese' |
| 7 Black Caribbean |  | 'M Black or Black British - Caribbean' |
| 8 Black African |  | 'N Black or Black British - African'  'PA Black Somali'  'PC Black Nigerian' |
| 9 Other |  | 'L Asian or Asian British - Any other Asian background'  'LA Asian Mixed'  'LB Asian Punjabi'  'LC Asian Kashmiri'  'LD Asian East African'  'LE Asian Sri Lankan'  'LF Asian Tamil'  'LG Asian Sinhalese'  'LH Asian British'  'LK Asian Unspecified'  'P Black or Black British - Any other Black background'  'PB Black Mixed'  'PD Black British'  'PE Black Unspecified'  'SA Vietnamese'  'SB Japanese'  'SC Filipino'  'SD Malaysian'  'SE Other Specified' |

### ICNC (ICNARC Intensive Care National Audit Research Centre)

| ONS category | Data source code | Data source description |
| --- | --- | --- |
| 1 White | A  B  C | White, British  White, Irish  White, Any other |
| 2 Mixed | D  E  F  G | Mixed, White and Black Caribbean  Mixed, White and Black African  Mixed, White and Asian  Mixed, Any other |
| 3 Asian | H  J  K  L  R | Asian, Indian  Asian, Pakistani  Asian, Bangladeshi  Asian, Any other  Asian, Chinese |
| 4 Black | M  N  P | Black, Caribbean  Black, African  Black, Any other |
| 5 Other | S | Any other ethnic group |

| NER category | Data source code | Data source description |
| --- | --- | --- |
| 1 White | A  B  C | White, British  White, Irish  White, Any other |
| 2 Mixed | D  E  F  G | Mixed, White and Black Caribbean  Mixed, White and Black African  Mixed, White and Asian  Mixed, Any other |
| 3 Indian | H | Asian, Indian |
| 4 Pakistani | J | Asian, Pakistani |
| 5 Bangladeshi | K | Asian, Bangladeshi |
| 6 Chinese | R | Asian, Chinese |
| 7 Black Caribbean | M | Black, Caribbean |
| 8 Black African | N | Black, African |
| 9 Other | L  P  S | Asian, Any other  Black, Any other  Any other ethnic group |

### LACW (Looked After Children Wales)

| ONS category | Data source code | Data source description |
| --- | --- | --- |
| 1 White | A1  A2  A3  WHTE | White British  White Irish  White Other  White |
| 2 Mixed | B1  B2  B3  B4  MIXD | White and Black Caribbean  White and Black African  White and Asian  Mixed Background Other  Mixed ethnic groups |
| 3 Asian | ASAB  C1  C2  C3  C4  E1 | Asian or Asian British  Indian  Pakistani  Bangladeshi  Asian Other  Chinese |
| 4 Black | BBAC  D1  D2  D3 | Black; African; Caribbean or Black British  Caribbean  African  Black Background Other |
| 5 Other | E2  OOTH | Any Other Ethnic Group  Other ethnic group |

| NER category | Data source code | Data source description |
| --- | --- | --- |
| 1 White | A1  A2  A3  WHTE | White British  White Irish  White Other  White |
| 2 Mixed | B1  B2  B3  B4  MIXD | White and Black Caribbean  White and Black African  White and Asian  Mixed Background Other  Mixed ethnic groups |
| 3 Indian | C1 | Indian |
| 4 Pakistani | C2 | Pakistani |
| 5 Bangladeshi | C3 | Bangladeshi |
| 6 Chinese | E1 | Chinese |
| 7 Black Caribbean | D1 | Caribbean |
| 8 Black African | D2 | African |
| 9 Other | ASAB  BBAC  C4  D3  E2  OOTH | Asian or Asian British  Black; African; Caribbean or Black British  Asian Other  Black Background Other  Any Other Ethnic Group  Other ethnic group |

### MIDS (Maternity Indicators DataSet)

| ONS category | Data source code | Data source description |
| --- | --- | --- |
| 1 White | A  B | Any White Background, including Welsh, English, Scottish, Northern Irish, Irish, British  Gypsy or Irish Traveller |
| 2 Mixed | D  E  F  G | White and Black Caribbean  White and Black African  White and Asian  Any other mixed background / multiple ethnic background |
| 3 Asian | H  J  K  L  R | Indian  Pakistani  Bangladeshi  Any other Asian Background  Chinese |
| 4 Black | M  N  P | Caribbean  African  Any other Black background |
| 5 Other | S  T | Any other ethnic group  Arab |

| NER category | Data source code | Data source description |
| --- | --- | --- |
| 1 White | A  B | Any White Background, including Welsh, English, Scottish, Northern Irish, Irish, British  Gypsy or Irish Traveller |
| 2 Mixed | D  E  F  G | White and Black Caribbean  White and Black African  White and Asian  Any other mixed background / multiple ethnic background |
| 3 Indian | H | Indian |
| 4 Pakistani | J | Pakistani |
| 5 Bangladeshi | K | Bangladeshi |
| 6 Chinese | R | Chinese |
| 7 Black Caribbean | M | Caribbean |
| 8 Black African | N | African |
| 9 Other | L  P  S  T | Any other Asian Background  Any other Black background  Any other ethnic group  Arab |

### NCCH (National Community Child Health data)

| ONS category | Data source code | Data source description |
| --- | --- | --- |
| 1 White | 0  A  B | White  Any White Background, including Welsh, English, Scottish, Northern Irish, Irish, British  Gypsy or Irish Traveller |
| 2 Mixed | D  E  F  G | White and Black Caribbean  White and Black African  White and Asian  Any other mixed background / multiple ethnic background |
| 3 Asian | 4  5  6  7  H  J  K  L  R | Indian  Pakistani  Bangladeshi  Chinese  Indian  Pakistani  Bangladeshi  Any other Asian Background  Chinese |
| 4 Black | 1  2  3  M  N  P | Black - Caribbean  Black - African  Black - Other  Caribbean  African  Any other Black background |
| 5 Other | 8  S  T | Any other ethnic group  Any other ethnic group  Arab |

| NER category | Data source code | Data source description |
| --- | --- | --- |
| 1 White | 0  A  B | White  Any White Background, including Welsh, English, Scottish, Northern Irish, Irish, British  Gypsy or Irish Traveller |
| 2 Mixed | D  E  F  G | White and Black Caribbean  White and Black African  White and Asian  Any other mixed background / multiple ethnic background |
| 3 Indian | 4  H | Indian  Indian |
| 4 Pakistani | 5  J | Pakistani  Pakistani |
| 5 Bangladeshi | 6  K | Bangladeshi  Bangladeshi |
| 6 Chinese | 7  R | Chinese  Chinese |
| 7 Black Caribbean | 1  M | Black - Caribbean  Caribbean |
| 8 Black African | 2  N | Black - African  African |
| 9 Other | 3  8  L  P  S  T | Black - Other  Any other ethnic group  Any other Asian Background  Any other Black background  Any other ethnic group  Arab |

### NHSO (NHS 111)

| ONS category | Data source code | Data source description |
| --- | --- | --- |
| 1 White |  | 'White - British'  'White - Any other White background'  'White - Irish' |
| 2 Mixed |  | 'Mixed - White and Black Caribbean'  'Mixed - White and Asian'  'Mixed - White and Black African'  'Mixed - Any other mixed background' |
| 3 Asian |  | 'Asian or Asian British - Indian'  'Asian or Asian British - Pakistani'  'Asian or Asian British - Bangladeshi'  'Other ethnic groups - Chinese'  'Asian or Asian British - Any other Asian background' |
| 4 Black |  | 'Black or Black British - Caribbean'  'Black or Black British - African'  'Black or Black British - Any other Black background' |
| 5 Other |  | 'Other ethnic groups - Any other ethnic group' |

| NER category | Data source code | Data source description |
| --- | --- | --- |
| 1 White |  | 'White - British'  'White - Any other White background'  'White - Irish' |
| 2 Mixed |  | 'Mixed - White and Black Caribbean'  'Mixed - White and Asian'  'Mixed - White and Black African'  'Mixed - Any other mixed background' |
| 3 Indian |  | 'Asian or Asian British - Indian' |
| 4 Pakistani |  | 'Asian or Asian British - Pakistani' |
| 5 Bangladeshi |  | 'Asian or Asian British - Bangladeshi' |
| 6 Chinese |  | 'Other ethnic groups - Chinese' |
| 7 Black Caribbean |  | 'Black or Black British - Caribbean' |
| 8 Black African |  | 'Black or Black British - African' |
| 9 Other |  | 'Asian or Asian British - Any other Asian background'  'Black or Black British - Any other Black background'  'Other ethnic groups - Any other ethnic group' |

### NSWD (National Survey for Wales Data)

| ONS category | Data source code | Data source description |
| --- | --- | --- |
| 1 White | 1  2  3  4  19 | White - Welsh/English/Scottish/Northern Irish/British  White - Irish  White - Gypsy or Irish Traveller  White – Other  White - Polish (SPONTANEOUS ONLY) |
| 2 Mixed | 5  6  7  8 | Mixed - White and Black Caribbean  Mixed - White and Black African  Mixed - White and Asian  Mixed - Other |
| 3 Asian | 9  10  11  12  13 | Asian - Indian  Asian - Pakistani  Asian - Bangladeshi  Asian - Chinese  Asian - Other |
| 4 Black | 14  15  16 | Black - African  Black - Caribbean  Black - Other |
| 5 Other | 17  18 | Other - Arab  Other - Any other ethnic group |

| NER category | Data source code | Data source description |
| --- | --- | --- |
| 1 White | 1  2  3  4  19 | White - Welsh/English/Scottish/Northern Irish/British  White - Irish  White - Gypsy or Irish Traveller  White – Other  White - Polish (SPONTANEOUS ONLY) |
| 2 Mixed | 5  6  7  8 | Mixed - White and Black Caribbean  Mixed - White and Black African  Mixed - White and Asian  Mixed - Other |
| 3 Indian | 9 | Asian - Indian |
| 4 Pakistani | 10 | Asian - Pakistani |
| 5 Bangladeshi | 11 | Asian - Bangladeshi |
| 6 Chinese | 12 | Asian - Chinese |
| 7 Black Caribbean | 15 | Black - Caribbean |
| 8 Black African | 14 | Black - African |
| 9 Other | 13  16  17  18 | Asian – Other  Black – Other  Other - Arab  Other - Any other ethnic group |

### OPRD (Out Patient Referrals Dataset)

| ONS category | Data source code | Data source description |
| --- | --- | --- |
| 1 White | 0  A  B | White  Any White Background, including Welsh, English, Scottish, Northern Irish, Irish, British  Gypsy or Irish Traveller |
| 2 Mixed | D  E  F  G | White and Black Caribbean  White and Black African  White and Asian  Any other mixed background / multiple ethnic background |
| 3 Asian | 4  5  6  7  H  J  K  L  R | Indian  Pakistani  Bangladeshi  Chinese  Indian  Pakistani  Bangladeshi  Any other Asian Background  Chinese |
| 4 Black | 1  2  3  M  N  P | Black - Caribbean  Black - African  Black - Other  Caribbean  African  Any other Black background |
| 5 Other | 8  S  T | Any other ethnic group  Any other ethnic group  Arab |

| NER category | Data source code | Data source description |
| --- | --- | --- |
| 1 White | 0  A  B | White  Any White Background, including Welsh, English, Scottish, Northern Irish, Irish, British  Gypsy or Irish Traveller |
| 2 Mixed | D  E  F  G | White and Black Caribbean  White and Black African  White and Asian  Any other mixed background / multiple ethnic background |
| 3 Indian | 4  H | Indian  Indian |
| 4 Pakistani | 5  J | Pakistani  Pakistani |
| 5 Bangladeshi | 6  K | Bangladeshi  Bangladeshi |
| 6 Chinese | 7  R | Chinese  Chinese |
| 7 Black Caribbean | 1  M | Black - Caribbean  Caribbean |
| 8 Black African | 2  N | Black - African  African |
| 9 Other | 3  8  L  P  S  T | Black - Other  Any other ethnic group  Any other Asian Background  Any other Black background  Any other ethnic group  Arab |

### PEDW (Patient Episode Database for Wales)

| ONS category | Data source code | Data source description |
| --- | --- | --- |
| 1 White | 0  A  B | White  Any White Background, including Welsh, English, Scottish, Northern Irish, Irish, British  Gypsy or Irish Traveller |
| 2 Mixed | D  E  F  G | White and Black Caribbean  White and Black African  White and Asian  Any other mixed background / multiple ethnic background |
| 3 Asian | 4  5  6  7  H  J  K  L  R | Indian  Pakistani  Bangladeshi  Chinese  Indian  Pakistani  Bangladeshi  Any other Asian Background  Chinese |
| 4 Black | 1  2  3  M  N  P | Black - Caribbean  Black - African  Black - Other  Caribbean  African  Any other Black background |
| 5 Other | 8  S  T | Any other ethnic group  Any other ethnic group  Arab |

| NER category | Data source code | Data source description |
| --- | --- | --- |
| 1 White | 0  A  B | White  Any White Background, including Welsh, English, Scottish, Northern Irish, Irish, British  Gypsy or Irish Traveller |
| 2 Mixed | D  E  F  G | White and Black Caribbean  White and Black African  White and Asian  Any other mixed background / multiple ethnic background |
| 3 Indian | 4  H | Indian  Indian |
| 4 Pakistani | 5  J | Pakistani  Pakistani |
| 5 Bangladeshi | 6  K | Bangladeshi  Bangladeshi |
| 6 Chinese | 7  R | Chinese  Chinese |
| 7 Black Caribbean | 1  M | Black - Caribbean  Caribbean |
| 8 Black African | 2  N | Black - African  African |
| 9 Other | 3  8  L  P  S  T | Black - Other  Any other ethnic group  Any other Asian Background  Any other Black background  Any other ethnic group  Arab |

### SACT (Systematic Anti Cancer Therapy)

| ONS category | Data source code | Data source description |
| --- | --- | --- |
| 1 White |  | 'Any White' |
| 2 Mixed |  | 'Mixed White and Asian'  'Mixed White and Black African'  'Mixed White and Black Caribbean'  'Other mixed' |
| 3 Asian |  | 'Bangladeshi'  'Chinese'  'Indian  'Other Asian',  'Pakistani' |
| 4 Black |  | 'Other Black'  'Black Caribbean'  'Black African' |
| 5 Other |  | 'Other' |

| NER category | Data source code | Data source description |
| --- | --- | --- |
| 1 White |  | 'Any White' |
| 2 Mixed |  | 'Mixed White and Asian'  'Mixed White and Black African'  'Mixed White and Black Caribbean'  'Other mixed' |
| 3 Indian |  | 'Indian' |
| 4 Pakistani |  | 'Pakistani' |
| 5 Bangladeshi |  | 'Bangladeshi' |
| 6 Chinese |  | 'Chinese' |
| 7 Black Caribbean |  | 'Black Caribbean' |
| 8 Black African |  | 'Black African' |
| 9 Other |  | 'Other Asian'  'Other Black'  'Other' |

### SMDS (Substance Misuse DataSet)

| ONS category | Data source code | Data source description |
| --- | --- | --- |
| 1 White | 0  A  B | White  Any White Background, including Welsh, English, Scottish, Northern Irish, Irish, British  Gypsy or Irish Traveller |
| 2 Mixed | D  E  F  G | White and Black Caribbean  White and Black African  White and Asian  Any other mixed background / multiple ethnic background |
| 3 Asian | 4  5  6  7  H  J  K  L  R | Indian  Pakistani  Bangladeshi  Chinese  Indian  Pakistani  Bangladeshi  Any other Asian Background  Chinese |
| 4 Black | 1  2  3  M  N  P | Black - Caribbean  Black - African  Black - Other  Caribbean  African  Any other Black background |
| 5 Other | 8  S  T | Any other ethnic group  Any other ethnic group  Arab |

| NER category | Data source code | Data source description |
| --- | --- | --- |
| 1 White | 0  A  B | White  Any White Background, including Welsh, English, Scottish, Northern Irish, Irish, British  Gypsy or Irish Traveller |
| 2 Mixed | D  E  F  G | White and Black Caribbean  White and Black African  White and Asian  Any other mixed background / multiple ethnic background |
| 3 Indian | 4  H | Indian  Indian |
| 4 Pakistani | 5  J | Pakistani  Pakistani |
| 5 Bangladeshi | 6  K | Bangladeshi  Bangladeshi |
| 6 Chinese | 7  R | Chinese  Chinese |
| 7 Black Caribbean | 1  M | Black - Caribbean  Caribbean |
| 8 Black African | 2  N | Black - African  African |
| 9 Other | 3  8  L  P  S  T | Black - Other  Any other ethnic group  Any other Asian Background  Any other Black background  Any other ethnic group  Arab |

### SWAC (School Workforce Annual Census)

| ONS category | Data source code | Data source description |
| --- | --- | --- |
| 1 White | WALB  WBGR  WBOS  WBRI  WBUL  WCRO  WCZE  WEEU  WEUR  WFRE  WGER  WGRE  WGRO  WHUN  WITA  WITH  WIRT  WKOS  WLAT  WLIT  WMAL  WMON  WNAG  WOBG  WOCC  WOER  WOGR  WOOG  WOOR  WOTG  WOTH  WOTR  WOTT  WOTW  WPOL  WPOR  WRMA  WROM  WRUS  WSCA  WSER  WSPA  WSVK  WSVN  WTUR  WUKR  WWEU | Albanian  British Gypsy / Gypsy Roma  Bosnian-Herzegovinian  White – British  Bulgarian  Croatian  Czech  White Eastern European  White European Other  French  German  Greek / Greek Cypriot  Gypsy / Gypsy Roma from Other Countries  Hungarian  Italian  Traveller of Irish Heritage  Traveller  Kosovan  Latvian  Lithuanian  Maltese  Montenegran  New Traveller  British Gypsy  Occupational Traveller  EU Roma  Other Gypsy / Gypsy Roma  Gypsy from Other Countries  Roma from Other Countries  Other Gypsy  Any other White Background  Other Roma  Other Traveller  Other White  Polish  Portuguese  Romanian  Gypsy / Gypsy Roma  Russian  Scandinavian  Serbian  Spanish  Slovakian  Slovenian  Turkish / Turkish Cypriot  Ukrainian  White Western European |
| 2 Mixed | MABL  MACH  MAOE  MBCH  MBOE  MCOE  MOTH  MOTM  MWAS  MWBA  MWBC  MWCH  MWOE | Asian and Black  Asian and Chinese  Asian and Any Other Ethnic Group  Black and Chinese  Black and Other Ethnic Group  Chinese and Any Other Ethnic Group  Any Other Mixed Background  Other Mixed Background  White and Asian  White and Black African  White and Black Caribbean  White and Chinese  White and Any Other Ethnic Group |
| 3 Asian | AAFR  ABAN  AIND  AKAS  AMPK  ANEP  AOTA  AOTH  APOK  APKN  ASNL  ASLT  CHKC  CHNE  CMAL  CSNG  CTWN  OFIL  OJPN  OKOR  OMAL  OOCH | African Asian  Bangladeshi  Indian  Kashmiri  Mirpuri Pakistani  Nepali  Other Asian  Any Other Asian Background  Other Pakistani  Pakistani  Sinhalese  Sri Lankan Tamil  Hong Kong Chinese  Chinese or Chinese British  Malaysian Chinese  Singaporean Chinese  Taiwanese  Filipino  Japanese  Korean  Malay  Other Chinese |
| 4 Black | BAFR  BAOF  BCRB  BERI  BEUR  BGHA  BNAM  BOTB  BOTH  BNGN  BSLN  BSOM  BSUD | African  Other Black African  Caribbean  Eritrean  Black European  Ghanaian  Black North American  Other Black  Any other Black background  Nigerian  Sierra Leonian  Somali  Sudanese |
| 5 Other | OAFG  OARA  OEGY  OIRN  OIRQ  OKRD  OLAM  OLEB  OLIB  OMRC  OOEG  OOTH  OPOL  OSYR  OTHA  OSAU  OVIE  OYEM | Afghanistani  Arab  Egyptian  Iranian  Iraqi  Kurdish  Latin / South / Central American  Lebanese  Libyan  Moroccan  Other ethnic group  Any other ethnic background  Polynesian  Syrian  Thai  Saudi Arabian  Vietnamese  Yemeni |

| NER category | Data source code | Data source description |
| --- | --- | --- |
| 1 White | WALB  WBGR  WBOS  WBRI  WBUL  WCRO  WCZE  WEEU  WEUR  WFRE  WGER  WGRE  WGRO  WHUN  WITA  WITH  WIRT  WKOS  WLAT  WLIT  WMAL  WMON  WNAG  WOBG  WOCC  WOER  WOGR  WOOG  WOOR  WOTG  WOTH  WOTR  WOTT  WOTW  WPOL  WPOR  WRMA  WROM  WRUS  WSCA  WSER  WSPA  WSVK  WSVN  WTUR  WUKR  WWEU | Albanian  British Gypsy / Gypsy Roma  Bosnian-Herzegovinian  White – British  Bulgarian  Croatian  Czech  White Eastern European  White European Other  French  German  Greek / Greek Cypriot  Gypsy / Gypsy Roma from Other Countries  Hungarian  Italian  Traveller of Irish Heritage  Traveller  Kosovan  Latvian  Lithuanian  Maltese  Montenegran  New Traveller  British Gypsy  Occupational Traveller  EU Roma  Other Gypsy / Gypsy Roma  Gypsy from Other Countries  Roma from Other Countries  Other Gypsy  Any other White Background  Other Roma  Other Traveller  Other White  Polish  Portuguese  Romanian  Gypsy / Gypsy Roma  Russian  Scandinavian  Serbian  Spanish  Slovakian  Slovenian  Turkish / Turkish Cypriot  Ukrainian  White Western European |
| 2 Mixed | MABL  MACH  MAOE  MBCH  MBOE  MCOE  MOTH  MOTM  MWAS  MWBA  MWBC  MWCH  MWOE | Asian and Black  Asian and Chinese  Asian and Any Other Ethnic Group  Black and Chinese  Black and Other Ethnic Group  Chinese and Any Other Ethnic Group  Any Other Mixed Background  Other Mixed Background  White and Asian  White and Black African  White and Black Caribbean  White and Chinese  White and Any Other Ethnic Group |
| 3 Indian | AIND | Indian |
| 4 Pakistani | AMPK  AOPK  APKN | Mirpuri Pakistani  Other Pakistani  Pakistani |
| 5 Bangladeshi | ABAN | Bangladeshi |
| 6 Chinese | CHKC  CHNE  CMAL  CSNG  CTWN  OOCH | Hong Kong Chinese  Chinese or Chinese British  Malaysian Chinese  Singaporean Chinese  Taiwanese  Other Chinese |
| 7 Black Caribbean | BCRB | Caribbean |
| 8 Black African | BAFR  BAOF  BERI  BGHA  BNGN  BSLN  BSOM  BSUD | African  Other Black African  Eritrean  Ghanaian  Nigerian  Sierra Leonian  Somali  Sudanese |
| 9 Other | AAFR  AKAS  ANEP  AOTA  AOTH  ASNL  ASLT  BEUR  BNAM  BOTB  OAFG  OARA  OFIL  OIRN  OIRQ  OJPN  OKOR  OKRD  OLAM  OLEB  OLIB  OMAL  OOEG  OOTH  OPOL  OSAU  OSYR  OTHA  OVIE  OYEM | African Asian  Kashmiri  Nepali  Other Asian  Any Other Asian Background  Sinhalese  Sri Lankan Tamil  Black European  Black North American  Other Black  Afghanistani  Arab  Filipino  Iranian  Iraqi  Japanese  Korean  Kurdish  Latin / South / Central American  Lebanese  Libyan  Malay  Other ethnic group  Any other ethnic background  Polynesian  Saudi Arabian  Syrian  Thai  Vietnamese  Yemeni |

### WASD (Welsh Ambulance Service Dataset)

| ONS category | Data source code | Data source description |
| --- | --- | --- |
| 1 White | 1  2  3 | British  Irish  Any other white background |
| 2 Mixed | 4  5  6  7 | White and black Caribbean  White and black African  White and Asian  Any other mixed background |
| 3 Asian | 8  9  10  11  15 | Indian  Pakistani  Bangladeshi  Any other Asian background  Chinese |
| 4 Black | 12  13  14 | Caribbean  African  Any other Black background |
| 5 Other | 16 | Any other ethnic group |

| NER category | Data source code | Data source description |
| --- | --- | --- |
| 1 White | 1  2  3 | British  Irish  Any other white background |
| 2 Mixed | 4  5  6  7 | White and black Caribbean  White and black African  White and Asian  Any other mixed background |
| 3 Indian | 8 | Indian |
| 4 Pakistani | 9 | Pakistani |
| 5 Bangladeshi | 10 | Bangladeshi |
| 6 Chinese | 15 | Chinese |
| 7 Black Caribbean | 12 | Caribbean |
| 8 Black African | 13 | Africa |
| 9 Other | 11  14  16 | Any other Asian background  Any other Black background  Any other ethnic group |

### WLGP (Wales Longitudinal General Practice)

| ONS category | Data source code | Data source description |
| --- | --- | --- |
| 1 White | '9i0..'  '9i00.'  '9i1..'  '9i10.'  '9i2..'  '9i20.'  '9i21.'  '9i22.'  '9i23.'  '9i24.'  '9i25.'  '9i26.'  '9i27.'  '9i28.'  '9i29.'  '9i2A.'  '9i2B.'  '9i2C.'  '9i2D.'  '9i2E.'  '9i2F.'  '9i2G.'  '9i2H.'  '9i2J.'  '9i2K.'  '9i2L.'  '9i2M.'  '9i2N.'  '9i2P.'  '9i2Q.'  '9i2R.'  '9i2S.'  '9i2T.'  '9S1..'  '9S10.'  '9S11.'  '9S12.'  '9S13.'  '9S14.'  '9SA9.'  '9SAA.'  '9SAB.'  '9SAC.'  '9SI..'  '9t00.'  '9t01.'  '9t02.'  '9t03.'  '9t10.'  '9t11.'  '9t20.'  '9t21.'  '9t22.'  '9t24.'  '9t25.' | British or mixed British - ethnic category 2001 census  White British - ethnic category 2001 census  Irish - ethnic category 2001 census  White Irish - ethnic category 2001 census  Other White background - ethnic category 2001 census  English - ethnic category 2001 census  Scottish - ethnic category 2001 census  Welsh - ethnic category 2001 census  Cornish - ethnic category 2001 census  Northern Irish - ethnic category 2001 census  Ulster Scots - ethnic category 2001 census  Cypriot (part not stated) - ethnic category 2001 census  Greek - ethnic category 2001 census  Greek Cypriot - ethnic category 2001 census  Turkish - ethnic category 2001 census  Turkish Cypriot - ethnic category 2001 census  Italian - ethnic category 2001 census  Irish Traveller - ethnic category 2001 census  Traveller - ethnic category 2001 census  Gypsy/Romany - ethnic category 2001 census  Polish - ethnic category 2001 census  Baltic Estonian/Latvian/Lithuanian - ethn categ 2001 census  Commonwealth (Russian) Indep States - ethn categ 2001 census  Kosovan - ethnic category 2001 census  Albanian - ethnic category 2001 census  Bosnian - ethnic category 2001 census  Croatian - ethnic category 2001 census  Serbian - ethnic category 2001 census  Other republics former Yugoslavia - ethnic categ 2001 census  Mixed Irish and other White - ethnic category 2001 census  Oth White European/Euro unsp/Mixed Euro 2001 census  Other mixed White - ethnic category 2001 census  Other White or White unspecified ethnic category 2001 census  White  White British  White Irish  Other white ethnic group  White Scottish  Other white British ethnic group  Irish (NMO)  Greek/Greek Cypriot (NMO)  Turkish/Turkish Cypriot (NMO)  Other European (NMO)  Irish traveller  White:Eng/Welsh/Scot/NI/Brit - England and Wales 2011 census  White: Irish - England and Wales ethnic category 2011 census  White: Gypsy/Irish Traveller - Eng+Wales eth cat 2011 census  White: other White backgrd- Eng+Wales ethnic cat 2011 census  White - Northern Ireland ethnic category 2011 census  Irish Traveller - Northern Ireland ethnic cat 2011 census  White: Scottish - Scotland ethnic category 2011 census  White: other British - Scotland ethnic category 2011 census  White: Irish - Scotland ethnic category 2011 census  White: Polish - Scotland ethnic category 2011 census  White: other White ethnic grp- Scotland ethnic cat 2011 cens |
| 2 Mixed | '9i3..'  '9i4..'  '9i5..'  '9i6..'  '9i60.'  '9i61.'  '9i62.'  '9i63.'  '9i65.'  '9iA7.'  '9S43.'  '9S46.'  '9S47.'  '9S48.'  '9S5..'  '9S51.'  '9S52.'  '9SA4.'  '9SA5.'  '9SA6.'  '9SB..'  '9SB1.'  '9SB2.'  '9SB3.'  '9SB4.'  '9SB5.'  '9SB6.'  '9t04.'  '9t05.'  '9t06.'  '9t07.'  '9t12.'  '9t13.'  '9t14.'  '9t15.'  '9t26.' | White and Black Caribbean - ethnic category 2001 census  White and Black African - ethnic category 2001 census  White and Asian - ethnic category 2001 census  Other Mixed background - ethnic category 2001 census  Black and Asian - ethnic category 2001 census  Black and Chinese - ethnic category 2001 census  Black and White - ethnic category 2001 census  Chinese and White - ethnic category 2001 census  Other Mixed or Mixed unspecified ethnic category 2001 census  Caribbean Asian - ethnic category 2001 census  Black N African/Arab/Iranian  Black Indian sub-continent  Black - other Asian  Black Black - other  Black - other, mixed  Other Black - Black/White orig  Other Black - Black/Asian orig  N African Arab/Iranian (NMO)  Other African countries (NMO)  E Afric Asian/Indo-Carib (NMO)  Other ethnic, mixed origin  Other ethnic, Black/White orig  Other ethnic, Asian/White orig  Other ethnic, mixed white orig  Other ethnic, other mixed orig  Black Caribbean and White  Black African and White  Mixed: White+Black Caribbean - Eng+Wales eth cat 2011 census  Mixed: White+Black African - Eng+Wales eth cat 2011 census  Mixed: White+Asian - Eng+Wales ethnic category 2011 census  Mixed: other Mixed/multiple backgrd - Eng+Wales 2011 census  Mixed: White and Black Caribbean - NI ethnic cat 2011 census  Mixed: White and Black African - NI ethnic cat 2011 census  Mixed: White and Asian - NI ethnic category 2011 census  Mixed: other Mixed/multiple ethnic backgrd - NI 2011 census  Mixed/multiple ethnic grps: any- Scot ethnic cat 2011 census |
| 3 Asian | '9i64.'  '9i7..'  '9i8..'  '9i9..'  '9iA..'  '9iA1.'  '9iA2.'  '9iA3.'  '9iA4.'  '9iA5.'  '9iA6.'  '9iA8.'  '9iA9.'  '9iAA.'  '9iE..'  '9iF0.'  '9iF1.'  '9iF2.'  '9iF3.'  '9S6..'  '9S7..'  '9S8..'  '9S9..'  '9SA7.'  '9SA8.'  '9SC..'  '9SH..'  '9t08.'  '9t09.'  '9t0A.'  '9t0B.'  '9t0C.'  '9t16.'  '9t17.'  '9t18.'  '9t19.'  '9t1A.'  '9t27.'  '9t28.'  '9t29.'  '9t2A.'  '9t2B.' | Asian and Chinese - ethnic category 2001 census  Indian or British Indian - ethnic category 2001 census  Pakistani or British Pakistani - ethnic category 2001 census  Bangladeshi or British Bangladeshi - ethn categ 2001 census  Other Asian background - ethnic category 2001 census  Punjabi - ethnic category 2001 census  Kashmiri - ethnic category 2001 census  East African Asian - ethnic category 2001 census  Sri Lankan - ethnic category 2001 census  Tamil - ethnic category 2001 census  Sinhalese - ethnic category 2001 census  British Asian - ethnic category 2001 census  Mixed Asian - ethnic category 2001 census  Other Asian or Asian unspecified ethnic category 2001 census  Chinese - ethnic category 2001 census  Vietnamese - ethnic category 2001 census  Japanese - ethnic category 2001 census  Filipino - ethnic category 2001 census  Malaysian - ethnic category 2001 census  Indian  Pakistani  Bangladeshi  Chinese  Indian sub-continent (NMO)  Other Asian (NMO)  Vietnamese  Other Asian ethnic group  Asian/Asian Brit: Indian - Eng+Wales ethnic cat 2011 census  Asian/Asian British:Pakistani- Eng+Wales eth cat 2011 census  Asian/Asian Brit: Bangladeshi- Eng+Wales eth cat 2011 census  Asian/Asian Brit: Chinese - Eng+Wales ethnic cat 2011 census  Asian/Asian Brit: other Asian- Eng+Wales eth cat 2011 census  Asian or Asian British: Indian - NI ethnic cat 2011 census  Asian/Asian British: Pakistani - NI ethnic cat 2011 census  Asian/Asian British: Bangladeshi - NI ethnic cat 2011 census  Asian/Asian British: Chinese - NI ethnic cat 2011 census  Asian/Asian British: other Asian - NI ethnic cat 2011 census  Asian: Pakistani/Pakistani Scot/Pakistani Brit- Scot 2011  Asian: Indian, Indian Scot/Indian Brit- Scotland 2011 census  Bangladeshi, Bangladeshi Scot or Bangladeshi Brit- Scot 2011  Asian: Chinese - Scotland ethnic category 2011 census  Asian: other Asian group - Scotland ethnic cat 2011 census |
| 4 Black | '9iB..'  '9iC..'  '9iD..'  '9iD0.'  '9iD1.'  '9iD2.'  '9iD3.'  '9iD4.'  '9iFA.'  '9S2..'  '9S3..'  '9S4..'  '9S41.'  '9S42.'  '9S44.'  '9S45.'  '9SA3.'  '9SG..'  '9t0D.'  '9t0E.'  '9t0F.'  '9t1B.'  '9t1C.'  '9t1D.'  '9t2C.'  '9t2D.'  '9t2E.'  '9t2F.' | Caribbean - ethnic category 2001 census  African - ethnic category 2001 census  Other Black background - ethnic category 2001 census  Somali - ethnic category 2001 census  Nigerian - ethnic category 2001 census  Black British - ethnic category 2001 census  Mixed Black - ethnic category 2001 census  Other Black or Black unspecified ethnic category 2001 census  North African - ethnic category 2001 census  Black Caribbean  Black African  Black, other, non-mixed origin  Black British  Black Caribbean/W.I./Guyana  Black - other African country  Black E Afric Asia/Indo-Caribb  Caribbean I./W.I./Guyana (NMO)  Other black ethnic group  Black/African/Carib/Black Brit: African- Eng+Wales 2011 cens  Black/African/Caribbn/Black Brit: Caribbean - Eng+Wales 2011  Black/Afr/Carib/Black Brit: other Black- Eng+Wales 2011 cens  Black/Afri/Carib/Black Brit: African- NI eth cat 2011 census  Black/Afri/Carib/Black Brit: Caribbean- NI eth cat 2011 cens  Black/Afri/Carib/Black Brit: other - NI eth cat 2011 census  African: African/African Scot/African Brit - Scotland 2011  African: any other African - Scotland ethnic cat 2011 census  Carib/Black: Caribbean/Carib Scot/Carib Brit- Scotland 2011  Carib/Black: Black/Black Scot/Black Brit- Scotland 2011 cens |
| 5 Other | '9iF..'  '9iF4.'  '9iF5.'  '9iF6.'  '9iF7.'  '9iF8.'  '9iF9.'  '9iFB.'  '9iFC.'  '9iFD.'  '9iFE.'  '9iFF.'  '9iFG.'  '9iFH.'  '9iFJ.'  '9iFK.'  '9SA..'  '9SAD.'  '9SJ..'  '9t0G.'  '9t0H.'  '9t1E.'  '9t1F.'  '9t2H.'  '9t2J.' | Other - ethnic category 2001 census  Buddhist - ethnic category 2001 census  Hindu - ethnic category 2001 census  Jewish - ethnic category 2001 census  Muslim - ethnic category 2001 census  Sikh - ethnic category 2001 census  Arab - ethnic category 2001 census  Mid East (excl Israeli, Iranian & Arab) - eth cat 2001 cens  Israeli - ethnic category 2001 census  Iranian - ethnic category 2001 census  Kurdish - ethnic category 2001 census  Moroccan - ethnic category 2001 census  Latin American - ethnic category 2001 census  South and Central American - ethnic category 2001 census  Mauritian/Seychellois/Maldivian/St Helena eth cat 2001census  Any other group - ethnic category 2001 census  Other ethnic non-mixed (NMO)  Other ethnic NEC (NMO)  Other ethnic group  Other ethnic group: Arab - Eng+Wales ethnic cat 2011 census  Other ethnic: any other grp - Eng+Wales eth cat 2011 census  Other ethnic group: Arab - NI ethnic category 2011 census  Other ethnic group: any other grp- NI ethnic cat 2011 census  Other ethnic grp: Arab/Arab Scot/Arab British- Scotland 2011  Other ethnic grp: any other ethnic grp- Scotland 2011 census |

| NER category | Data source code | Data source description |
| --- | --- | --- |
| 1 White | '9i0..'  '9i00.'  '9i1..'  '9i10.'  '9i2..'  '9i20.'  '9i21.'  '9i22.'  '9i23.'  '9i24.'  '9i25.'  '9i26.'  '9i27.'  '9i28.'  '9i29.'  '9i2A.'  '9i2B.'  '9i2C.'  '9i2D.'  '9i2E.'  '9i2F.'  '9i2G.'  '9i2H.'  '9i2J.'  '9i2K.'  '9i2L.'  '9i2M.'  '9i2N.'  '9i2P.'  '9i2Q.'  '9i2R.'  '9i2S.'  '9i2T.'  '9S1..'  '9S10.'  '9S11.'  '9S12.'  '9S13.'  '9S14.'  '9SA9.'  '9SAA.'  '9SAB.'  '9SAC.'  '9SI..'  '9t00.'  '9t01.'  '9t02.'  '9t03.'  '9t10.'  '9t11.'  '9t20.'  '9t21.'  '9t22.'  '9t24.'  '9t25.' | British or mixed British - ethnic category 2001 census  White British - ethnic category 2001 census  Irish - ethnic category 2001 census  White Irish - ethnic category 2001 census  Other White background - ethnic category 2001 census  English - ethnic category 2001 census  Scottish - ethnic category 2001 census  Welsh - ethnic category 2001 census  Cornish - ethnic category 2001 census  Northern Irish - ethnic category 2001 census  Ulster Scots - ethnic category 2001 census  Cypriot (part not stated) - ethnic category 2001 census  Greek - ethnic category 2001 census  Greek Cypriot - ethnic category 2001 census  Turkish - ethnic category 2001 census  Turkish Cypriot - ethnic category 2001 census  Italian - ethnic category 2001 census  Irish Traveller - ethnic category 2001 census  Traveller - ethnic category 2001 census  Gypsy/Romany - ethnic category 2001 census  Polish - ethnic category 2001 census  Baltic Estonian/Latvian/Lithuanian - ethn categ 2001 census  Commonwealth (Russian) Indep States - ethn categ 2001 census  Kosovan - ethnic category 2001 census  Albanian - ethnic category 2001 census  Bosnian - ethnic category 2001 census  Croatian - ethnic category 2001 census  Serbian - ethnic category 2001 census  Other republics former Yugoslavia - ethnic categ 2001 census  Mixed Irish and other White - ethnic category 2001 census  Oth White European/Euro unsp/Mixed Euro 2001 census  Other mixed White - ethnic category 2001 census  Other White or White unspecified ethnic category 2001 census  White  White British  White Irish  Other white ethnic group  White Scottish  Other white British ethnic group  Irish (NMO)  Greek/Greek Cypriot (NMO)  Turkish/Turkish Cypriot (NMO)  Other European (NMO)  Irish traveller  White:Eng/Welsh/Scot/NI/Brit - England and Wales 2011 census  White: Irish - England and Wales ethnic category 2011 census  White: Gypsy/Irish Traveller - Eng+Wales eth cat 2011 census  White: other White backgrd- Eng+Wales ethnic cat 2011 census  White - Northern Ireland ethnic category 2011 census  Irish Traveller - Northern Ireland ethnic cat 2011 census  White: Scottish - Scotland ethnic category 2011 census  White: other British - Scotland ethnic category 2011 census  White: Irish - Scotland ethnic category 2011 census  White: Polish - Scotland ethnic category 2011 census  White: other White ethnic grp- Scotland ethnic cat 2011 cens |
| 2 Mixed | '9i3..'  '9i4..'  '9i5..'  '9i6..'  '9i60.'  '9i61.'  '9i62.'  '9i63.'  '9i65.'  '9iA7.'  '9S43.'  '9S46.'  '9S47.'  '9S48.'  '9S5..'  '9S51.'  '9S52.'  '9SA4.'  '9SA5.'  '9SA6.'  '9SB..'  '9SB1.'  '9SB2.'  '9SB3.'  '9SB4.'  '9SB5.'  '9SB6.'  '9t04.'  '9t05.'  '9t06.'  '9t07.'  '9t12.'  '9t13.'  '9t14.'  '9t15.'  '9t26.' | White and Black Caribbean - ethnic category 2001 census  White and Black African - ethnic category 2001 census  White and Asian - ethnic category 2001 census  Other Mixed background - ethnic category 2001 census  Black and Asian - ethnic category 2001 census  Black and Chinese - ethnic category 2001 census  Black and White - ethnic category 2001 census  Chinese and White - ethnic category 2001 census  Other Mixed or Mixed unspecified ethnic category 2001 census  Caribbean Asian - ethnic category 2001 census  Black N African/Arab/Iranian  Black Indian sub-continent  Black - other Asian  Black Black - other  Black - other, mixed  Other Black - Black/White orig  Other Black - Black/Asian orig  N African Arab/Iranian (NMO)  Other African countries (NMO)  E Afric Asian/Indo-Carib (NMO)  Other ethnic, mixed origin  Other ethnic, Black/White orig  Other ethnic, Asian/White orig  Other ethnic, mixed white orig  Other ethnic, other mixed orig  Black Caribbean and White  Black African and White  Mixed: White+Black Caribbean - Eng+Wales eth cat 2011 census  Mixed: White+Black African - Eng+Wales eth cat 2011 census  Mixed: White+Asian - Eng+Wales ethnic category 2011 census  Mixed: other Mixed/multiple backgrd - Eng+Wales 2011 census  Mixed: White and Black Caribbean - NI ethnic cat 2011 census  Mixed: White and Black African - NI ethnic cat 2011 census  Mixed: White and Asian - NI ethnic category 2011 census  Mixed: other Mixed/multiple ethnic backgrd - NI 2011 census  Mixed/multiple ethnic grps: any- Scot ethnic cat 2011 census |
| 3 Indian | '9i7..'  '9S6..'  '9SA7.'  '9t08.'  '9t16.'  '9t28.' | Indian or British Indian - ethnic category 2001 census  Indian  Indian sub-continent (NMO)  Asian/Asian Brit: Indian - Eng+Wales ethnic cat 2011 census  Asian or Asian British: Indian - NI ethnic cat 2011 census  Asian: Indian, Indian Scot/Indian Brit- Scotland 2011 census |
| 4 Pakistani | '9i8..'  '9S7..'  '9t09.'  '9t17.'  '9t27.' | Pakistani or British Pakistani - ethnic category 2001 census  Pakistani  Asian/Asian British:Pakistani- Eng+Wales eth cat 2011 census  Asian/Asian British: Pakistani - NI ethnic cat 2011 census  Asian: Pakistani/Pakistani Scot/Pakistani Brit- Scot 2011 |
| 5 Bangladeshi | '9i9..'  '9S8..'  '9t0A.'  '9t18.'  '9t29.' | Bangladeshi or British Bangladeshi - ethn categ 2001 census  Bangladeshi  Asian/Asian Brit: Bangladeshi- Eng+Wales eth cat 2011 census  Asian/Asian British: Bangladeshi - NI ethnic cat 2011 census  Bangladeshi, Bangladeshi Scot or Bangladeshi Brit- Scot 2011 |
| 6 Chinese | '9iE..'  '9S9..'  '9t0B.'  '9t19.'  '9t2A.' | Chinese - ethnic category 2001 census  Chinese  Asian/Asian Brit: Chinese - Eng+Wales ethnic cat 2011 census  Asian/Asian British: Chinese - NI ethnic cat 2011 census  Asian: Chinese - Scotland ethnic category 2011 census |
| 7 Black Caribbean | '9iB..'  '9S2..'  '9S42.'  '9SA3.'  '9t2E.'  '9t2F.' | Caribbean - ethnic category 2001 census  Black Caribbean  Black Caribbean/W.I./Guyana  Caribbean I./W.I./Guyana (NMO)  Carib/Black: Caribbean/Carib Scot/Carib Brit- Scotland 2011  Carib/Black: Black/Black Scot/Black Brit- Scotland 2011 cens |
| 8 Black African | '9iC..'  '9iD0.'  '9iD1.'  '9iFA.'  '9S3..'  '9S44.'  '9t2C.'  '9t2D.' | African - ethnic category 2001 census  Somali - ethnic category 2001 census  Nigerian - ethnic category 2001 census  North African - ethnic category 2001 census  Black African  Black - other African country  African: African/African Scot/African Brit - Scotland 2011  African: any other African - Scotland ethnic cat 2011 census |
| 9 Other | '9i64.'  '9iA..'  '9iA1.'  '9iA2.'  '9iA3.'  '9iA4.'  '9iA5.'  '9iA6.'  '9iA8.'  '9iA9.'  '9iAA.'  '9iD..'  '9iD2.'  '9iD3.'  '9iD4.'  '9iF..'  '9iF0.'  '9iF1.'  '9iF2.'  '9iF3.'  '9iF4.'  '9iF5.'  '9iF6.'  '9iF7.'  '9iF8.'  '9iF9.'  '9iFB.'  '9iFC.'  '9iFD.'  '9iFE.'  '9iFF.'  '9iFG.'  '9iFH.'  '9iFJ.'  '9iFK.'  '9S4..'  '9S41.'  '9S45.'  '9SA..'  '9SA8.'  '9SAD.'  '9SC..'  '9SG..'  '9SH..'  '9SJ..'  '9t0C.'  '9t0D.'  '9t0E.'  '9t0F.'  '9t0G.'  '9t0H.'  '9t1A.'  '9t1B.'  '9t1C.'  '9t1D.'  '9t2B.'  '9t1E.'  '9t1F.'  '9t2H.'  '9t2J.' | Asian and Chinese - ethnic category 2001 census  Other Asian background - ethnic category 2001 census  Punjabi - ethnic category 2001 census  Kashmiri - ethnic category 2001 census  East African Asian - ethnic category 2001 census  Sri Lankan - ethnic category 2001 census  Tamil - ethnic category 2001 census  Sinhalese - ethnic category 2001 census  British Asian - ethnic category 2001 census  Mixed Asian - ethnic category 2001 census  Other Asian or Asian unspecified ethnic category 2001 census  Other Black background - ethnic category 2001 census  Black British - ethnic category 2001 census  Mixed Black - ethnic category 2001 census  Other Black or Black unspecified ethnic category 2001 census  Other - ethnic category 2001 census  Vietnamese - ethnic category 2001 census  Japanese - ethnic category 2001 census  Filipino - ethnic category 2001 census  Malaysian - ethnic category 2001 census  Buddhist - ethnic category 2001 census  Hindu - ethnic category 2001 census  Jewish - ethnic category 2001 census  Muslim - ethnic category 2001 census  Sikh - ethnic category 2001 census  Arab - ethnic category 2001 census  Mid East (excl Israeli, Iranian & Arab) - eth cat 2001 cens  Israeli - ethnic category 2001 census  Iranian - ethnic category 2001 census  Kurdish - ethnic category 2001 census  Moroccan - ethnic category 2001 census  Latin American - ethnic category 2001 census  South and Central American - ethnic category 2001 census  Mauritian/Seychellois/Maldivian/St Helena eth cat 2001census  Any other group - ethnic category 2001 census  Black, other, non-mixed origin  Black British  Black E Afric Asia/Indo-Caribb  Other ethnic non-mixed (NMO)  Other Asian (NMO)  Other ethnic NEC (NMO)  Vietnamese  Other black ethnic group  Other Asian ethnic group  Other ethnic group  Asian/Asian Brit: other Asian- Eng+Wales eth cat 2011 census  Black/African/Carib/Black Brit: African- Eng+Wales 2011 cens  Black/African/Caribbn/Black Brit: Caribbean - Eng+Wales 2011  Black/Afr/Carib/Black Brit: other Black- Eng+Wales 2011 cens  Other ethnic group: Arab - Eng+Wales ethnic cat 2011 census  Other ethnic: any other grp - Eng+Wales eth cat 2011 census  Asian/Asian British: other Asian - NI ethnic cat 2011 census  Black/Afri/Carib/Black Brit: African- NI eth cat 2011 census  Black/Afri/Carib/Black Brit: Caribbean- NI eth cat 2011 cens  Black/Afri/Carib/Black Brit: other - NI eth cat 2011 census  Asian: other Asian group - Scotland ethnic cat 2011 census  Other ethnic group: Arab - NI ethnic category 2011 census  Other ethnic group: any other grp- NI ethnic cat 2011 census  Other ethnic grp: Arab/Arab Scot/Arab British- Scotland 2011  Other ethnic grp: any other ethnic grp- Scotland 2011 census |
